# Decoding the Deadly Dance: NETosis Genes Predict Neonatal Sepsis Fate

**DOI:** 10.1101/2024.09.10.24313397

**Authors:** Deepshikha Shaw, Sridhar Santhanam, Tapas Kumar Som, Samsiddhi Bhattacharjee, Saroj Kant Mohapatra

## Abstract

**Background:** Neonatal sepsis, a systemic inflammatory response to infection, is a major cause of morbidity and mortality in newborns. Neutrophil extracellular trap formation (NETosis), while crucial for pathogen clearance, can contribute to organ dysfunction in sepsis. This study aimed to identify key NETosis-related genes for prognostication in neonatal sepsis.

**Methods:** We analysed whole blood transcriptome datasets (GSE26440, GSE26378, GSE25504) from neonates with sepsis and controls. Differentially expressed NETosis genes (DE-NET genes) were identified, and a machine learning approach was used to select the most influential genes. A NET score model was constructed and validated using single-sample gene set enrichment analysis (ssGSEA). The model’s performance was evaluated using ROC analysis. The interplay between key-NET genes and the complement-coagulation (CC) system was investigated. Clinical samples were also collected for validation.

**Results:** Sixteen DE-NET genes were identified, and LASSO further refined these to 8 key-NET genes. The key-NET gene signature and NET score model showed excellent predictive performance (AUCs > 89%) in distinguishing survivors from non-survivors. Mediation analysis revealed that key-NET gene expression precedes and potentially drives complement-coagulation activation.

**Conclusions:** We present an 8-gene prognostic model for risk stratification in neonatal sepsis, based on early blood transcript signatures in neonates. Our findings underscore the central role of NETosis in sepsis-induced coagulopathy, revealing potential therapeutic targets for intervention.

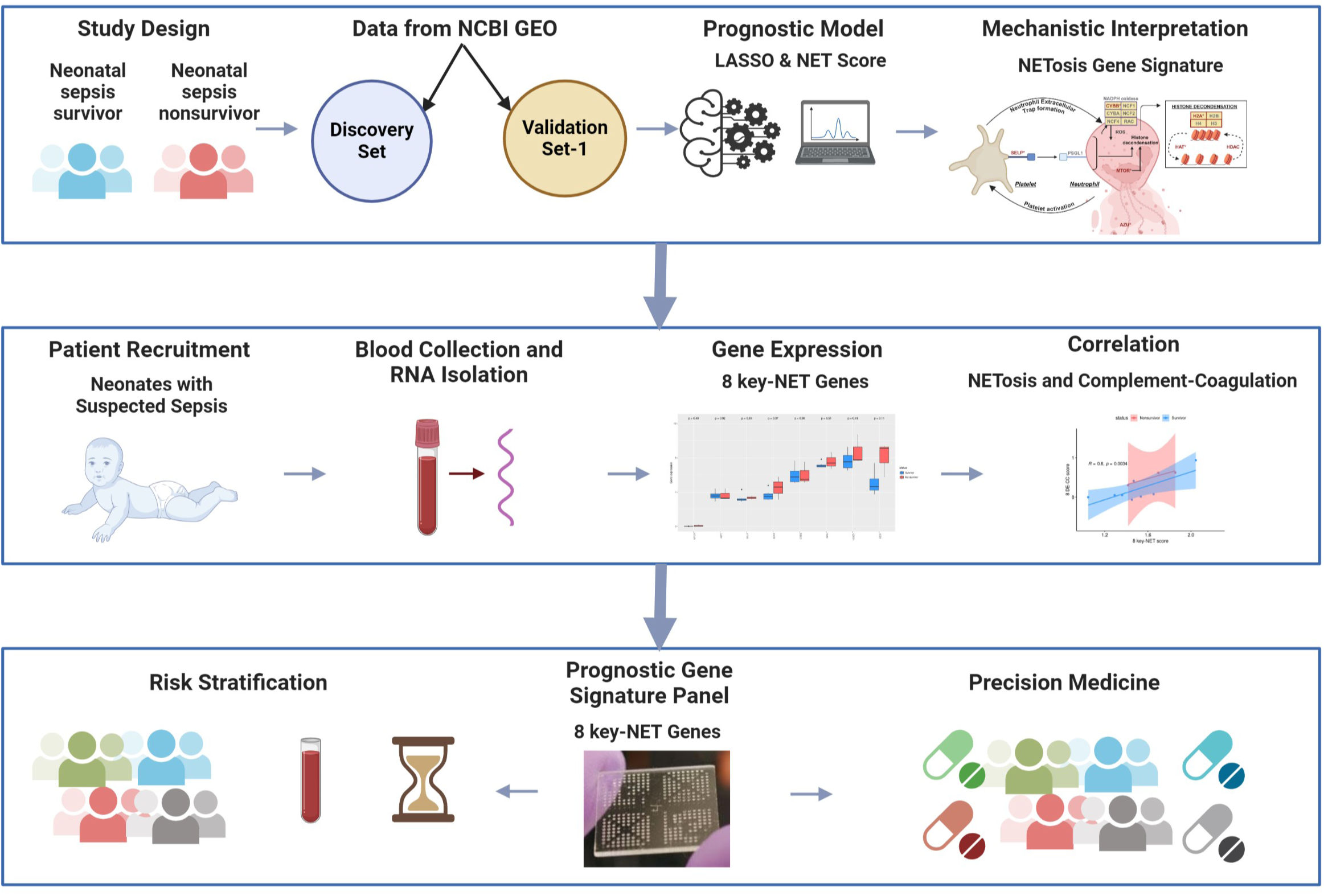

## 1. Introduction

Sepsis, a silent killer claiming millions of lives each year, poses a dire threat to global health, particularly for newborns. This life-threatening condition arises from a dysregulated host response to infection, potentially leading to organ dysfunction and death (1). Neonatal sepsis occurs when newborns are infected by pathogens(2). Sepsis in neonates differs substantially from that in adult and older children since they have altered immune function and the potential intrauterine exposure to infection(3–5). The global estimate for neonatal sepsis is 3 million cases per year (22 per 1000 livebirths), with mortality between 11%-19%(6). Neonatal sepsis can be the result of infections with bacterial, viral, or fungal (yeast) microorganisms(2,7). The microbiological approach to defining neonatal sepsis is increasingly being reconsidered. Current emphasis is shifting toward recognizing a dysregulated host response to infection, characterized by an acute onset of clinical or laboratory evidence of organ dysfunction, rather than relying solely on pathogen detection as the primary diagnostic criterion for sepsis(1,8). The host response to infection, if dysregulated, can result in self-inflicted damage to cells and tissues, culminating in multiple organ failure and potentially death(9). However, the pathophysiological processes of neonatal sepsis, as well as effective therapeutic strategies, remain insufficiently explored. Evidence suggests that mortality in neonatal sepsis is closely linked to dysregulation of the pro-inflammatory response(10), highlighting the importance of the acute pro-inflammatory phase in prognosis and therapeutic intervention. Therefore, it is critical to rigorously investigate the molecular changes that occur during the pathogenesis of neonatal sepsis and to identify novel and effective algorithms for prognosis and therapeutic options. The complexity of the transcriptional response to sepsis presents a research bottleneck, necessitating advanced analytical approaches. Bioinformatics and gene expression profiling offer powerful tools to unravel this complexity, enabling the identification of molecular targets that could inform critical care strategies.

In this context, neutrophils, one of the most abundant immune cell subsets, emerge as key players in the early immune response to sepsis. Neutrophils release de-adherent DNA chromatin into the extracellular space, forming reticular structures known as neutrophil extracellular traps (NETs)(11). While NETs serve to physically capture bacteria and enhance pathogen-antimicrobial interactions (12), their formation also acts as a double-edged sword in sepsis. Excessive NET formation can lead to tissue damage and inflammation, driving the progression to multiorgan failure (13,14). Major components of NETs function as Damage-Associated Molecular Patterns (DAMPs), further exacerbating the inflammatory response and triggering a harmful cascade of continued NET release (15). Thus, NETs are considered critical contributors to organ failure during sepsis.

Building on these insights, we have developed a prognostic model centered on NET formation to stratify outcomes in neonatal sepsis. By integrating multiple high-throughput transcriptomic datasets, our study explores the interplay between NETosis and the complement-coagulation pathway, highlighting the central role of NETosis in sepsis pathogenesis and offering new avenues for therapeutic intervention.

## 2. Methods

### 2.1. Selection and preprocessing of the data

We systematically queried and filtered datasets for inclusion in our analysis (Figure 1). Three primary datasets (GSE26440, GSE26378, and GSE25504) were downloaded from NCBI GEO database(16), which include gene expression profiles along with corresponding clinical information (Supplementary Table 1). GSE26440 (1 platform), GSE26378 (1 platform) and GSE25504 (3 platforms) contain gene expression profiles of neonatal sepsis samples. All five blocks of datasets were merged using the “ComBat” algorithm from sva(17) package in R to adjust for batch effects (Supplementary Figure 1 A-F). The merged dataset included control samples (n=74) and neonatal sepsis samples (n=109). The neonatal sepsis samples were divided into a discovery set (56 survivors and 10 non-survivors) and validation set-1 (37 survivors and 6 non-survivors). External validation sets were used, validation set-2 for temporal data analysis, validation set-3 for pediatric sepsis, validation set-4 for adult sepsis (Supplementary Table 2). Validation set-5 consisted of cases of neonatal sepsis recruited prospectively by our laboratory (Supplementary Table 3).

**Figure 1:**
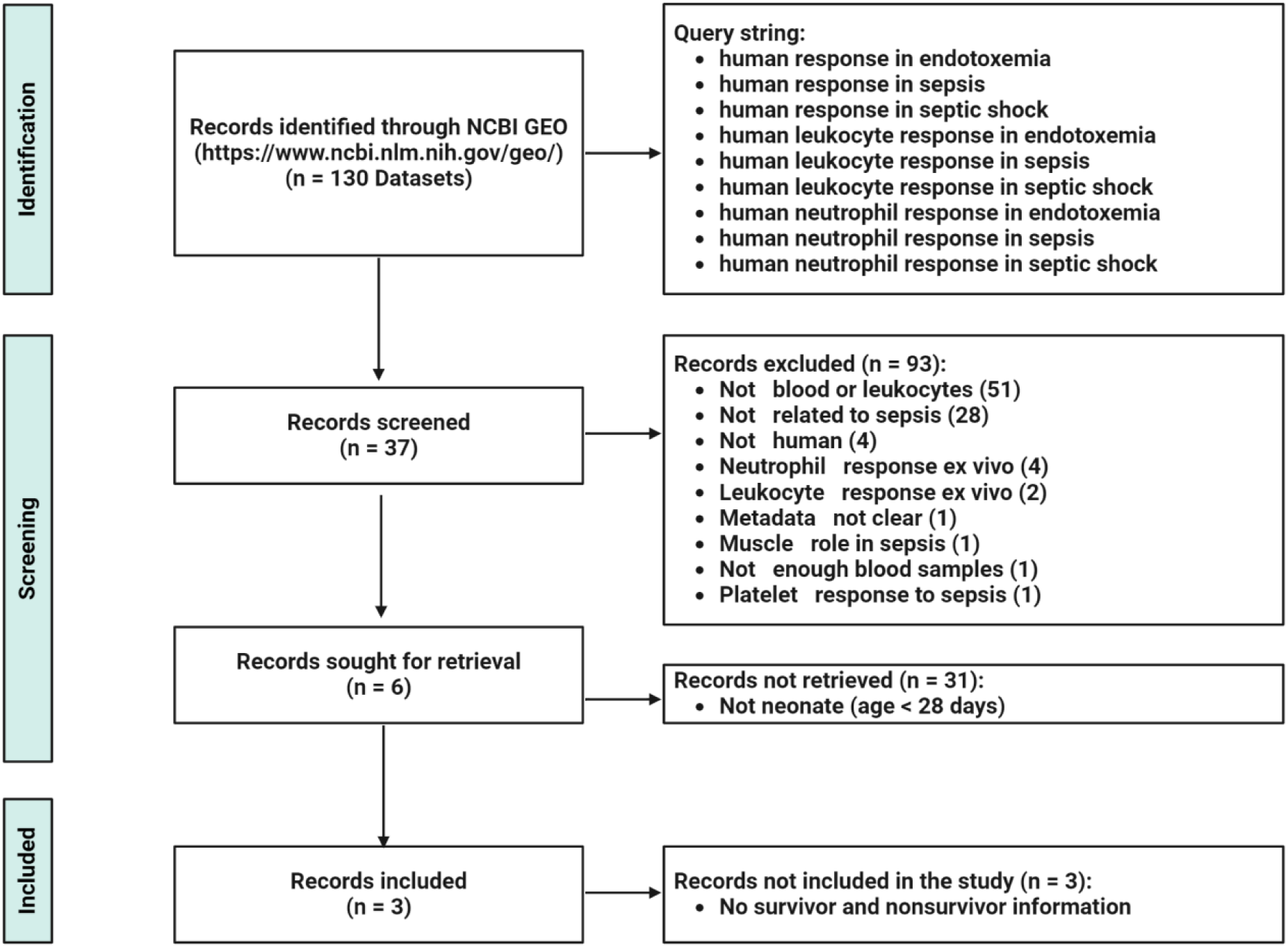
PRISMA flow chart of the transcriptional meta-analysis. Identification, screening and inclusion of eligible GEO records (datasets) for this study.

### 2.2. Permutation-based Gene Set Enrichment Analysis (GSEA)

For each gene, a t-statistic was computed to denote the change in gene expression between groups. For each pathway, a score was calculated through weighted averaging – the sum of the gene-level t-statistics divided by the square root of the number of genes in the pathway. Significance of the observed pathway score was determined by permutation testing, performed as follows: In each permutation, samples were randomly relabelled as neonatal sepsis survivor or neonatal sepsis non-survivor, and a score for a simulated pathway was calculated by weighted averaging as described above. This process was repeated 10,000 times generating 10,000 simulated pathway scores to represent the null distribution of the pathway score. The deviation of the observed pathway score from the null distribution was quantified by the proportion of times the simulated score was more extreme than the observed score. This result was assigned as a permutation p-value of the observed pathway score. Pathway enrichment analysis was performed using “Category” package(18) in R.

### 2.3. Selection and analysis of Differentially Expressed NETosis genes (DE-NET genes)

Genes functional in NETosis pathway were retrieved from the Kyoto Encyclopedia of Genes and Genomes (KEGG)(19). The genes associated with the NETosis pathway (KEGG ID: hsa04613) were analysed for differential expression. Genes exhibiting a |log (fold change) | ≥ 0.5 and p-value <0.05 were identified as differentially expressed NETosis genes (DE-NET genes) between neonatal sepsis non-survivors and neonatal sepsis survivors.

### 2.4. Pruning DE-NET genes using the least absolute shrinkage and selection operator (LASSO) method

To further define the most informative set of genes out of 16 DE-NET genes, we applied the LASSO(20) (Least Absolute Shrinkage and Selection Operator) algorithm to construct an optimal model. This process was conducted using the R software and the ‘glmnet’(21) package. The LASSO algorithm operates by applying a penalty function that shrinks the regression coefficients of variables, effectively setting coefficients of irrelevant or redundant features to zero. To select the penalty parameters (optimal lambda), we employed five-fold cross-validation. Based on the best lambda value, the key-NET genes (prognostic signature) were identified by refitting the model with the gene expression data. The validity of the Key-NET genes was confirmed using validation set-1 and their prognostic power was assessed by calculating the area under the receiver operating characteristic curve (ROC) in the discovery and validation set-1 using the “pROC”(22) package in R. The significance of the area under the ROC curve (AUC) was determined through permutation testing, in which the samples were randomly re-labelled as neonatal sepsis survivors or non-survivors (10,000 iterations).

### 2.5. Construction and validation of the NET Score model using single sample GSEA (ssGSEA)

Eight key NETosis genes (key-NET genes) were included in subsequent analyses. The single-sample Gene Set Enrichment Analysis(23) (ssGSEA) from GSVA(24) package in R computes a gene set enrichment score for each sample (referred to as the NET Score) by calculating the normalized difference in empirical cumulative distribution functions of gene expression ranks within and outside the gene set. The predictive power of the NET score model was assessed by the AUC in discovery and validation set-1. The significance of the AUC was determined through permutation testing, wherein samples were randomly re-labelled as sepsis survivors or non-survivors 10,000 times.

### 2.6. Functional Enrichment Analysis of DE-NET Genes and Key-NET Genes

Functional enrichment analysis was performed on DE-NET genes and key-NET genes using the R package “clusterProfiler”(25). This analysis was performed to assess the biological significance (Biological Process) of these genes through Gene Ontology(26) (GO) term enrichment.

### 2.7. Correlation Analysis of Key-NET Genes and DE-CC Genes

DE-CC genes were identified in the same way as DE-NET genes were identified, using the same thresholds of |log (fold change) | ≥ 0.5 and p-value <0.05. CC scores were calculated as NET scores were calculated using ssGSEA algorithm for each sample. Pearson correlation coefficient was calculated to assess the relationship between two pathway scores: Neutrophil Extracellular Trap Formation (NET score) and the Complement Coagulation Pathway (CC score).

### 2.8. Mediation Analysis

We tested for mediation(27) effects within the association triangles formed by the NET score, CC score and survival status using the R package “Lavaan”(28). In these analyses, survival status was consistently treated as the outcome, while the NET score and the CC score were treated as the exposure and mediator, respectively. Conceptually, mediation analysis serves as a third-variable analysis, examining the mediating effect of a variable (the mediator) on the relationship between two other variables, i.e., the association between exposure and outcome.

### 2.9. Construction of Protein-Protein Interaction (PPI) network

The STRING(29) database (https://string-db.org, version 12.0) online tool was used to predict and visualize protein-protein interaction (PPI) network models based on the sixteen DE- NET genes and the eight key-NET genes separately. The construction parameters included interaction sources such as text mining, experiments, databases, and co-expression, with the species restricted to *Homo sapiens* and a minimum interaction score threshold of 0.4.

### 2.10. Deconvolution of immune cells

CIBERSORTx(30) online tool (https://cibersortx.stanford.edu/) is an algorithm that uses gene expression based deconvolution to infer 22 human immune cell types, and applies marker gene signatures to quantify the relative scores for each cell type. To enhance the robustness of the results, CIBERSORTx uses Monte Carlo sampling to obtain deconvolution p values for each sample, and the abundance of immune cells can also be estimated based on gene expression data. Here, gene expression matrix data were uploaded to CIBERSORTx and differences with p value < 0.05 considered significant. Pearson’s correlation analysis was employed to assess the relationship between the expression of key-NET genes and the abundance scores of different immune cell types in neonatal sepsis survivors and neonatal sepsis nonsurvivors.

### 2.11. Recruitment of neonatal sepsis patients and whole blood transcriptome profiling

Neonates with sepsis were recruited by neonatologists at the Christian Medical College and Hospital, Vellore (CMC Vellore), and the All India Institute of Medical Sciences, Bhubaneswar (AIIMS BBSR), using the following inclusion criteria: Newborn babies (birth to 28 days) with one or more of the following clinical features (1) temperature instability (hypothermia, hyperthermia), (2) respiratory alterations (grunting, intercostal retractions, apnea, tachypnea, cyanosis), (3) cardiovascular alterations (bradycardia, tachycardia, poor perfusion, hypotension), (4) neurologic alterations (hypotonia, lethargy, seizures), (5) gastrointestinal alterations (feeding intolerance, abdominal distension), (6) suspicion of sepsis by the clinician with a decision to start antibiotic. Both term and pre-term babies were included. Babies with major congenital malformation were excluded. This set included 11 samples from neonatal sepsis patients and 2 samples from control subjects (evaluated for sepsis but with negative blood cultures and sepsis screens, leading to antibiotic discontinuation after 48-72 hours). 1.0 mL of blood was collected from each neonate via venipuncture or arterial stab at the time that the baby became symptomatic into designated tubes containing PAXgene blood RNA reagent (BD Cat No. 762165). Laboratory parameters are provided in Supplementary Table 3. Total RNA was extracted from whole blood received in PAXgene Blood RNA tubes using PAXgene Blood RNA kit (PreAnalytiX Cat No. 762174). Transcriptome profiling was done on gene expression microarray platform (Affymetrix) using Human Gene 2.0 ST Array, or on Nanopore PromethION system (PromethION P24) using PromethION flow cell (FLO- PRO114M).

### 2.12. Statistical Analysis and Data Visualization

The statistical significance of all box plots was evaluated using a two-tailed unpaired Student’s t-test and for all correlation analysis Pearson’s correlation was used. All statistical analyses, except otherwise mentioned, were performed in R software v.4.3.1(31). P value < 0.05 was considered significant. The following R packages were used for data visualization: “pheatmap”(32) (for heat maps), “corrplot”(33) (for correlation matrix), “EnhancedVolcano”(34) (for volcano plot), “ggplot2”(35) (for permutation histograms, box plots, scatter plots and ROC plots).

## 3. Results

### 3.1. Upregulation of NETosis in nonsurvivors of neonatal sepsis patients

A detailed analysis flowchart of this study is shown in Figure 2. Targeted analysis was conducted to examine the differential expression of genes functional in the NETosis pathway (KEGG ID: hsa04613). Gene set enrichment analysis, utilizing permutation testing, revealed that genes functional in NETosis are significantly upregulated in the whole blood of neonatal sepsis patients compared to healthy subjects (Supplementary Figure 2 A-B). Furthermore, there is up- regulation of the NETosis pathway in non-survivors compared to sepsis survivors as demonstrated for the discovery set (Figure 3 A) and validation set-1 (Figure 3 B).

**Figure 2:**
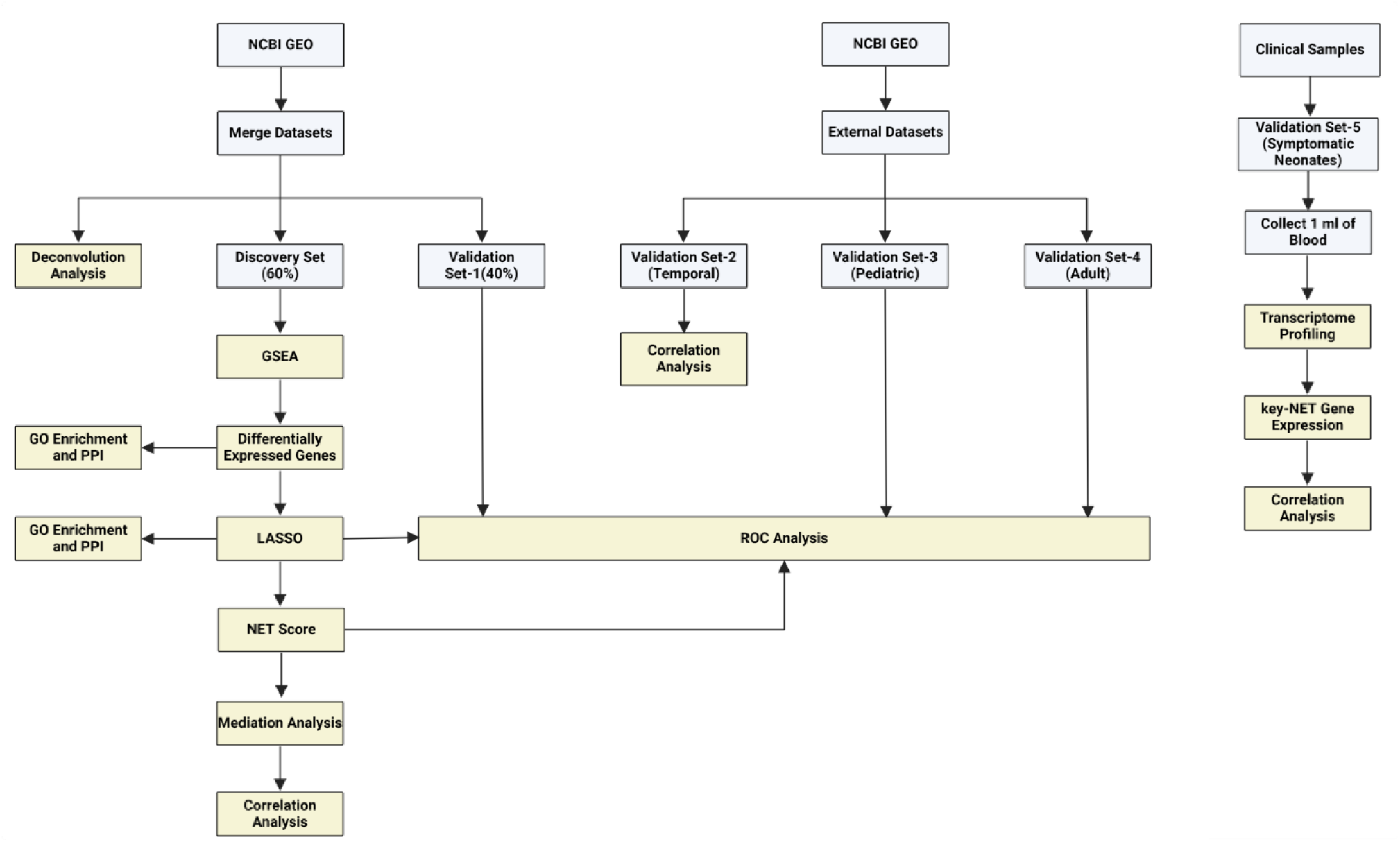
Analysis flow chart. GEO: Gene Expression Omnibus, GO: Gene Ontology, GSEA: Gene Set Enrichment Analysis, LASSO: Least Absolute Shrinkage and Selection Operator, NCBI: National Center for Biotechnology Information, NET: Neutrophil Extracellular Trap, PPI: Protein Protein Interaction network, ROC: Receiver-operating characteristic curve.

**Figure 3:**
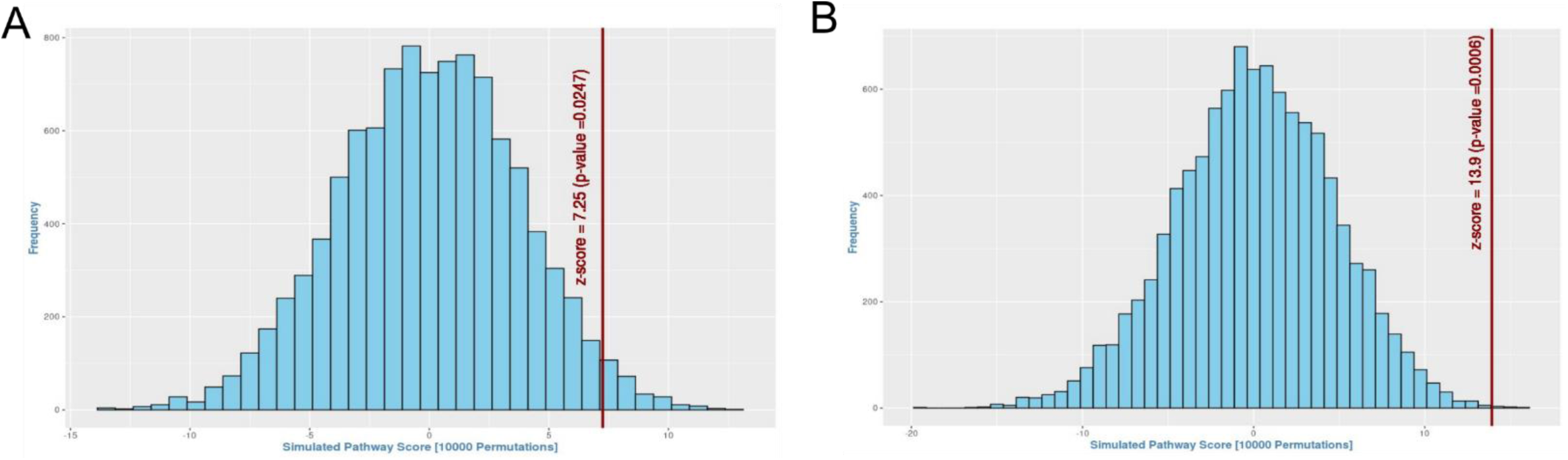
Upregulation of gene set functional in NETosis in blood of patients from the sepsis nonsurvivor group compared to the survivor group. (A) For the gene set functional in NETosis, pathway score (red vertical line) was calculated by weighted averaging of t-statistic in Discovery set between sepsis survivor group (n=56) and sepsis nonsurvivor group (n=10), as described in the Methods section. The histogram (blue bars) represents the null distribution of the pathway score calculated for each of the 10,000 iterations of permuting the sample labels. The position of the red vertical line (observed pathway score) with respect to the histogram (null distribution) suggests that the pathway is significantly upregulated (p = 0.025) in the blood of sepsis nonsurvivor. (B) Upregulation of NETosis pathway score (p = 0.001) was observed between sepsis nonsurvivor (n=6) and sepsis survivor (n=37) was further validated in validation set-1.

### 3.2. Identification of Differentially Expressed genes functional in NETosis

To identify neonatal sepsis-associated NETosis genes with a high mortality risk, we analysed differentially expressed genes (DEGs) between sepsis survivors and sepsis non-survivors in the discovery set. Based on the screening criteria of |log (fold change) | ≥ 0.5 and p value < 0.05, a total of 16 NETosis genes (DE-NET genes) including HAT1, SELP, H2AX, MTOR, CYBB, AZU1, H2AZ1, CTSG, H2AJ, HDAC2, HMGB1, ITGB3, MPO, PIK3CB, MAP2K1, and MACROH2A1 were observed to be upregulated (Figure 4A and Supplementary Table 4). Figure 4 B and Supplementary Figure 3 shows heatmap of 16 DE-NETs in discovery and validation set-1 respectively.

**Figure 4:**
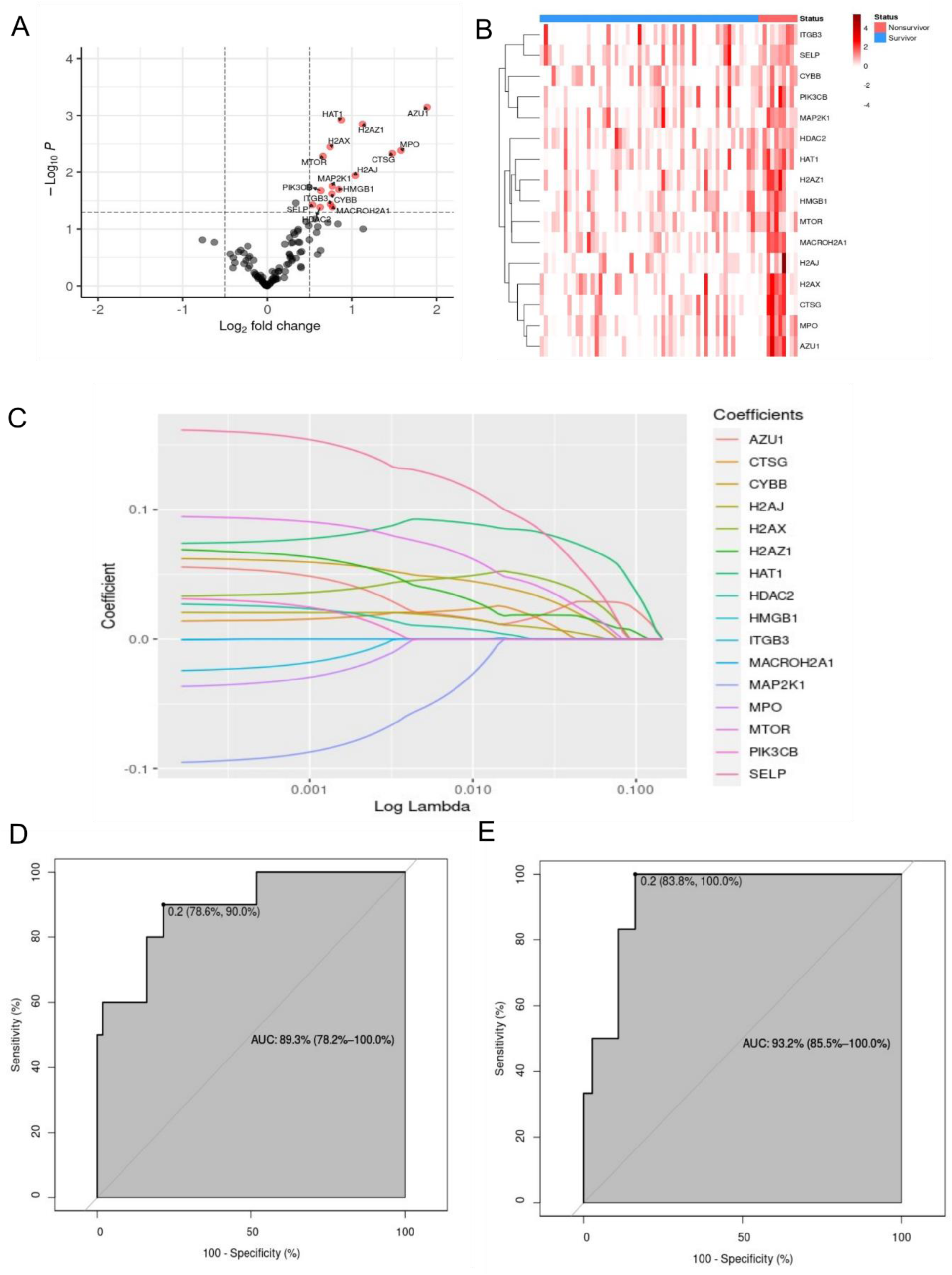
Identification of differentially expressed NETosis genes (DE-NETs) and key-NET genes in neonatal sepsis. (A) Volcano plot showing increased up-regulation of DE-NETs (red dots) in sepsis nonsurvivor group (n=10) compared to the sepsis survivor group (n=56) in the discovery set. (B) Expression of DE-NET genes in discovery set. (C) LASSO coefficient profiles of the key-NET genes selected by the optimal Lambda. (D-E) ROC analysis of the key-NETs for prognosis of neonatal sepsis nonsurvivor in (D) discovery set and (E) validation set-1.

### 3.3. Pruning DE-NET genes using Least Absolute Shrinkage and Selection Operator (LASSO)

To refine the prognostic signature further, we applied a LASSO regression model to analyse the 16 DE-NET genes in the discovery set (Figure 4 C). The regression coefficients are presented in Supplementary Table 5. We used five-fold cross-validation to identify the optimal lambda value (0.043) for constructing the model. The model showed optimal performance with 8 genes (key- NETs). Thus, the LASSO algorithm reduced the 16 DE-NET genes to a core set of 8 key-NET genes: HAT1, SELP, H2AX, MTOR, CYBB, AZU1, H2AZ1, and H2AJ, in the discovery set. Subsequent validation demonstrated the key-NET gene signature’s ability to distinguish between sepsis non-survivors and survivors, with high predictive accuracy. The AUC values were 89.3% (95% CI = 78%- 100%, p-value < 0.001) in the discovery set (Figure 4 D) and 93.2% (95% CI = 85.5%-100%, p-value < 0.001) in validation set-1 (Figure 4 E).

### 3.4. Construction and validation of the NET score model

A NET score model was constructed based on the ssGSEA scores of 8 key-NET genes. In the discovery set, the box plot illustrates that the NET score significantly (p-value < 0.001) differentiates between sepsis non-survivors, sepsis survivors, and controls (Figure 5 A), demonstrating notable predictive accuracy in discriminating between sepsis survivors and sepsis nonsurvivors with an AUC of 92.1% (Figure 5 B). The discriminative power of the NET score model was confirmed in validation set-1, with an AUC of 89.6% (Figure 5 C-D) indicating that the NET score model possesses substantial prognostic value. Furthermore, individual expression levels of key-NET genes did not show consistent performance in discriminating survivors from non-survivors (Supplementary Figure 4 A-B). To further investigate the clinical relevance of these eight genes, their relative mRNA expression levels in the discovery and validation set-1 were calculated and displayed as box plots in Supplementary Figure 4 C. All eight genes show up-regulation in non-survivors compared to survivors, with statistically significant differences (p < 0.05) observed in seven out of the eight genes. These eight genes are subsequently analysed for their functional significance.

**Figure 5:**
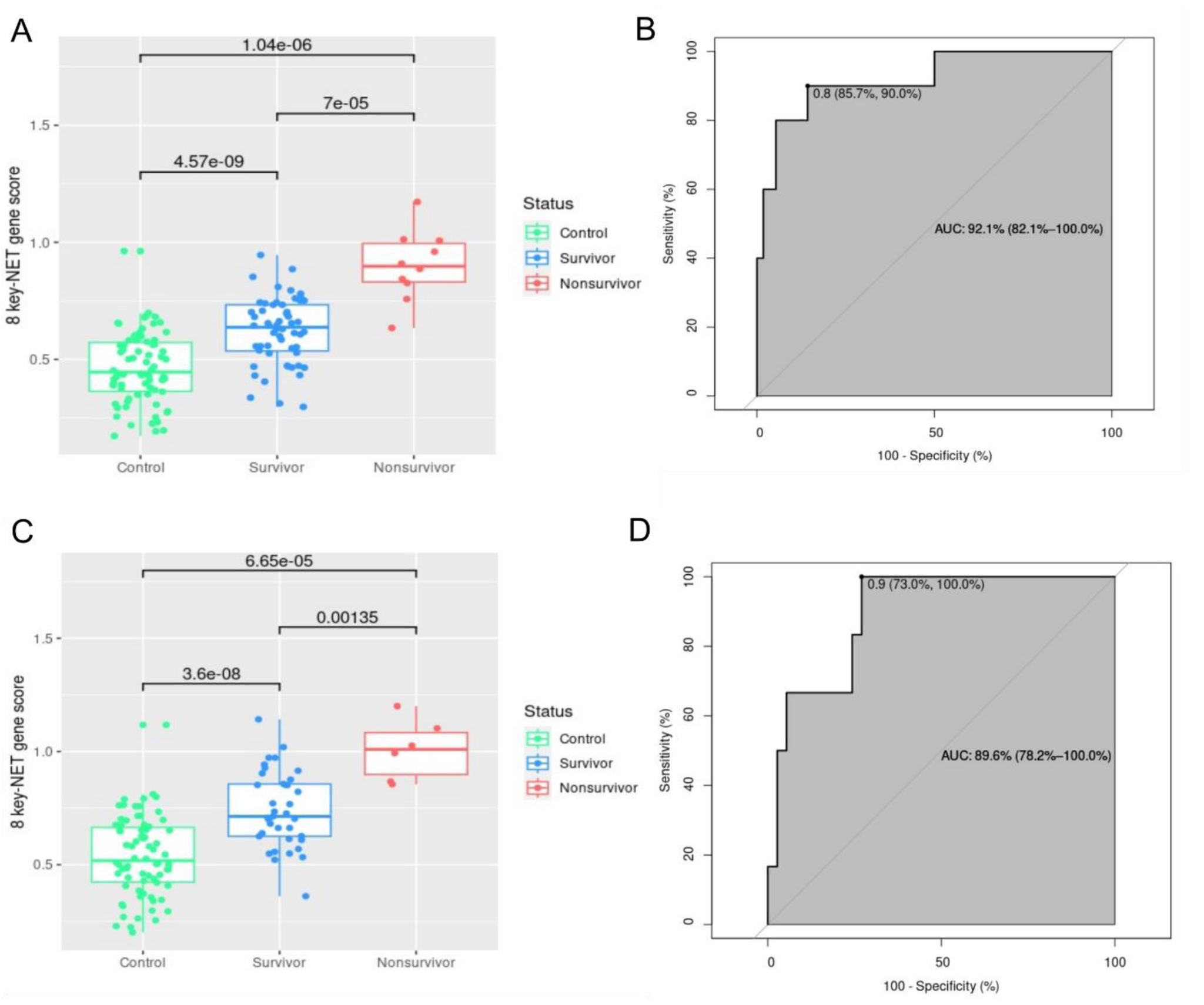
Construction and verification of NET score model using 8 key-NET genes. (A) Boxplot of the distribution of NET score model among the control (n=74), sepsis survivor (n=56) and sepsis nonsurvivor (n=10) groups within the discovery set. (B) ROC curve generated by the NET score model in discovery set stratifying sepsis survivor and sepsis nonsurvivor cases. (C) Boxplot of NET score among the control (n=74), sepsis survivor (n=37) and sepsis nonsurvivor (n=6) groups within validation set-1. (D) ROC curve generated by the NET score model in validation set-1 stratifying sepsis survivor and sepsis nonsurvivor cases.

### 3.5. Biological function analysis of DE-NET genes and Key-NET Genes

Comparing the functional enrichment pathways of biological processes (BP) enriched in DE- NET genes (Supplementary Table 6) and key-NET genes (Supplementary Table 7) revealed significant enrichment in regulation of platelet activation and regulation pathways. In view of the known biology of neutrophil-platelet-complement-coagulation interactions, we carried out further investigations into the complement-coagulation pathway.

### 3.6. Correlation Analysis of Key-NET and DE-CC Genes

Differentially Expressed Complement-Coagulation genes (DE-CC genes) were identified using a similar approach as for DE-NETs (Supplementary Figure 5 and Supplementary Table 8). Eight Complement-Coagulation genes including C1QC, C1QB, C1QA, SERPINB2, VSIG4, PROS1, PLAU and SERPINE2 were observed to be upregulated. The CC score was calculated using ssGSEA, employing a similar methodology as the NET score calculation. The NET score and the CC score exhibit significant positive correlations across both the discovery set, and validation set-1, showing high correlation in sepsis nonsurvivors (Figure 6 A-B). Individual gene expression of DE-CC genes and key-NET genes also shows significant positive correlation (Supplementary Figure 6).

**Figure 6:**
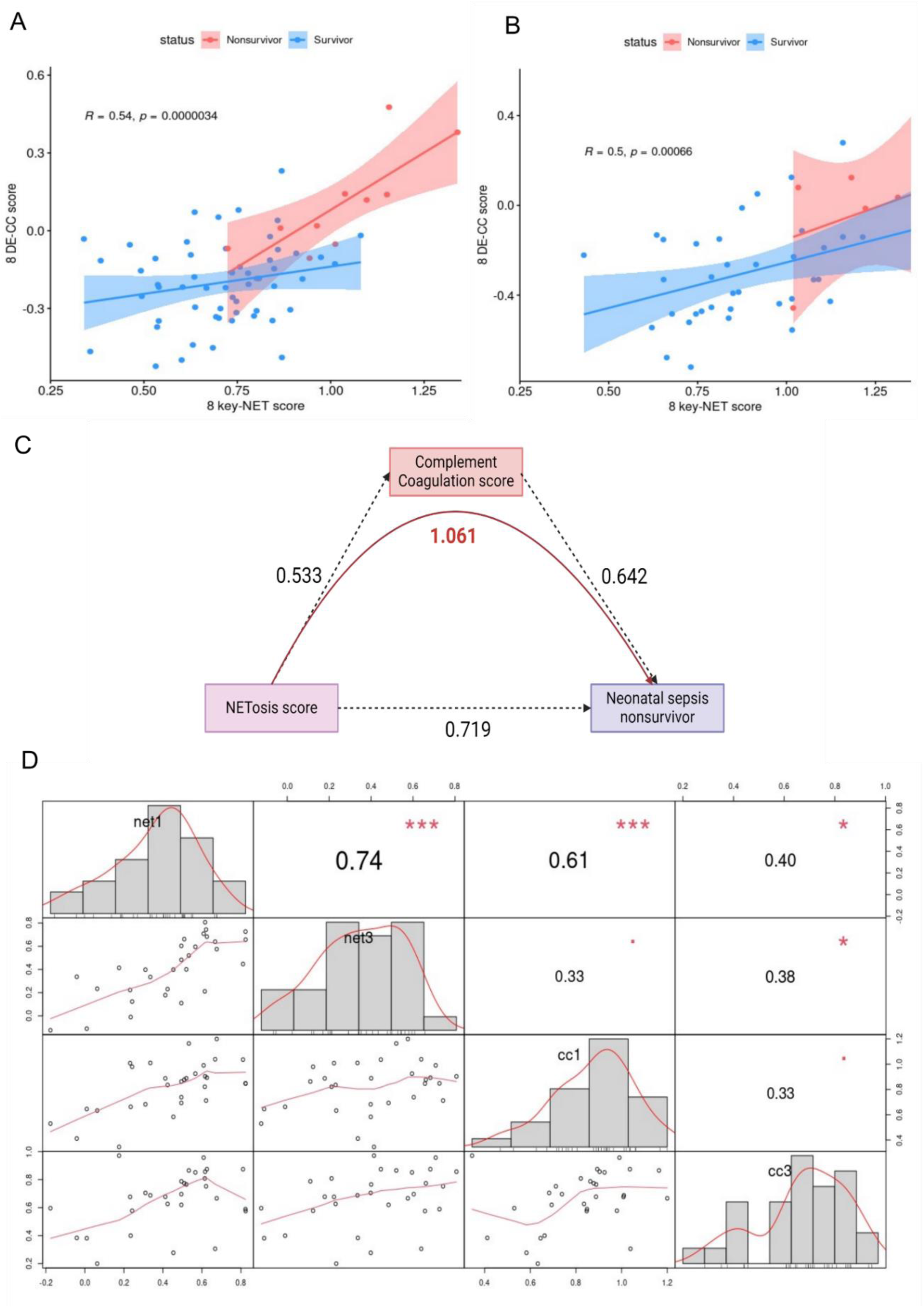
Correlation and temporal data validation of DE-NET genes and DE-CC genes. (A) Correlation of DE-NET genes and DE-CC genes in discovery set. (B) Correlation of DE-NET genes and DE-CC genes in validation set-1. (C) Mediation analysis (see Methods for details). (D) Correlation of DE-NET genes and DE-CC genes in independent temporal data (validation set-2) at two time points, Day 1 (n=30) and Day 3 (n=30). net1= NET score at day1, net3 = NET score at day3, cc1 = complement coagulation score at day 1, cc3 =

### 3.7. Mediation analysis between NET score and CC score

Mediation analysis revealed that the estimated coefficient of the direct effect of NETosis on sepsis outcome is 0.719, of coagulation on sepsis outcome is 0.642 and of NETosis on coagulation is 0.533. Whereas the estimated coefficient of indirect effect of NETosis acting through coagulation is 0.342, giving us a total effect of NETosis acting through coagulation on neonatal sepsis nonsurvivor to be 1.061 (0.719 + 0.342) (Figure 6 C). Thus, the influence of key-NET genes appears to be largely indirect, acting through DE-CC genes. This implies that key-NET expression precedes and triggers DE-CC expression. To corroborate these findings, validation set-2, containing data from two-time points - Day 1 and Day 3 – was analysed (Supplementary Table 2). A strong significant positive correlation was observed between the NET score at Day 1 and Day 3 as well as between the CC score at Day 1 and Day 3. Of note, the CC score at Day 1 and Day 3 were not correlated (Figure 6 D).

### 3.8. Construction of the Protein-Protein Interaction (PPI) Network and KEGG pathway

A PPI network was constructed for both DE-NET and key-NET genes with an interaction score of 0.4 (medium confidence). In this network, a node represents a protein, and an edge indicates an association between two proteins (either functional and/or physical), with the line thickness denoting the strength of data support. As depicted in Supplementary Figure 7 A, proteins encoded by the DE-NET genes are interconnected. Upon analysing the key-NET genes, the PPI network is segmented into two clusters and one node: the signalling molecules inside the neutrophil, the histone group and AZU1, the protein expelled out during NETosis process (Supplementary Figure 7 B). This is also evident from the KEGG pathways painted with appropriate colour-coding for DE- NET, key-NET and DE-CC genes (Supplementary Figure 8 A-B).

### 3.9. Immune cell deconvolution analysis

We conducted immune cell deconvolution analysis (Supplementary Table 9) to further investigate the differences in immune cell phenotypes between neonatal sepsis survivors and neonatal sepsis nonsurvivors (Supplementary Figure 9 A). Among the 22 immune cell types, neutrophils, T cells CD8 and dendritic cells activated shows significant downregulation in neonatal sepsis nonsurvivors whereas eosinophils show significant upregulation in neonatal sepsis nonsurvivors. Moreover, out of these four immune cell types (Neutrophils, T cells CD8, Dendritic cells and Eosinophils) only neutrophil shows positive correlation with key-NET genes consistently in both outcome groups (Supplementary Figure 9 B-C). This suggests that neutrophils are likely source of key-NET signature. Further, neutrophils are significantly depleted in neonatal sepsis nonsurvivors, strongly suggesting overwhelming suicidal NETois in this group. We have also checked correlation between the actual neutrophil count from clinical samples and neutrophil count generated from CIBERSORTx, showing significant positive correlation (p < 0.00001) (Supplementary Figure 10 A).

### 3.10. Association of NET score with pathogens

NET score was observed to be significantly associated with the presence of pathogen (Supplementary figure 10 B). Among the pathogen species detected, coagulase negative *Staphylococcus* was observed to be significantly associated with nonsurvival (p < 0.05) (Supplementary Figure 10 C).

### 3.11. Prognostic performance in sepsis from other age groups (Pediatric and Adult)

Supplementary Table 2 shows details about the datasets used. validation set-3 contains samples from pediatric sepsis patients from three datasets. No batch effect was observed between the three datasets (Supplementary Figure 11 A-C). NET score is higher in each of the sepsis groups – survivor (n=181) or nonsurvivor (n=47) - compared to control group (n=47) (Figure 7 A). However, NET score fails to significantly stratify sepsis nonsurvivors from sepsis survivor with an AUC of 60.8% (Figure 7 B). Similarly, LASSO model generated from neonatal sepsis patients shows poor performance in stratifying sepsis nonsurvivors form sepsis survivors with an AUC of 58.8% (Figure 7 C). Consistent with the above results individual gene expression of eight key-NET genes cannot distinguish between pediatric sepsis survivor from pediatric sepsis nonsurvivors, (Supplementary Figure 13 A).

**Figure 7:**
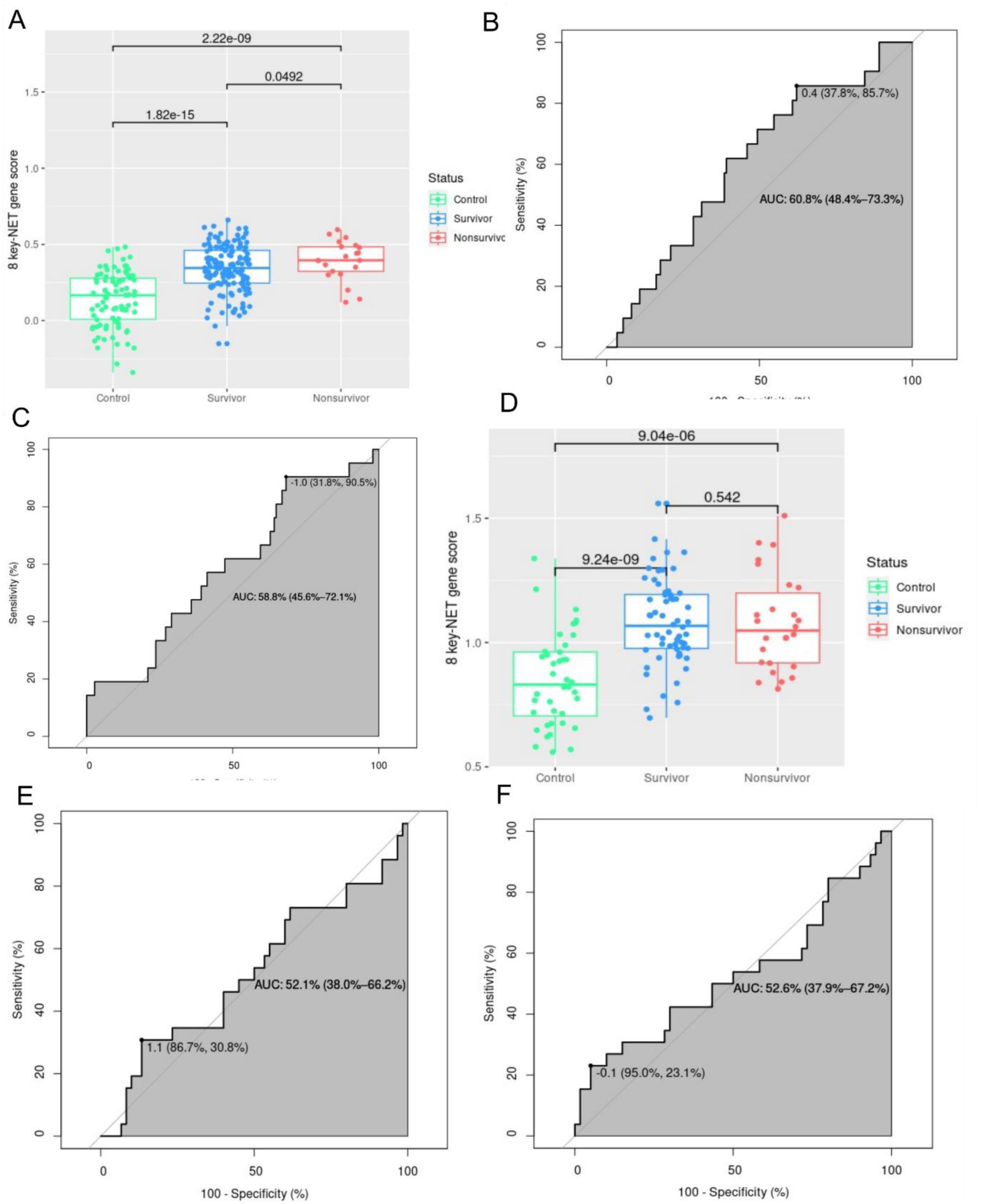
Evaluation of NET score and LASSO model developed from neonatal sepsis patients in other age groups (Pediatric sepsis: Validation set-3 and Adult sepsis: Validation set-4). (A) Boxplot of NET score in pediatric control (n=47), pediatric sepsis survivor (n=181) and pediatric sepsis nonsurvivor (n=27). Testing NET score in pediatric samples using (B) ssGSEA and (C) LASSO model generated from neonatal sepsis. (D) Boxplot of NET score in adult control (n=40), adult sepsis survivor (n=60) and adult sepsis nonsurvivor (n=26) (E) Testing NET score in adult sepsis using (E) ssGSEA, and (F) LASSO model generated from neonatal sepsis.

Validation set-4 contains sepsis samples from adult age group from two different datasets, batch effect correction was done as described earlier in neonatal sepsis cases (Supplementary Figure 12 A-F). NET score is significantly higher in sepsis groups - survivor (n=60) or nonsurvivor (n=26) – compared to the control group (n=40) (Figure 7 D) but does not discriminate between survivor and nonsurvivor, AUC = 52.1% (Figure 7 E). Similarly, LASSO model generated from neonatal sepsis does not stratify sepsis nonsurvivors form sepsis survivors with an AUC of 52.6% (Figure 7 F). Only H2AJ and CYBB are significantly upregulated in non-survivors of adult sepsis (Supplementary figure 13 B).

### 3.12. Validation of key-NET genes in clinical neonatal sepsis patients

To translate the potential value of key-NET genes into a clinical setting, we have used validation set-5 which contains neonates with sepsis (n=11) and neonates without sepsis (n=2). Details of their clinical records are provided in Supplementary Table 3 and batch effect was removed as described earlier (Supplementary Figure 14 A-F). Firstly, we observed an increasing trend of NET score in sepsis compared to control (Supplementary Figure 15). Further, we observed that except HAT1 and CYBB all other key-NET genes (Figure 8 A) tend to be upregulated in neonatal sepsis nonsurvivor (n=3) group as compared to neonatal sepsis survivor group (n=7) although none of them could reach the significance level (p < 0.05). When tested for NET score and CC score relation, a significant correlation of 0.8 (p < 0.005) was observed (Figure 8 B).

**Figure 8:**
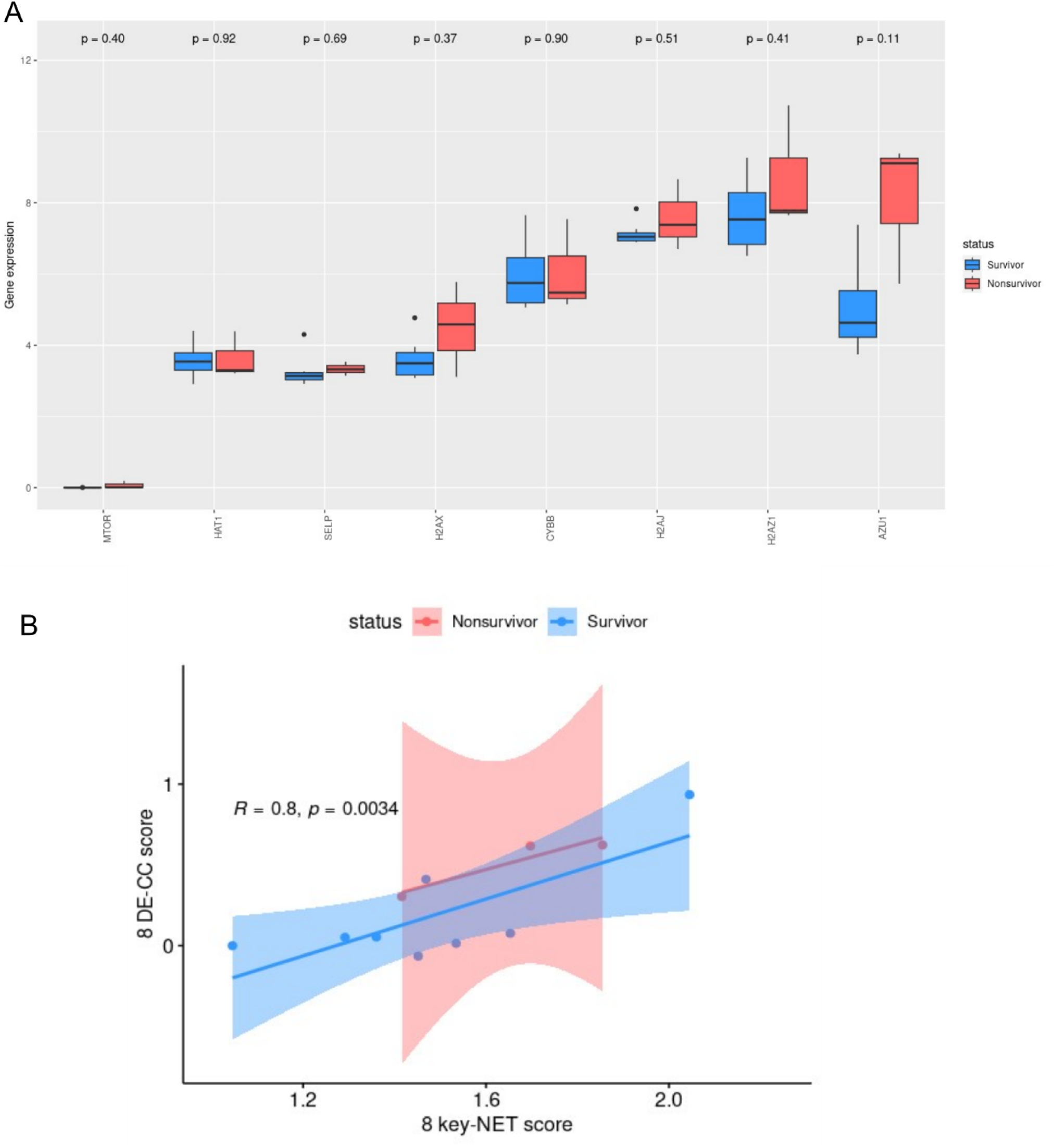
Validation of key-NET genes in clinical samples of neonatal sepsis. (A) Individual gene expression of eight key-NET genes in neonatal sepsis survivor (n=7) and neonatal sepsis nonsurvivor (n=3). (B) Correlation of NET score and CC score generated by using ssGSEA algorithm.

## 4 Discussion

Neonatal sepsis is a dysregulated host response caused by various pathogenic bacteria and infectious factors. Regarding the pathogenesis of sepsis, newborns are not small children(36). It has been reported that in pediatric septic shock, developmental ages display obvious effects on the host transcriptomic response to sepsis(37). This highlights the necessity for age-specific research into the development of sepsis to improve outcomes for neonates. However, such research presents unique challenges, particularly due to the limitations in blood volume available from newborns.

Pathogens trigger formation of intravascular reticular chromatin-based structures derived from neutrophils known as Neutrophil Extracellular Traps (NETs)(11). These structures are released into the extracellular environment to help eliminate infections. However, NETosis is also linked to hyperinflammatory responses that worsen tissue damage, boost autoimmunity and contribute to vascular thrombosis(38). Beyond their direct effects on neutrophils, pathogens activate TLR-4 receptors on platelets in sepsis, leading to platelet-neutrophil interaction, NET formation and enhanced bacterial trapping within blood vessels(39).

The levels of neutrophil-derived circulating free DNA (cf-DNA/NETs) have been shown to directly correlate with multiple organ dysfunction score, sepsis-related organ failure assessment, leukocyte counts, and MPO levels(40). Furthermore, MPO-DNA complex (plasma myeloperoxidase-conjugated DNA) has been identified as a predictor of outcomes and organ dysfunction in septic shock patients(41).

A preclinical model of neonatal sepsis demonstrated that the timely use of NET inhibitors improved survival(42). Thus, NET-targeting strategies may offer promising new avenues for risk stratification of neonatal sepsis based on organ dysfunction. This approach is particularly relevant given the strong association between abnormal coagulation and the high mortality rates observed in neonatal sepsis(43,44).

Big data, machine learning, and artificial intelligence together present a significant opportunity to develop more effective tools for risk stratification in neonatal sepsis. In this study, we conducted a comprehensive analysis of transcriptomic data from 120 neonates with sepsis and employed the LASSO (Least Absolute Shrinkage and Selection Operator) machine learning method to make a substantial advancement in the prognostication of neonatal sepsis. This analysis identified an 8-gene signature associated with NETosis, which forms the basis of a predictive model.

The model’s ability to accurately differentiate between survivors and nonsurvivors of sepsis underscores its potential as a screening tool for risk stratification in neonatal sepsis cases. Notably, this score is significantly high in neonatal sepsis nonsurvivors compared to neonatal sepsis survivors, highlighting its potential utility in clinical practice.

The Gene Ontology (GO) enrichment analysis for Biological Processes of key-NET genes revealed that platelet activation and regulation are consistently enriched among differentially expressed NETosis-related (DE-NET) genes, indicating their critical role in neonatal sepsis mortality. A close relationship was identified between the NET score and the complement-coagulation (CC) score when these were correlated in both the discovery and validation sets.

Further mediation analysis demonstrated that the expression of NETosis genes (key-NET) in the blood precedes the abnormal activation of genes involved in intravascular complement and coagulation cascades (DE-CC). This finding was substantiated by temporal data collected at two time points—Day 1 and Day 3—where a strong, significant positive correlation was observed between the NET score at Day 1 and Day 3, as well as between the NET score and the CC score at both time points. Interestingly, no significant correlation was found between the CC scores at Day 1 and Day 3, suggesting that NETosis may be the driving force behind the activation of complement-coagulation genes in neonatal sepsis cases.

A Protein-Protein Interaction (PPI) analysis was conducted to explore the relationships among all sixteen differentially expressed NETosis-related (DE-NET) genes and the key-NET genes. Our analysis revealed that the key-NET genes form three distinct clusters: one cluster comprises signaling genes (SELP, CYBB, and MTOR); another includes histone-related genes (HAT1, H2AX, H2AZ1, and H2AJ); while AZU1 appears as a single, unconnected node (Supplementary Figure 7A). It is noteworthy that histone proteins are well-documented triggers of platelet aggregation and intravascular coagulation(45).

When the fractions of immune cells in both survivors and nonsurvivors were correlated with the respective expression of key-NET genes, we observed that dendritic cells (activated), CD8 T cells, and eosinophils exhibited a significant negative correlation, whereas neutrophils showed a significant positive correlation with most of the eight key-NET genes. Moreover, the correlation between neutrophil fraction and key-NET gene expression was notably stronger in neonatal sepsis nonsurvivors compared to survivors. This finding supports our hypothesis that neutrophils in nonsurvivors are undergoing NETosis to a significantly greater extent compared to the survivors. The positive correlation between neutrophil count and key-NET gene expression further suggests that neutrophils are the primary blood cell type expressing these key-NET genes, reinforcing their central role in the pathogenesis of neonatal sepsis.

We observed that the NET score is elevated in neonatal sepsis cases where a pathogen is detected, consistent with the role of neutrophils in pathogen clearance. As neutrophils perform their function and undergo NETosis, the expression of key-NET genes increases, leading to a higher NET score in samples with detectable pathogens. When we stratified the data based on specific pathogens, we observed that coagulase-negative *Staphylococcus* was associated with a significantly higher NET score in nonsurvivors compared to survivors. This aligns with previous reports on the causal role of coagulase negative *staphylococcus* in neonatal sepsis(46).

Our model not only predicts outcomes but also sheds light on the mechanisms underlying morbidity and mortality. The explanatory power of our model reveals the complex interactions between NETosis and the pathophysiological trajectory of sepsis. However, when the model was tested on other age groups, including pediatric and adult sepsis, it demonstrated poor performance (Supplementary Figure 13). Given the well-documented unique transcriptomic response to sepsis across different age groups(47), it is expected that this prognostic model is specifically tailored to the neonatal age group.

In our study, we identified eight key genes (Supplementary Table 10) involved in the formation and regulation of neutrophil extracellular traps (NETs), which highlight potential therapeutic targets and their relationship to coagulation and complement processes (Figure 9). Among these, Plasma soluble P-selectin (sP-selectin) is derived from these two cellular sources- endothelial cells and platelets(48,49).Previous studies have established a strong positive correlation between coagulation markers—such as disseminated intravascular coagulation, fibrinogen consumption, and thrombin activation—and sP-selectin levels in septic patients, consistent with platelet shedding of P-selectin(50). Our data similarly shows increased expression of the SELP gene in the whole blood transcriptome, indicating that platelet shedding is a significant contributor to sP-selectin levels.

**Figure 9:**
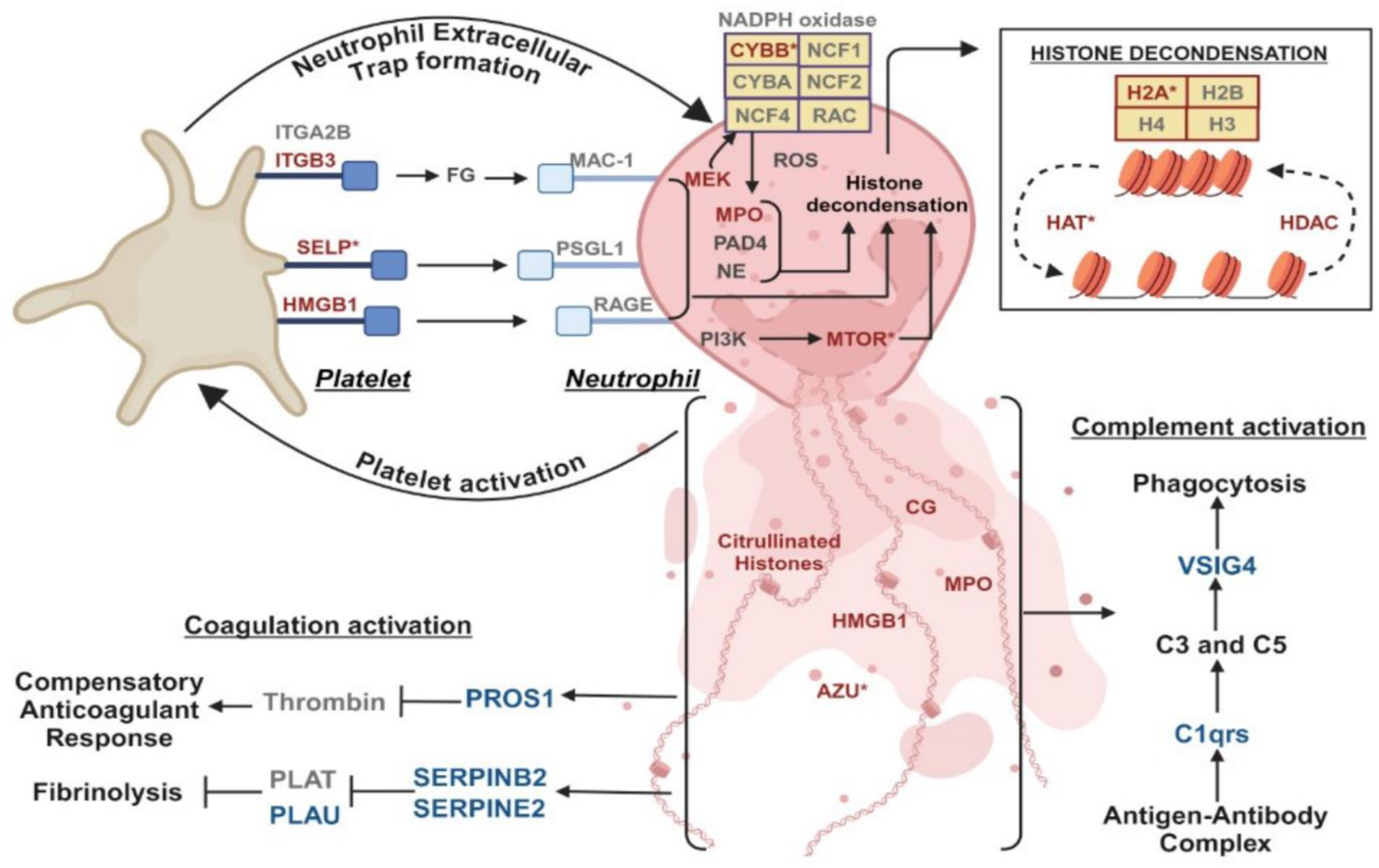
Proposed model of neutrophil-platelet interaction in neonatal sepsis. Where, genes in red are DE-NET genes, genes in red and * are key-NET genes, genes in blue are DE-CC genes and genes in grey are other genes involved in the interaction. Detailed annotation of the highlighted genes included in this model are listed in Supplementary Table 10.

Regardless of its origin, sP-selectin can negatively modulate direct leukocyte-endothelial interactions and/or indirect platelet-mediated secondary capture of leukocytes on the endothelium, both processes being dependent on the interaction between P-selectin and its primary ligand, platelet selectin glycoprotein ligand-1 (PSGL-1) (49). PSGL-1 primes neutrophils for NETosis(51), making it a promising therapeutic target for modulating inflammatory response.

Another key gene identified is CYBB, also known as NOX2, which encodes the gp91^phox^ subunit of the NADPH oxidase complex. NETosis is dependent on reactive oxygen species (ROS) such as superoxide, generated by NADPH oxidase 2 (NOX2)(52,53). Extensive studies on sepsis have shown that levels of soluble NOX2-derived peptides are significantly elevated in sepsis and septic shock patients compared to healthy controls. Additionally, sepsis nonsurvivors tend to exhibit higher levels of these peptides compared to survivors(54,55). Consistent with these findings, we observed elevated levels of CYBB in our adult sepsis cases, with significant differences between survivors and nonsurvivors.

The mechanistic target of rapamycin (MTOR) is another key gene identified in our study. MTOR, a highly conserved PI3K-like serine/threonine kinase(56), regulates NET formation through the posttranscriptional control of hypoxia-inducible factor 1α (HIF-1α), a crucial modulator of antimicrobial defense(57). Elevated serum levels of phosphorylated ribosome S6 protein kinase (PS6K), indicative of abnormal mTOR pathway activation, have been identified as an independent risk factor for 28-day mortality in sepsis patients(58).

Azurocidin 1 (AZU1), also known as heparin-binding protein (HBP), is another key gene. AZU1 is released early in response to bacterial antigens and is found in azurophilic granules and secretory vesicles of neutrophils(59,60). HBP is a promising serum biomarker for septic patients in intensive care units (ICUs)(61,62).

Histone acetyltransferase 1 (HAT1), a type B histone acetyltransferase (HAT1), is also among the genes identified. HAT1 belongs to the Gen5-related N-acetyltransferase (GNAT) family and is involved in the posttranscriptional modification of lysine residues on histones, which weakens the binding of negatively charged DNA, leading to an open-chromatin structure and active transcription. The balance between HATs and histone deacetylases (HDACs), which trigger a compact nucleosome structure preventing active transcription, is crucial for gene expression regulation during immune responses and NET formation(63).

Finally, circulating histones, such as H2AZ1, H2AX, and H2AJ, which are mainly derived from neutrophil extracellular traps, play a critical role in sepsis by mediating inflammatory responses, organ injury, and death(64).

Prognostic models for neonatal sepsis have been reported previously(65–69), primarily relying on clinical and routine laboratory parameters. In contrast, we aimed to develop a model with greater explanatory power by focusing on a crucial dysregulated pathway. First, we constructed a prognostic model for neonatal sepsis centred on the neutrophil extracellular trap (NET) formation pathway, resulting in higher AUC values. Second, our eight key-NET gene signatures demonstrated a trend of higher expression in sepsis nonsurvivors compared to survivors in the clinical samples collected. Third, the model facilitates the ’rapid’ prognosis of neonatal sepsis, with results potentially available in a few hours after the first suspicion of sepsis in the neonate. Lastly, we examined the relationship between the NET score and the CC score, confirming that coagulopathy is downstream of and likely triggered by increased formation of NETs in the intravascular space.

Our research has a few limitations. First, our study is based on data from 120 cases of neonatal sepsis. We conducted a comprehensive search and utilized all available neonatal sepsis data with outcome information in the NCBI GEO database to date. Additionally, we generated data from a small, well-defined cohort for which detailed clinical data were collected, ensuring that the transcriptional signature is suitable for prognostic stratification. Second, we did not differentiate between sepsis and septic shock in our study; septic shock cases were included in the broader category of sepsis for data analysis. This approach was primarily taken because our objective was not to diagnose sepsis per se but to detect signals of rapid deterioration and non-survival. We corrected for batch effects in data from cohorts that included both sepsis and septic shock patients, which we believe makes our signature appropriate for predicting outcomes in neonates with sepsis, regardless of the presence of shock.

## 5 Conclusions

In conclusion, our study unveils a novel 8 gene prognostic model for neonatal sepsis. This model not only accurately predicts outcome but also offers significant insights into the underlying mechanisms of sepsis-related morbidity and mortality. By linking NETosis with the complement-coagulation cascade, our findings highlight the critical role of neutrophils in the pathophysiology of neonatal sepsis and suggest potential therapeutic targets for improving outcomes. Despite the study’s limitations, including a small cohort size and the inclusion of septic shock within the broader sepsis category, our results provide a foundation for age-specific, rapid prognostication in neonatal sepsis. This work paves the way for future research that could transform our approach to neonatal sepsis, offering hope for improved survival and reduced morbidity in this vulnerable population.

## Data Availability

All data produced in the present study are available upon reasonable request to the authors.

## 6 List of abbreviations

AUC: Area Under the ROC Curve
AZU1: Azurocidin 1
BP: Biological Processes
cf-DNA: Circulating free DNA
CC: Complement-Coagulation
CSIR: Council of Scientific and Industrial Research
DAMP: Damage-Associated Molecular Pattern
DE-CC: Differentially Expressed Complement-Coagulation genes
DE-NET: Differentially Expressed NETosis Genes
GNAT: Gen5-related N-acetyltransferase
GO: Gene Ontology
GSEA: Gene Set Enrichment Analysis
HBP: Heparin-Binding Protein
HAT1: Histone Acetyltransferase 1
HDAC: Histone Deacetylase
HIF-1α: Hypoxia-Inducible Factor 1α
ICU: Intensive Care Unit
KEGG: Kyoto Encyclopedia of Genes and Genomes
LASSO: Least Absolute Shrinkage and Selection Operator
MTOR: Mechanistic Target of Rapamycin
MPO-DNA: Myeloperoxidase-conjugated
DNA NOX2: NADPH oxidase 2
NET: Neutrophil Extracellular Trap
PS6K: Phosphorylated ribosome S6 protein Kinase
PSGL-1: Platelet Selectin Glycoprotein Ligand-1
PPI: Protein-Protein Interaction network
ROS: Reactive Oxygen Species
ROC: Receiver Operating Characteristic
IRB: Research Institutional Review Board
ssGSEA: single-sample Gene Set Enrichment Analysis
sP-selectin: soluble P-selectin

## 7 Ethics approval and consent to participate

The study was approved by the Office of Research Institutional Review Board (IRB) of Christian Medical College (Registration No: ECR/326/INST/TN/2013) and Institutional Ethics Committee of All India Institute of Medical Sciences Bhubaneswar (Registration No: ECR/534/Inst/OD/2014/RR-20). Written informed consent was obtained from the patients’ legal representatives.

## 8 Consent for publication

Not applicable.

## 9 Availability of data and materials

All data and custom code written in R software for this study are available in the Supplementary_data.zip.

## Acknowledgement

SKM acknowledges intramural support from BRIC-NIBMG for generation of part of the validation data. DS acknowledges Senior Research Fellowship received from Council of Scientific and Industrial Research (CSIR), Government of India.

## Supplementary Figures

**Supplementary Figure 1:**
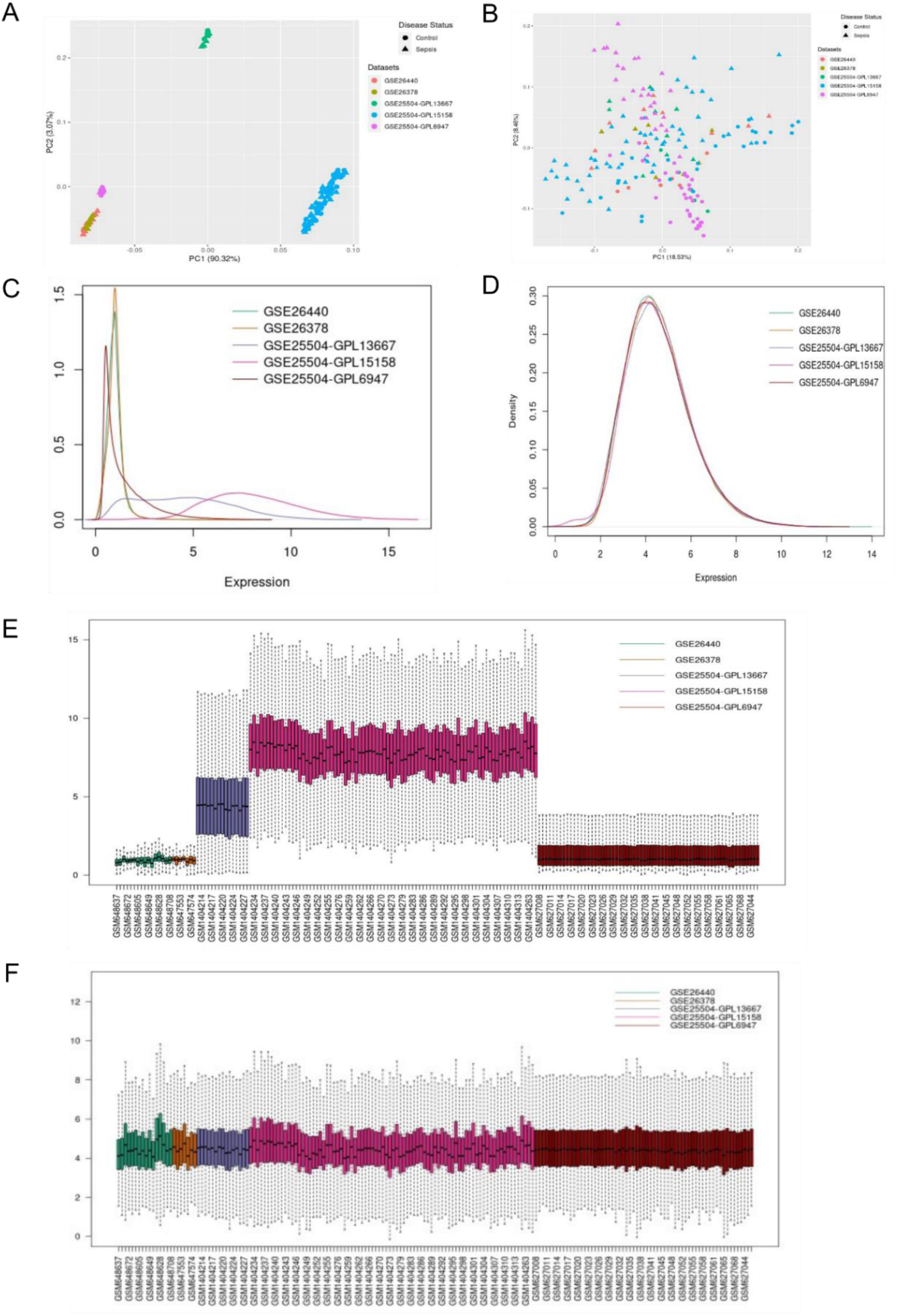
Batch effect removal. (A) PCA plot of the five datasets before batch effect correction. (B) PCA plot after batch effect correction. (C) Expression density plot of the five datasets before batch effect correction. (D) Expression density plot of the five datasets after batch effect correction. (E) Expression distribution plot of samples for

**Supplementary figure 2:**
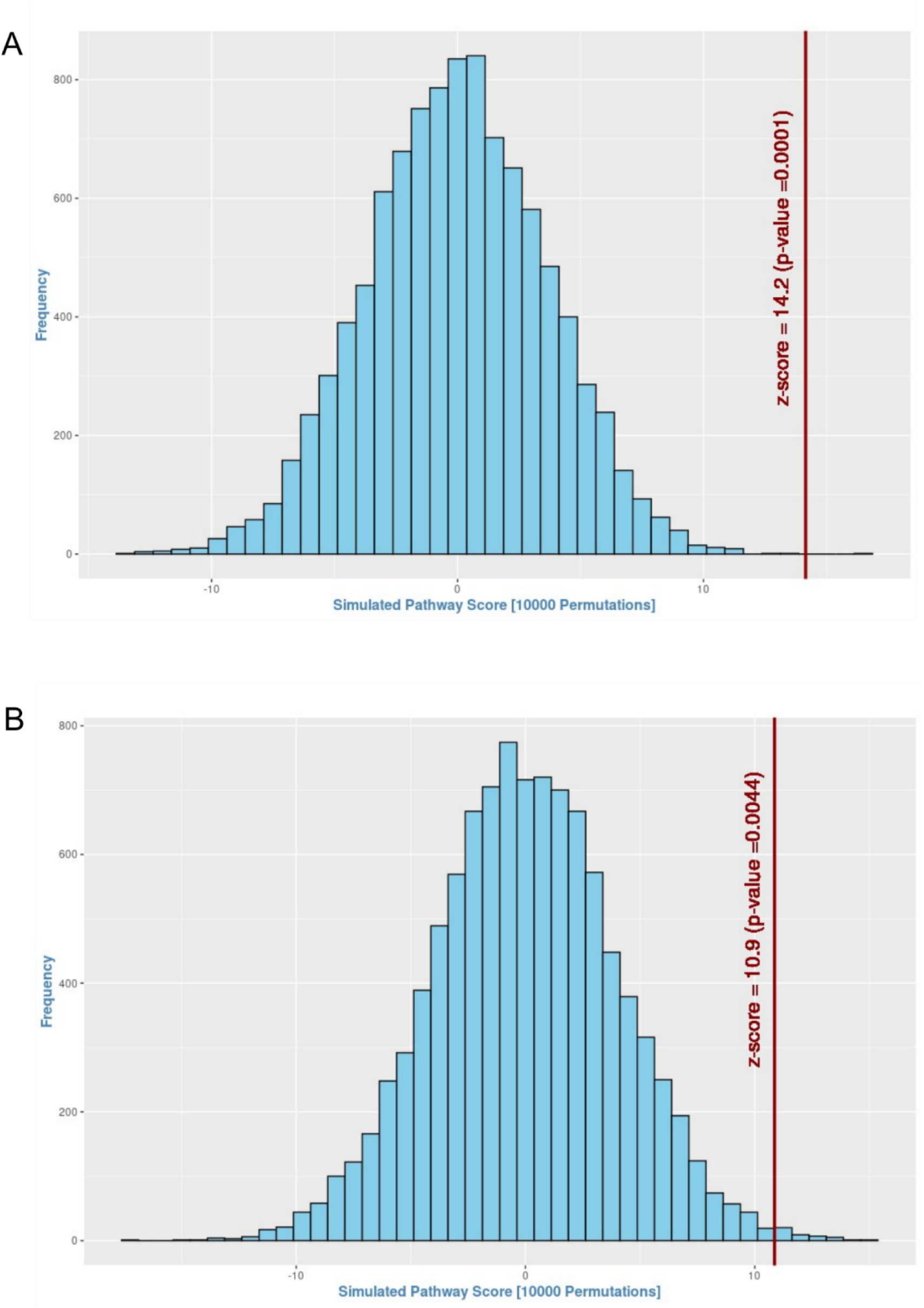
Upregulation of genes functional in NETosis in blood of patients with sepsis. Gene Set Enrichment Analysis (GSEA) was performed in (A) Discovery Set: between control group (n=74) and neonatal sepsis group (n=66); and (B) Validation Set-1: between control group (n=74) and neonatal sepsis group (n=43). The red vertical line on the extreme right suggests upregulation of the NETosis pathway. The magnitude (zscore) and the significant (p-value) of up-regulation are indicated beside the red vertical line. The histogram (blue bars) represents the null distribution of the pathway score calculated for each of the 10,000 iterations of permuting the sample labels.

**Supplementary figure 3:**
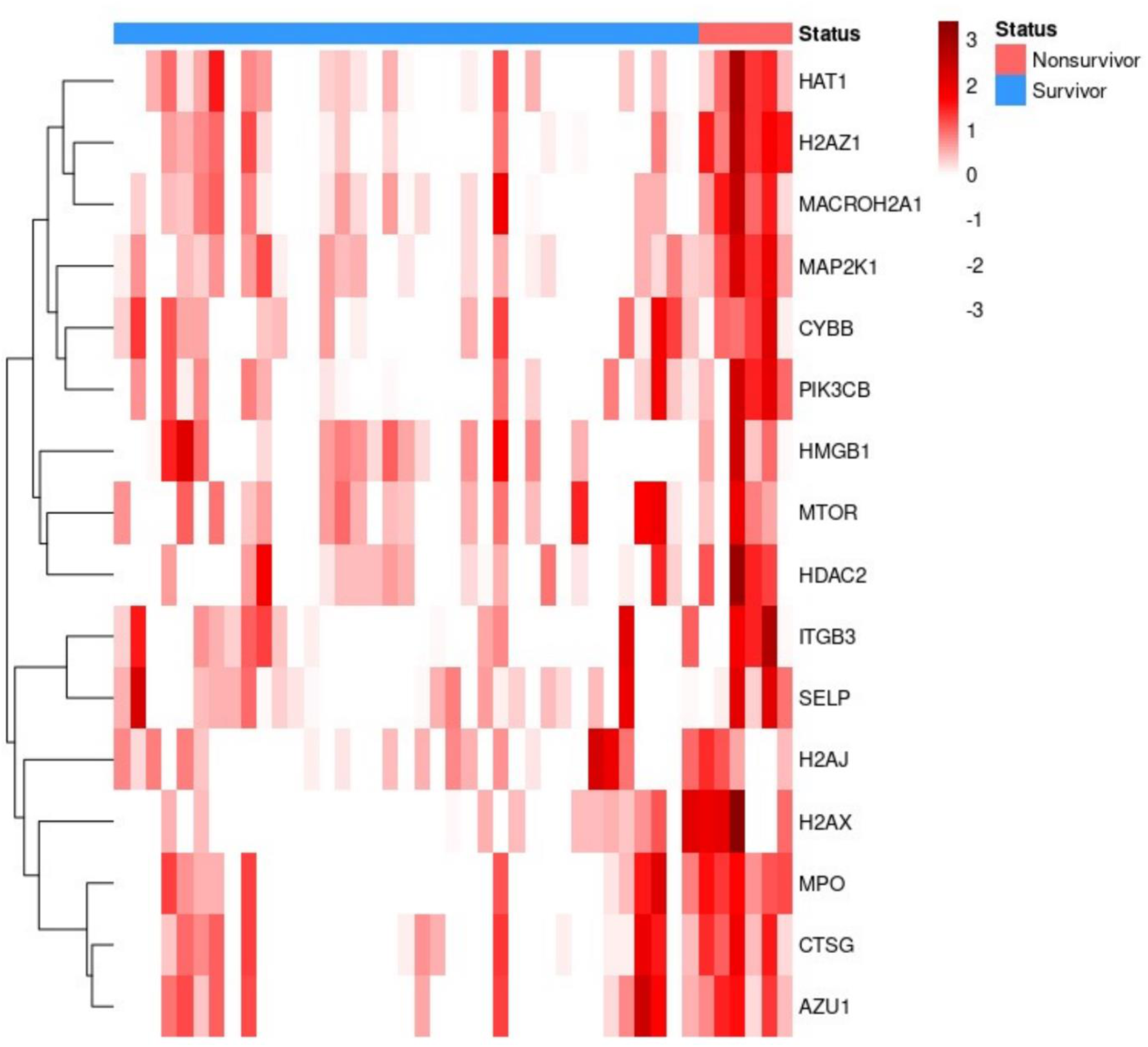
Expression of the 16 DE-NET genes in validation set-1. There is increased expression of DE-NET genes in the non-survivor group.

**Supplementary figure 4:**
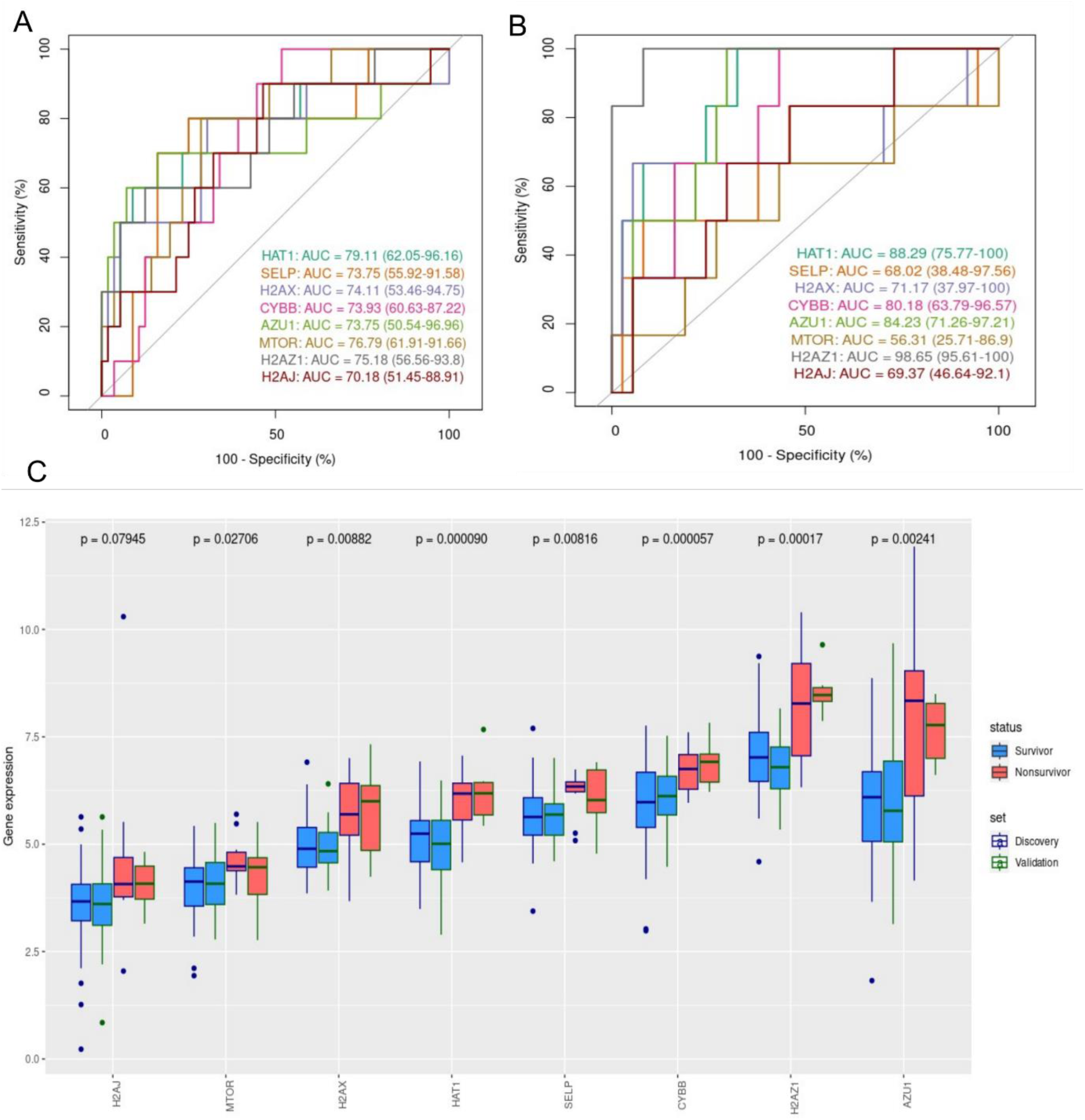
Generation of ROC curve and boxplots. (A) Individual gene (key-NET) ROC curve in discovery set. (B) Individual gene (key-NET) ROC curve in validation set-1. (C) Box plot showing comparison of individual (key-NET) gene expression in discovery and validation set-1.

**Supplementary figure 5:**
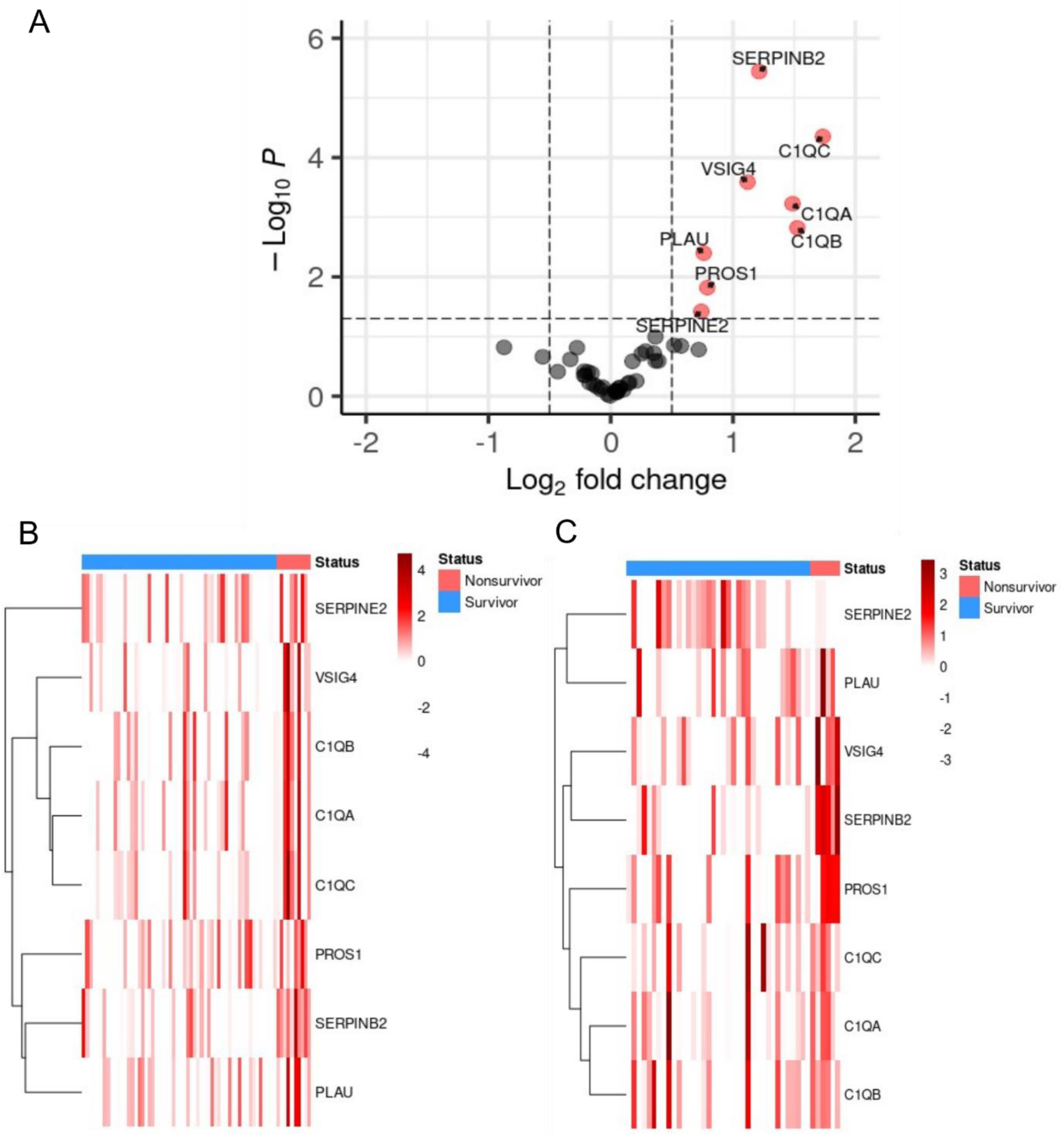
Identification and expression of differentially expressed complement coagulation genes (DE-CCs). (A) Volcano plot shows the eight DE-CC genes up-regulated in non-survivors compared to survivors (B) Expression of DE-CC genes in discovery set. (C) Expression of DE-CC genes in validation set-1.

**Supplementary figure 6:**
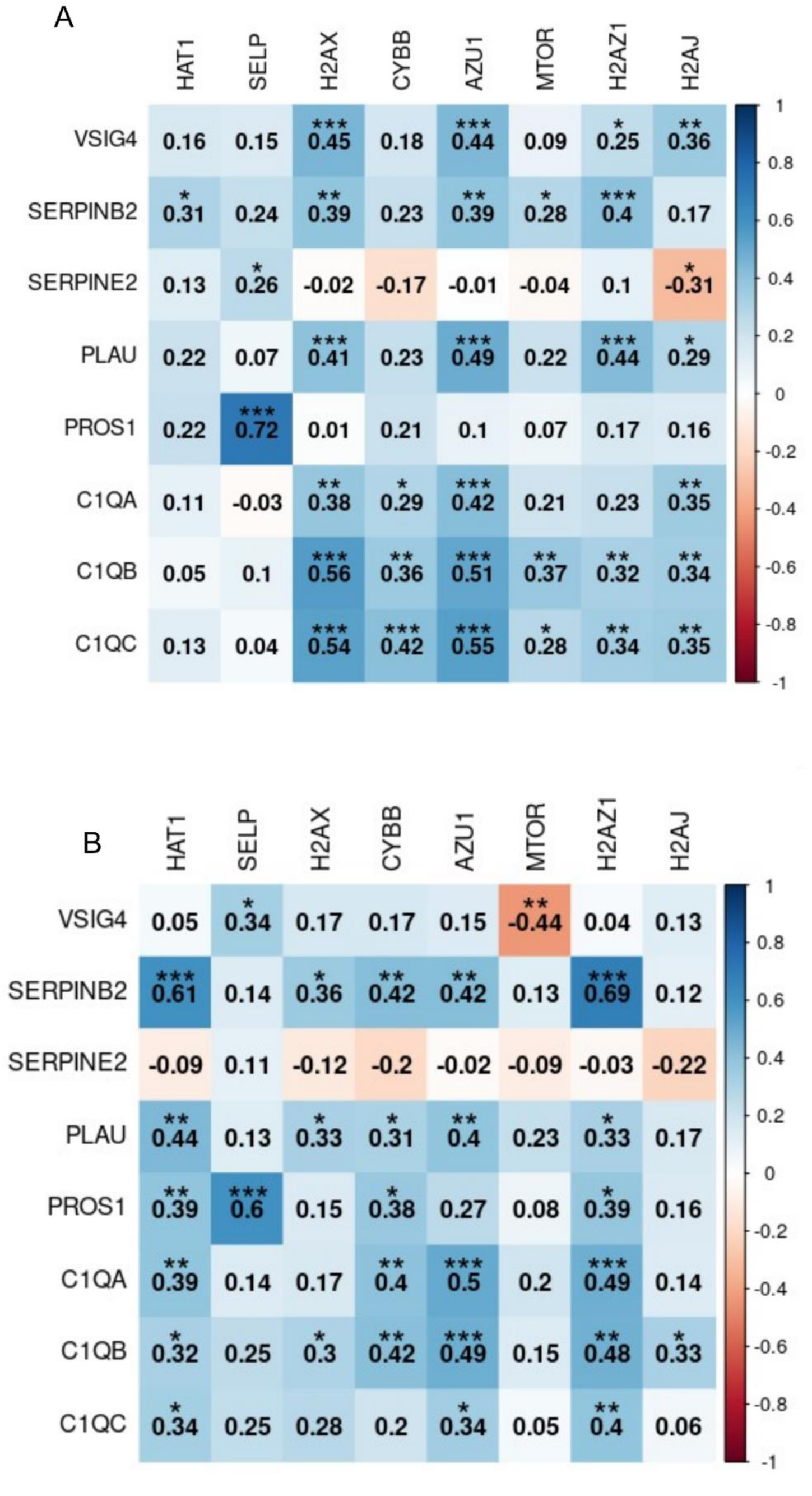
Correlation of key-NET gene expression and DE-CC gene expression. (A) Correlation in discovery set. (B) Correlation in validation set-1. (*** p value < 0.001, ** p value < 0.01, * p value <0.05)

**Supplementary figure 7:**
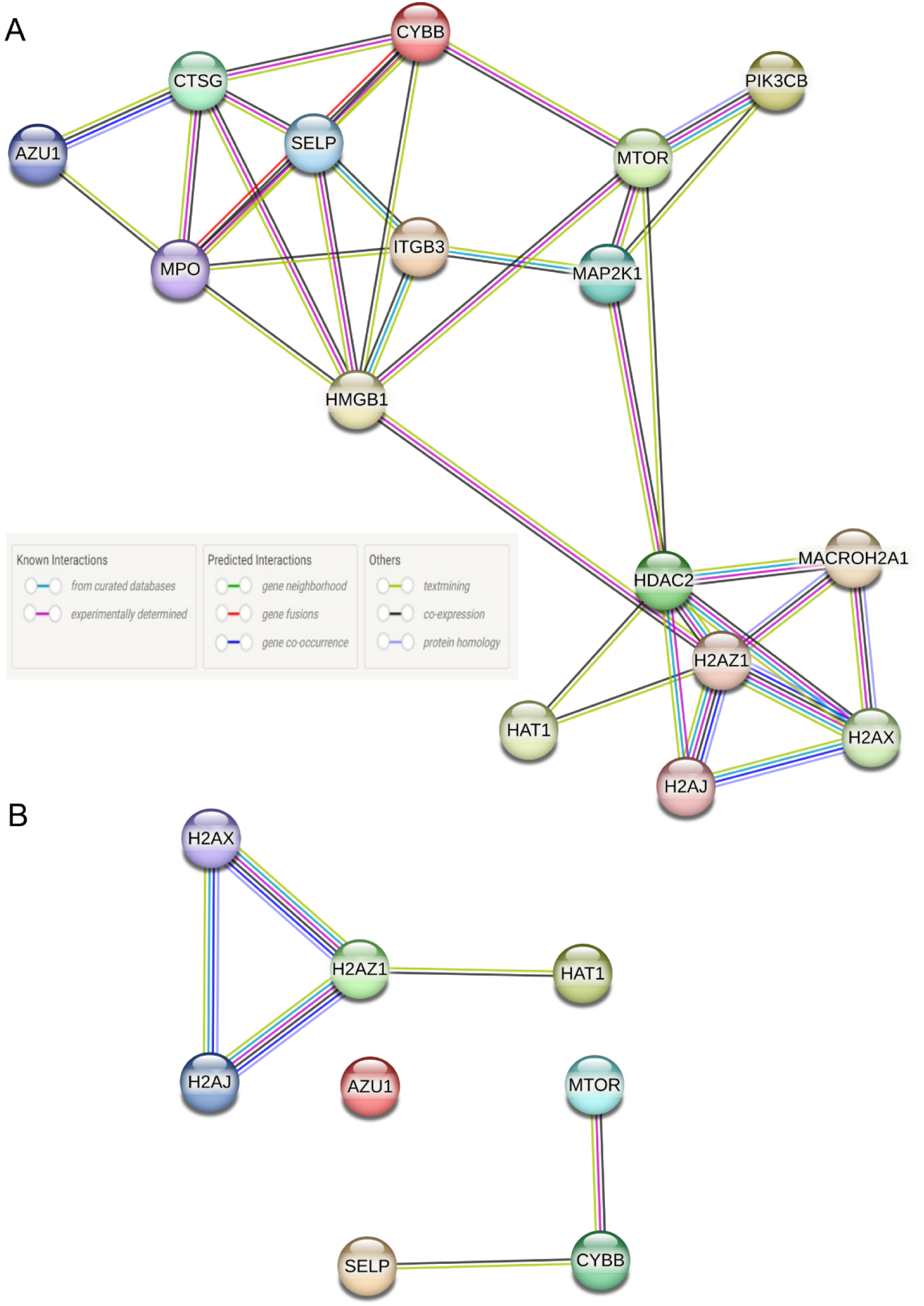
Construction of Protein-Protein Interaction network. (A) DE-NET genes (B) key-NET genes - the PPI network is segmented into two clusters and one node: the signalling molecules inside the neutrophil, the histone group and AZU1, the protein expelled out during NETosis process.

**Supplementary figure 8:**
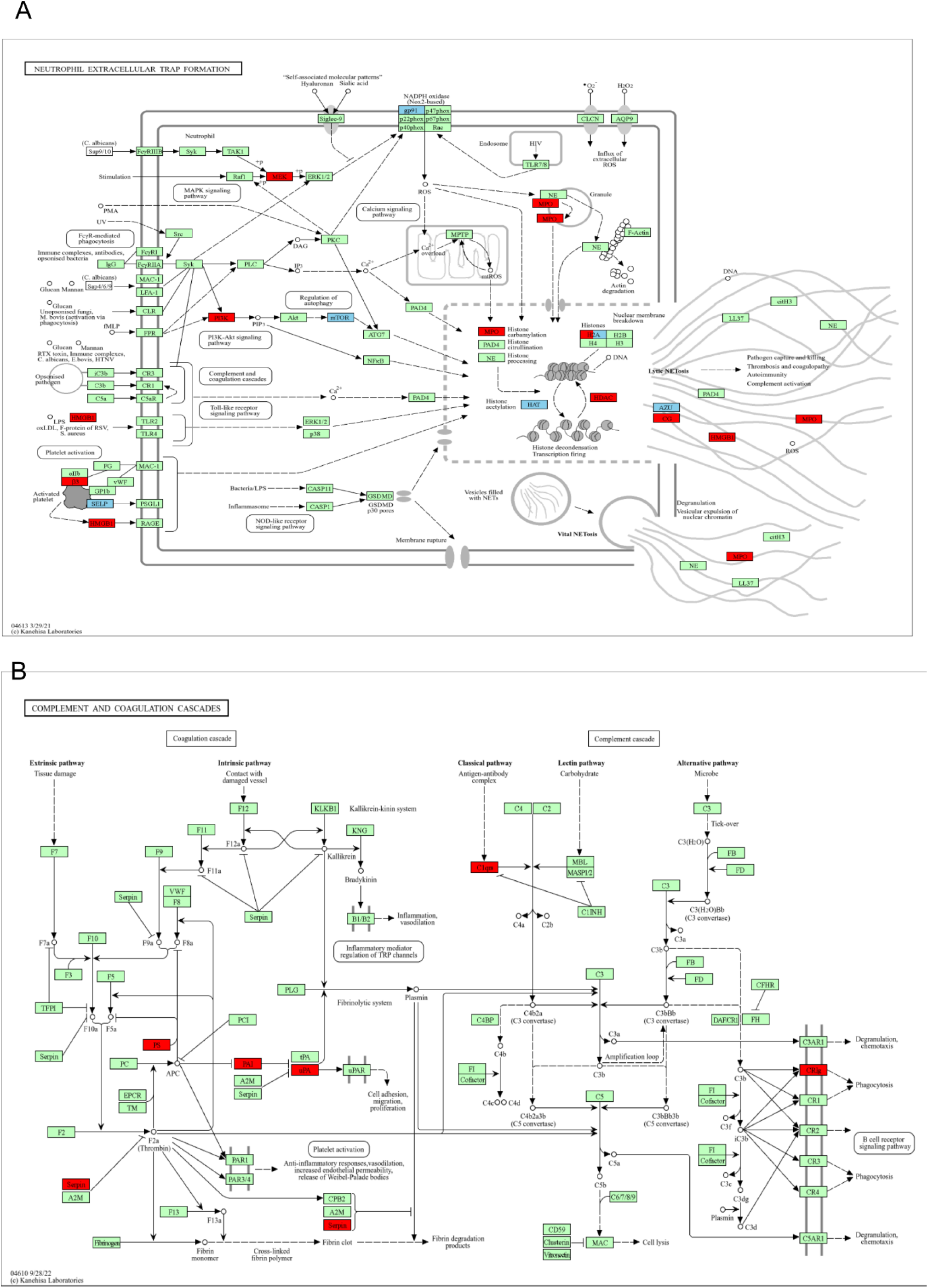
Painting the KEGG pathways. (A) DE-NET (red) and key-NET (blue) genes in neutrophil extracellular pathway formation (NETosis) – hsa04613. (B) DE-CC genes (red) in complement coagulation pathway – hsa04610.

**Supplementary figure 9:**
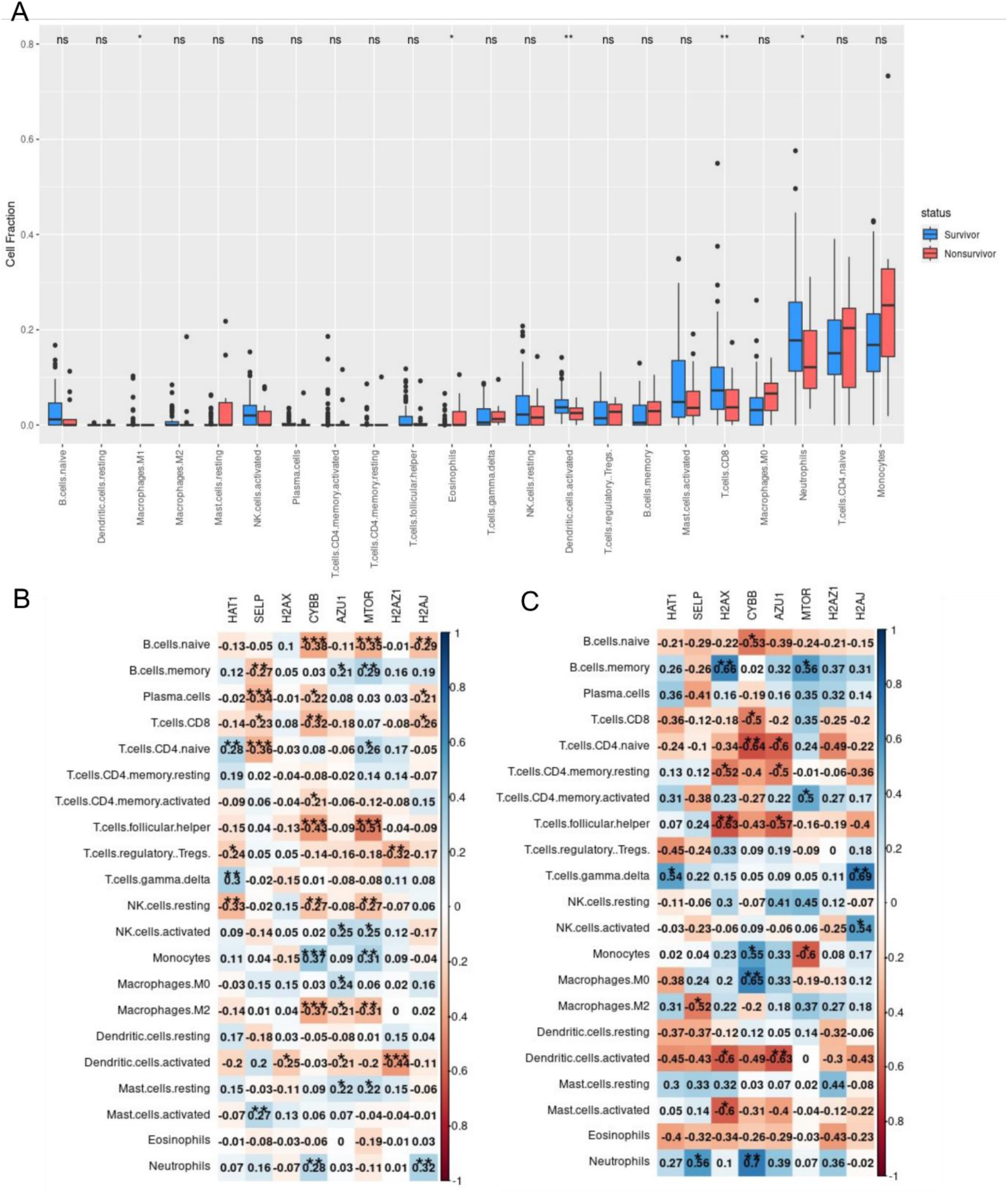
Immune cell deconvolution analysis in neonatal sepsis. (A) Abundance of 2 infiltrating immune cells between neonatal sepsis survivors (n=93) and nonsurvivors (n=16). (B) Correlation of immune cells with gene expression of key-NET genes in sepsis survivor group. (C) Correlation of immune cells with gene expression of key-NET genes in sepsis nonsurvivor group.

**Supplementary figure 10:**
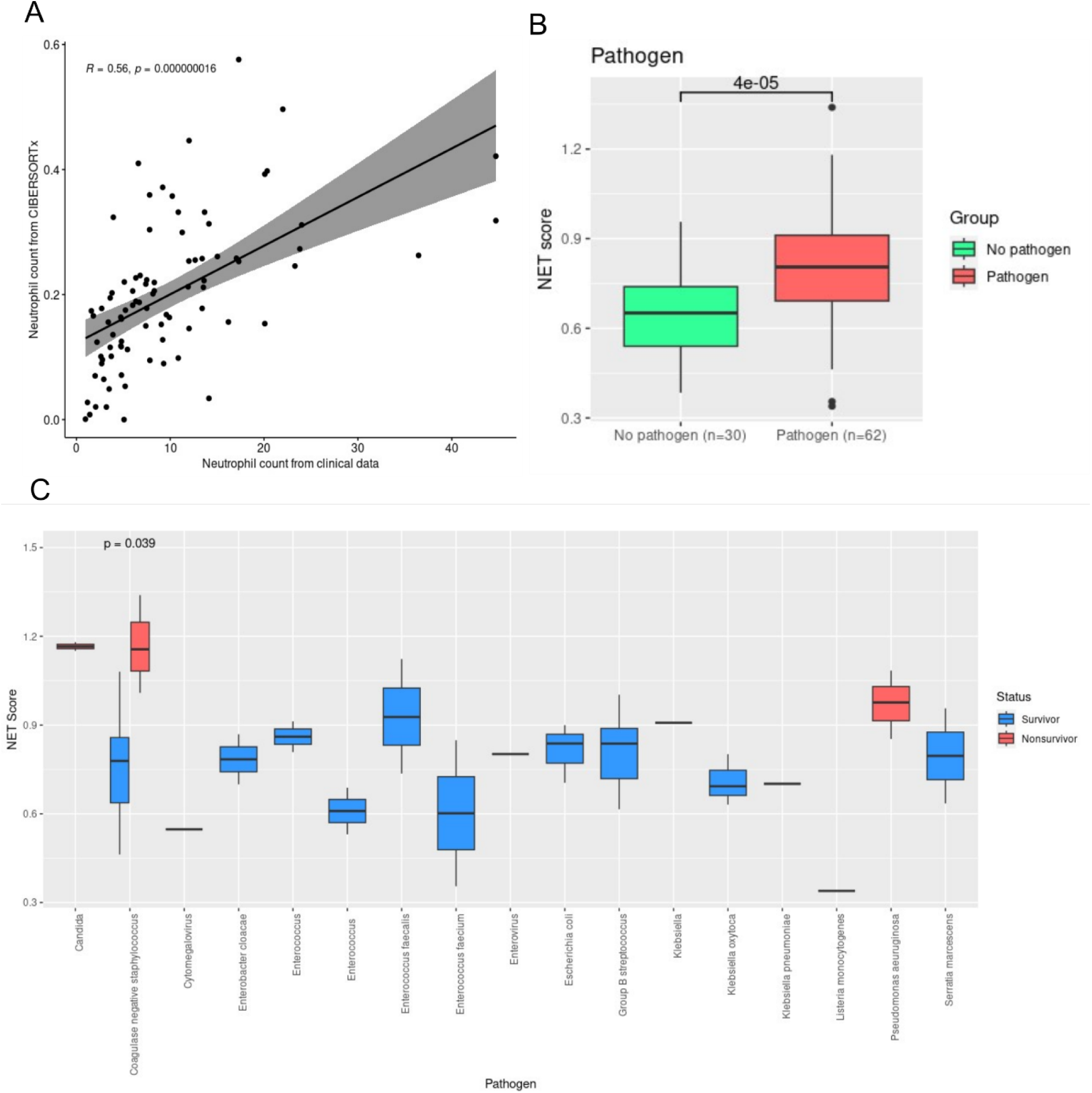
Clinical data analysis. (A) Correlation of neutrophil count from clinical samples and neutrophil count obtained from CIBERSORTx results. (B) Box plots show higher NET score in samples with detected pathogens (C) NET score is significantly higher in neonates infected by coagulase negative *Staphylococcus*.

**Supplementary figure 11:**
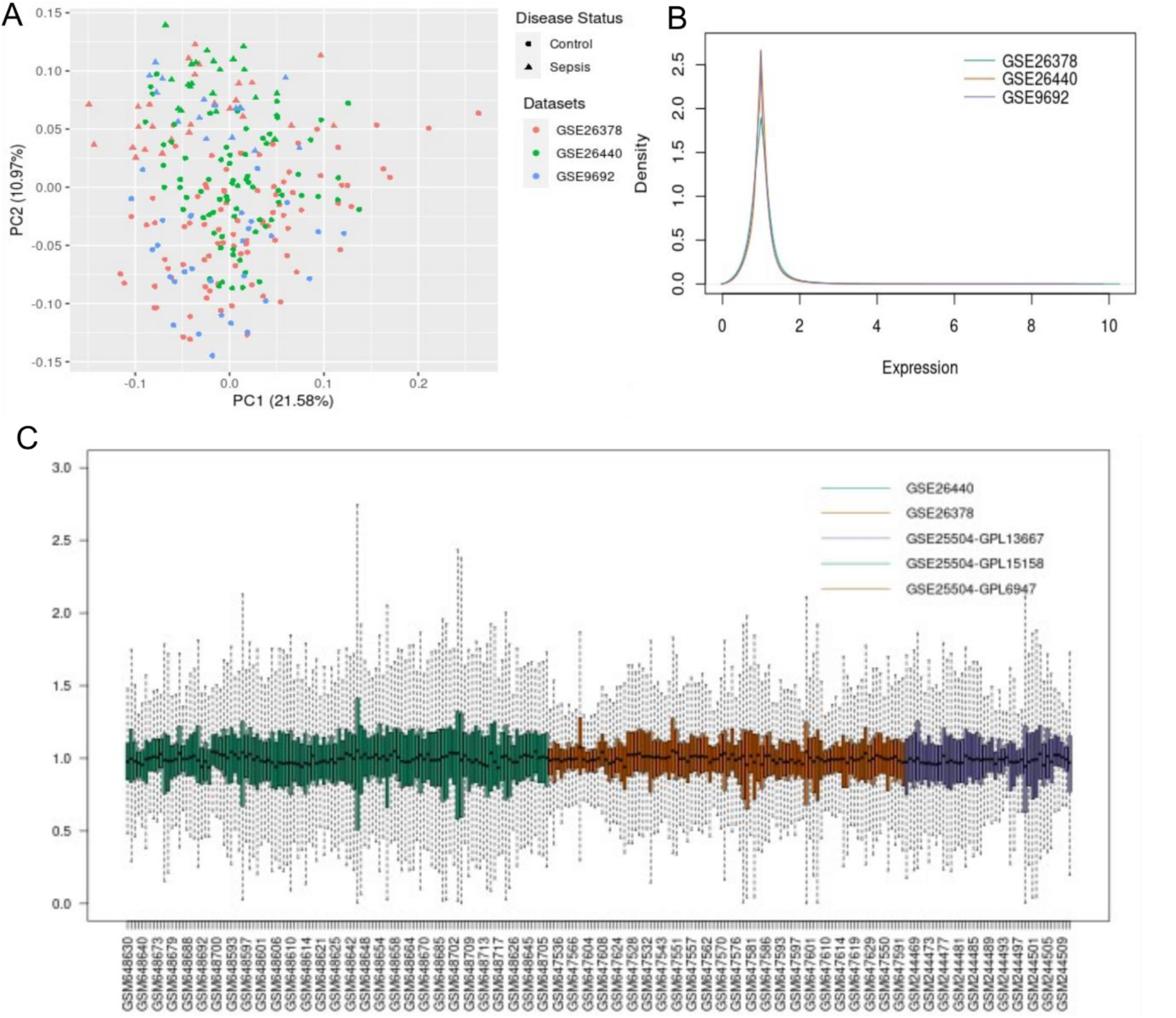
Batch effect in pediatric datasets (GSE26378, GSE26440, GSE9692) (Validation set-3). (A) PCA plot showing no batch effect. (B) Density plot is in consistent with PCA plot. (C) Sample level gene expression box plots show no batch effect in three datasets.

**Supplementary figure 12:**
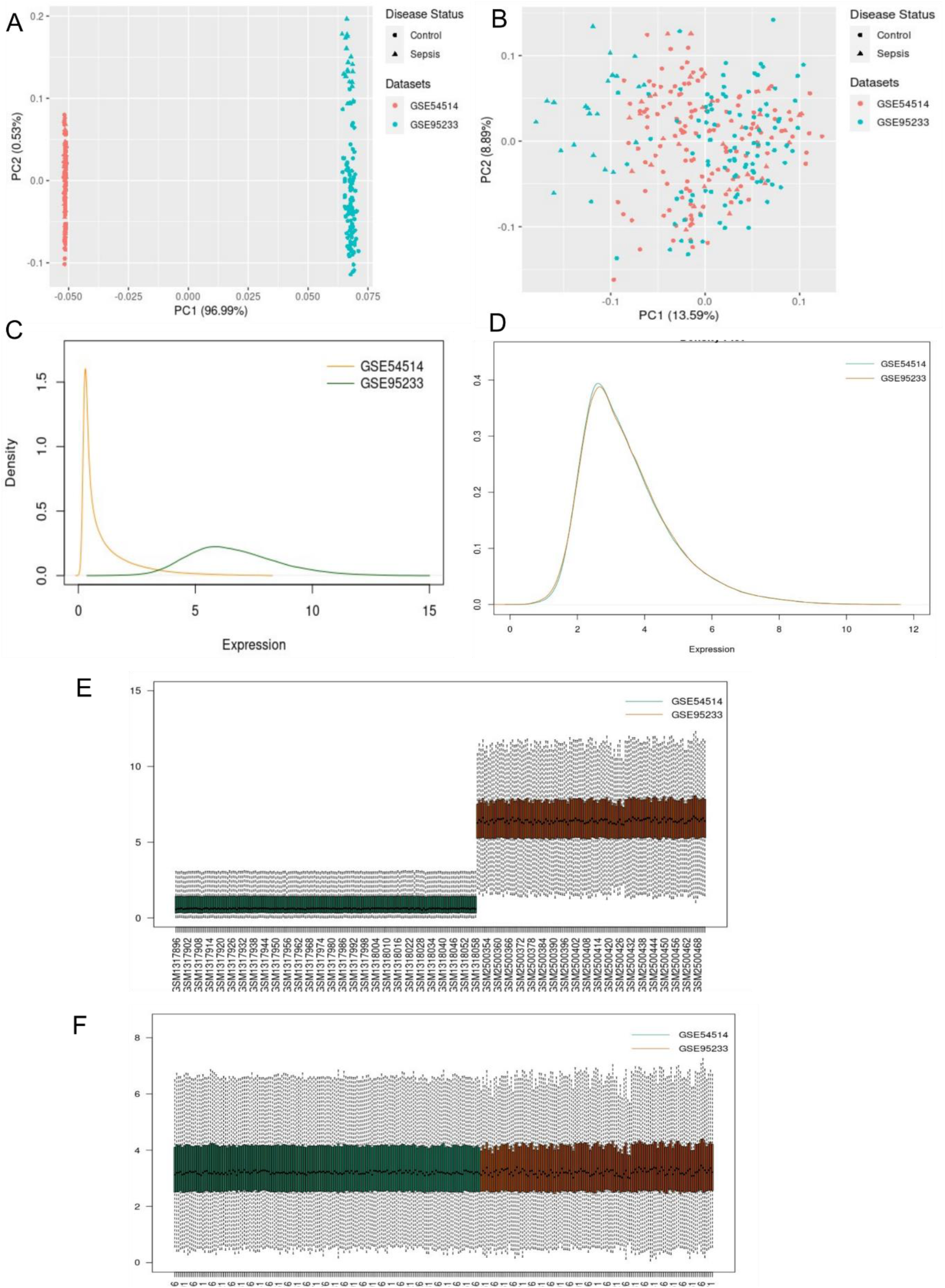
Batch effect in adult datasets (GSE54514, GSE95233) (Validation set-4). (A) PCA plot before batch effect correction. (B) PCA plot after batch effect correction. (C) Density plot before batch effect correction. (D) Density plot after batch effect correction. (E) Expression distribution plot of samples for the two datasets before batch effect correction. (F) Expression distribution plot of samples for the two

**Supplementary figure 13:**
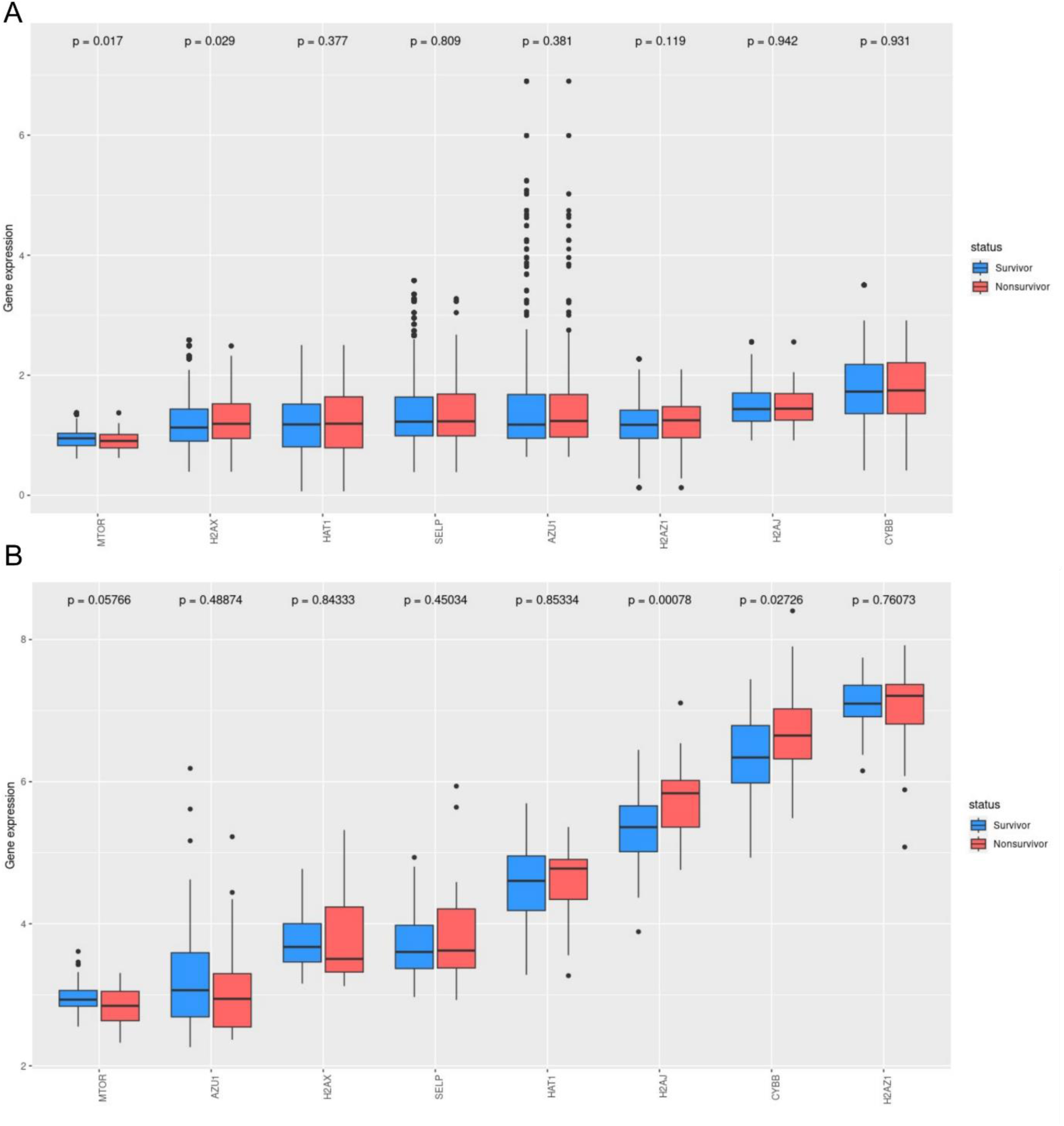
key-NET gene expression in other age groups. (A) Pediatric sepsis samples (Validation set-3). (B) Adult sepsis samples (Validation set-4)

**Supplementary figure 14:**
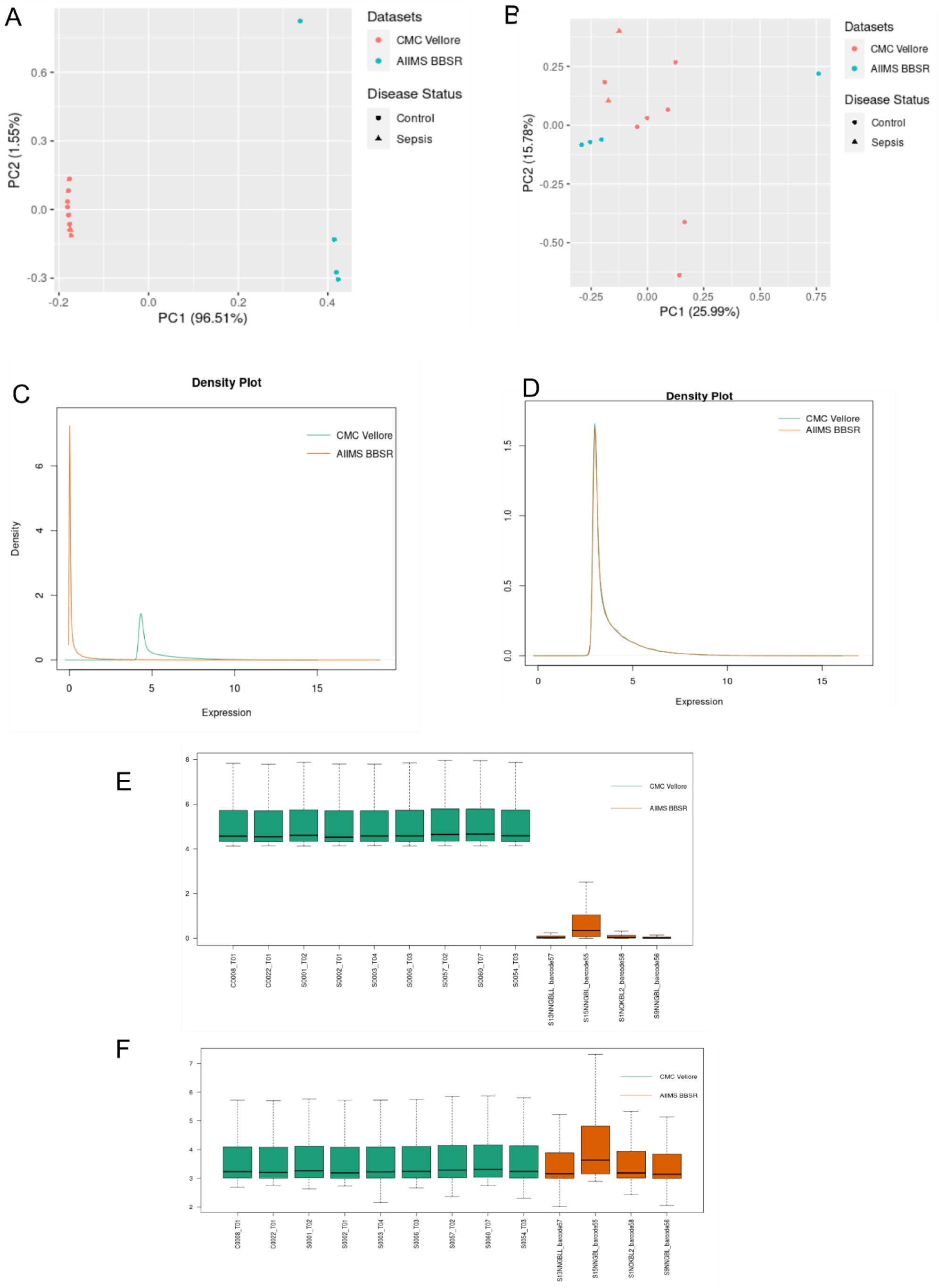
Batch effect in neonatal clinical samples (CMC Vellore, AIIMS BBSR) (Validation set-5). (A) PCA plot before batch effect correction. (B) PCA plot after batch effect correction. (C) Density plot before batch effect correction. (D) Density plot

**Supplementary figure 15:**
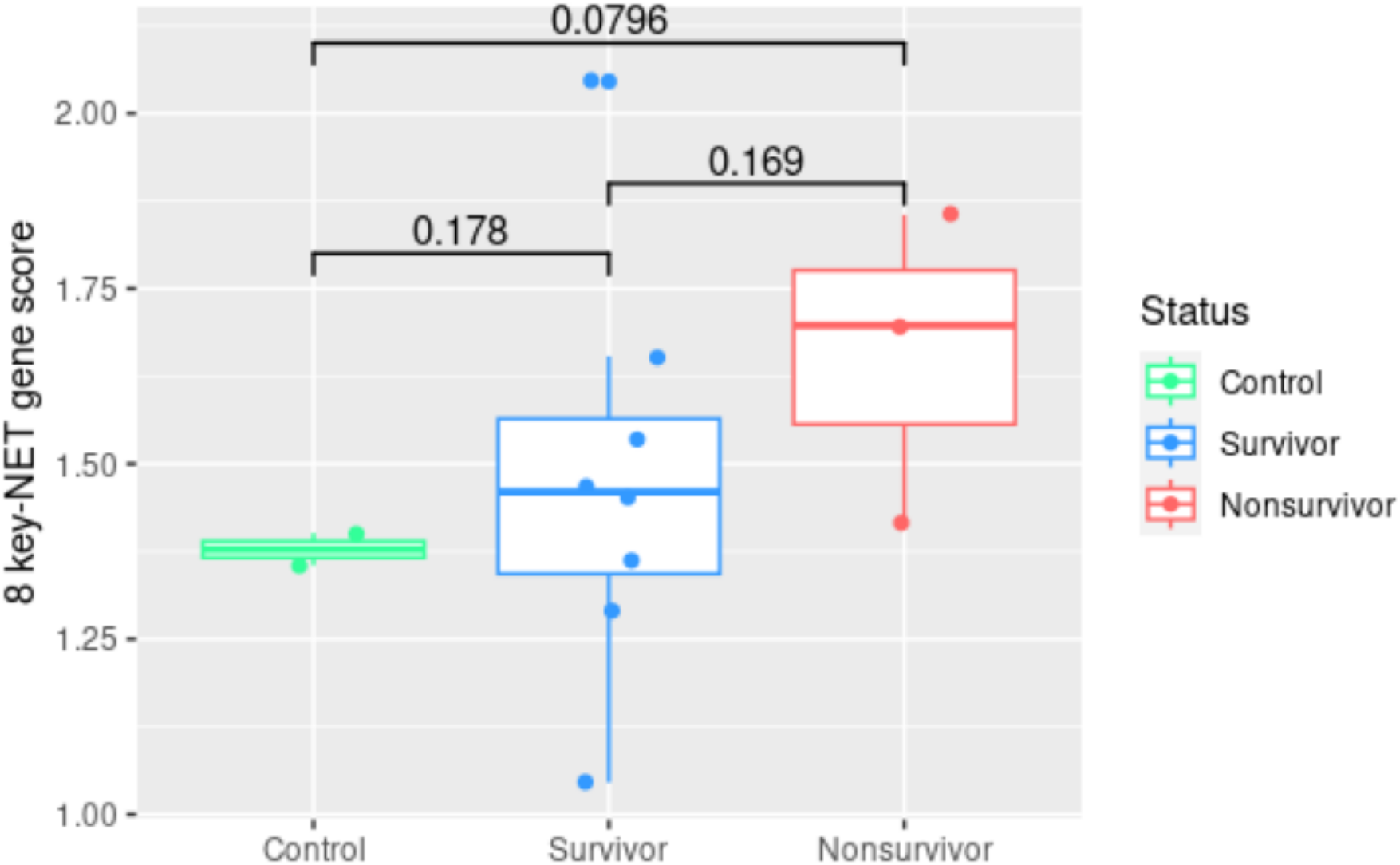
NET score model. There is an increasing trend of NET score in clinical samples of neonatal sepsis compared to control.

**Supplementary Table 1.**
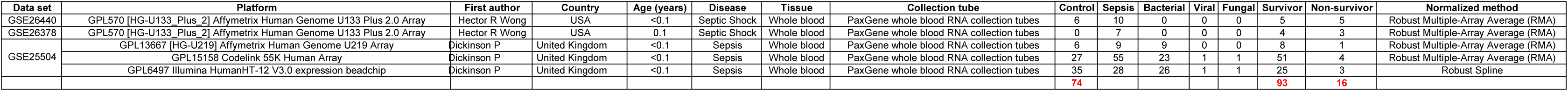
Discovery (60%) and Validation set-1 (40%)

**Supplementary Table 2.**

|  | Data set | Platform | First author | Country | Age (years) | Disease | Tissue | Collection tube | Control | Time point | Survivor | Non-survivor | Normalized method |
| --- | --- | --- | --- | --- | --- | --- | --- | --- | --- | --- | --- | --- | --- |
| Validation set-2 | GSE8121 | [HG-U133_Plus_2] Affymetrix Human Genome U133 Plus 2.0 Array | Hector R Wong | USA | < 10 years | Septic shock | Whole blood | PaxGene whole blood RNA collection tubes | 15 | day1 - day3 | 0 | 0 | Robust Multiple-Array Average (RMA) |
| Validation set-3 | GSE26440 | GPL570 [HG-U133_Plus_2] Affymetrix Human Genome U133 Plus 2.0 Array | Hector R Wong | USA | 0.2-10.9 | Septic Shock | Whole blood | PaxGene whole blood RNA collection tubes | 26 | 0 | 76 | 12 | Robust Multiple-Array Average (RMA) |
|  | GSE26378 | GPL570 [HG-U133_Plus_2] Affymetrix Human Genome U133 Plus 2.0 Array | Hector R Wong | USA | 0.2-9.8 | Septic Shock | Whole blood | PaxGene whole blood RNA collection tubes | 21 | 0 | 66 | 9 | Robust Multiple-Array Average (RMA) |
|  | GSE9692 | GPL570 [HG-U133_Plus_2] Affymetrix Human Genome U133 Plus 2.0 Array | Hector R Wong | USA | < 10 years | Septic Shock | Whole blood | PaxGene whole blood RNA collection tubes | 0 | 0 | 39 | 6 | Robust Multiple-Array Average (RMA) |
| Validation set-4 | GSE54514 | GPL6947[Illumina HumanHT-12 V3.0 expression beadchip] | Parnell GP | Australia | 18-86 | Sepsis | Whole blood | PaxGene whole blood RNA collection tubes | 18 | day1 | 26 | 9 | Quantile-normalised |
|  | GSE95233 | GPL570 [HG-U133_Plus_2] Affymetrix Human Genome U133 Plus 2.0 Array | Tabone O | France | 53-76 | Septic Shock | Whole blood | PaxGene whole blood RNA collection tubes | 22 | day1 | 34 | 17 | Robust Multiple-Array Average (RMA) |

**Supplementary Table 3.**
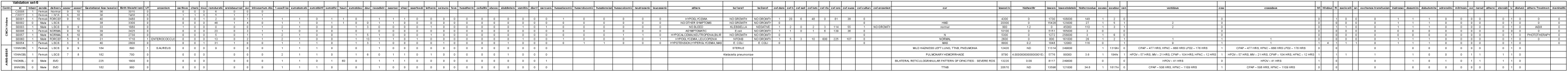
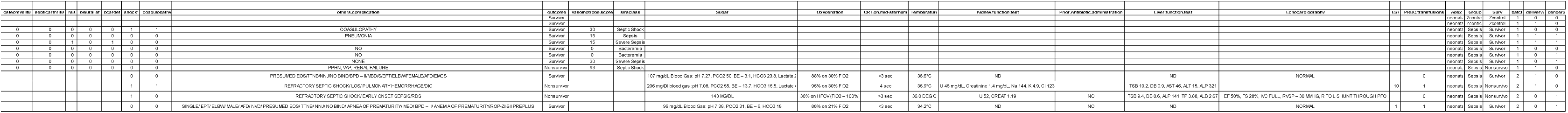

**Supplementary Table 4.**
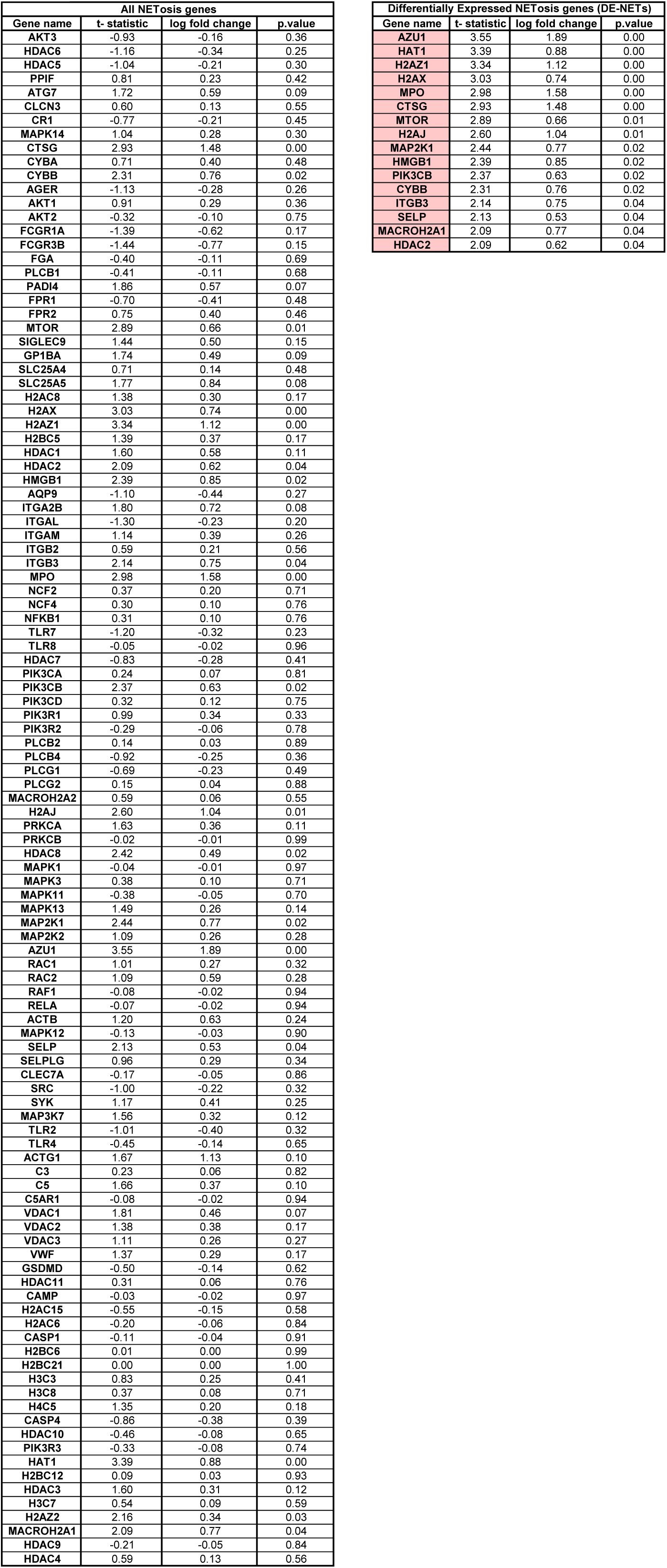

**Supplementary Table 5.**

| LASSO Coefficients |  |
| --- | --- |
| (Intercept) | -1.3055 |
| CTSG | 0.0000 |
| HDAC2 | 0.0000 |
| HMGB1 | 0.0000 |
| ITGB3 | 0.0000 |
| MPO | 0.0000 |
| PIK3CB | 0.0000 |
| MAP2K1 | 0.0000 |
| MACROH2A1 | 0.0000 |
| H2AJ | 0.0050 |
| H2AZ1 | 0.0182 |
| CYBB | 0.0204 |
| MTOR | 0.0249 |
| AZU1 | 0.0291 |
| H2AX | 0.0375 |
| SELP | 0.0569 |
| HAT1 | 0.0735 |

**Supplementary Table 6.**
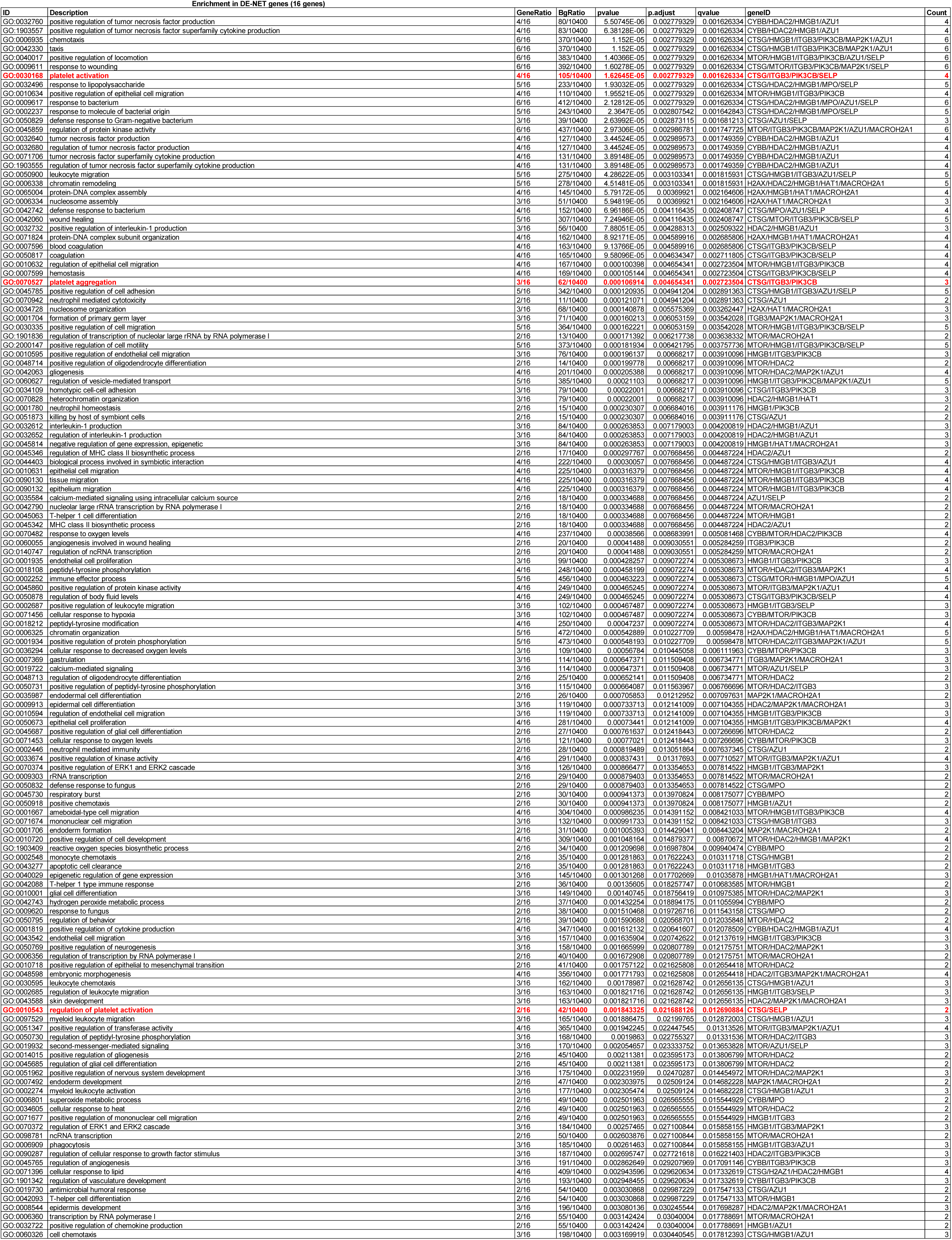
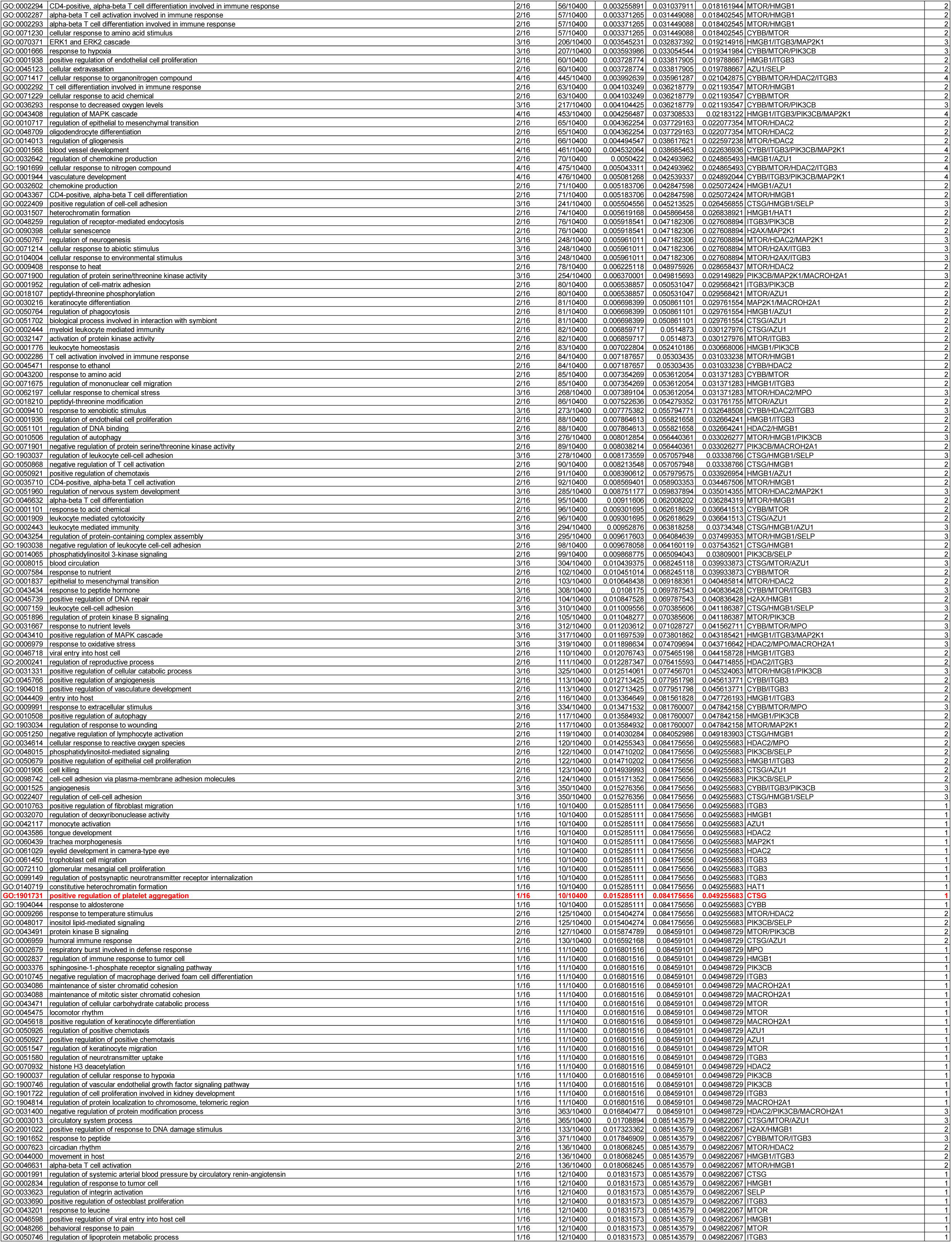
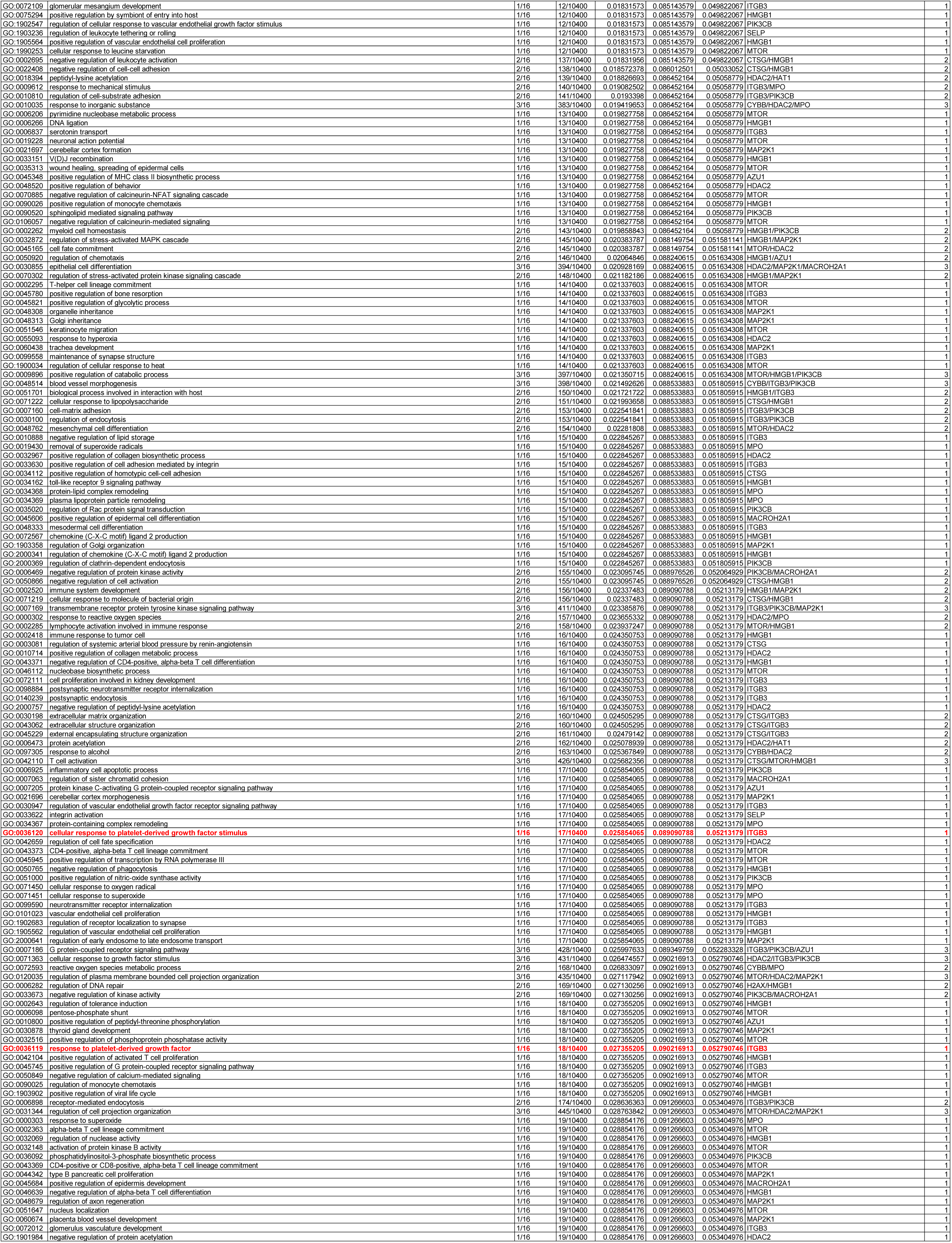
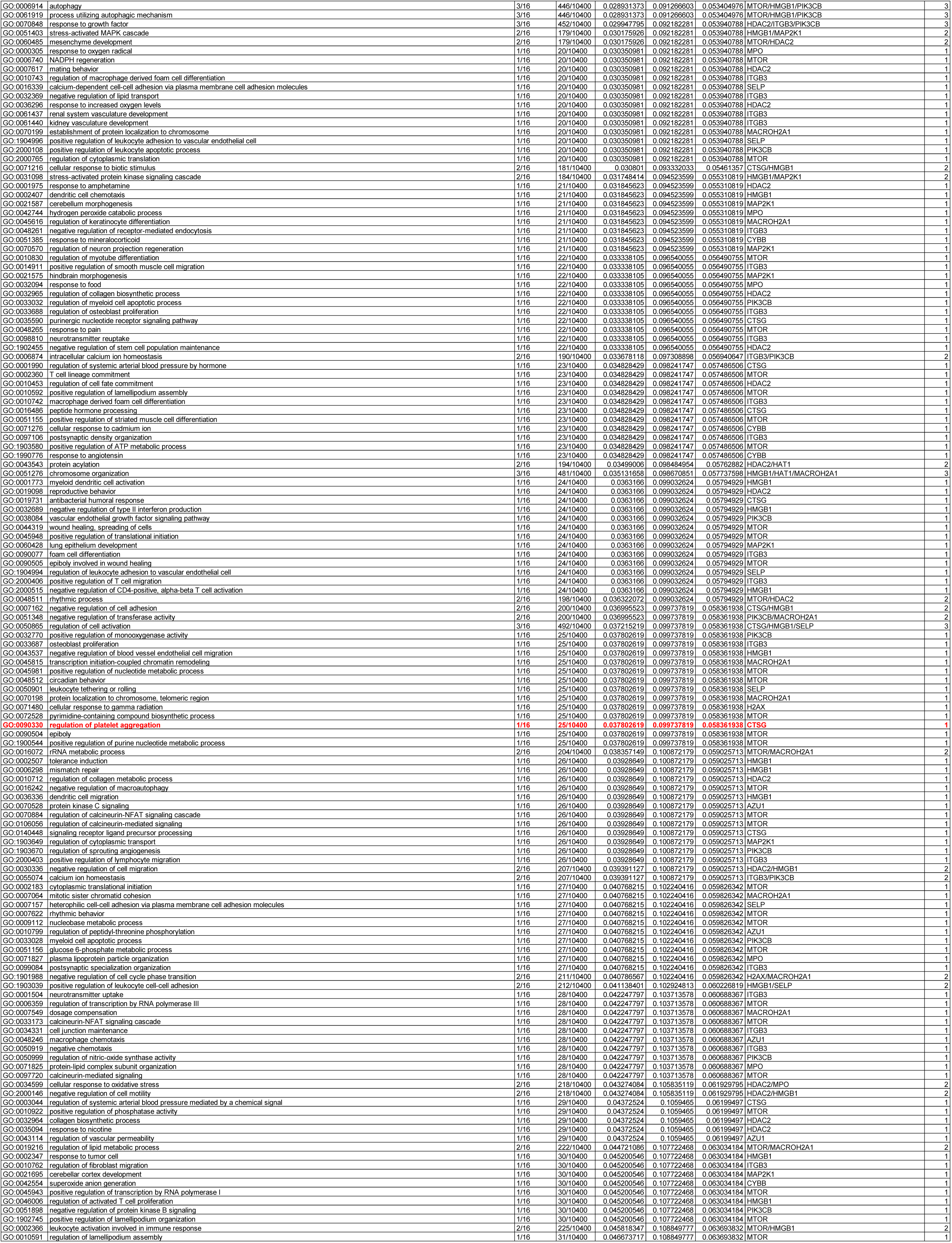
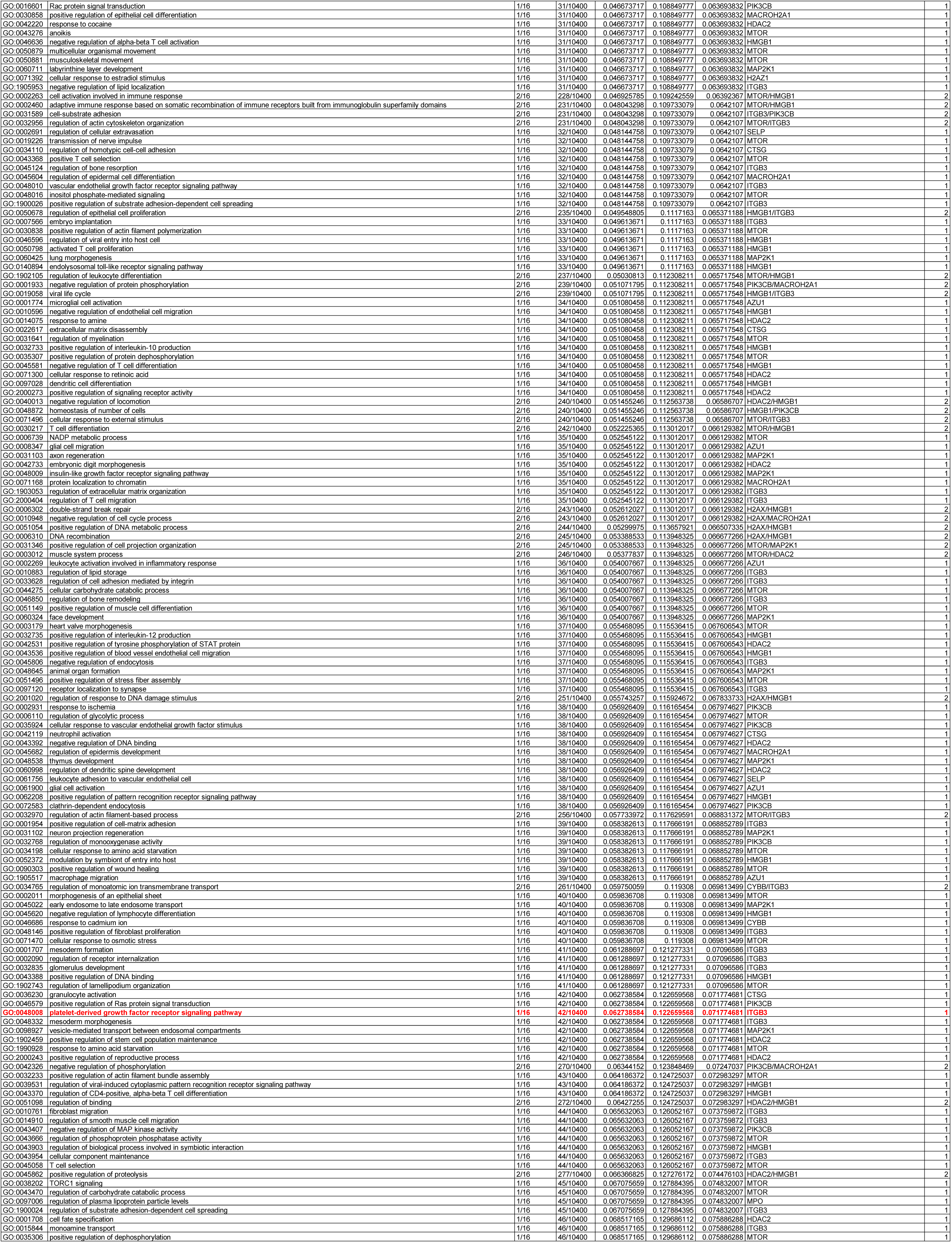
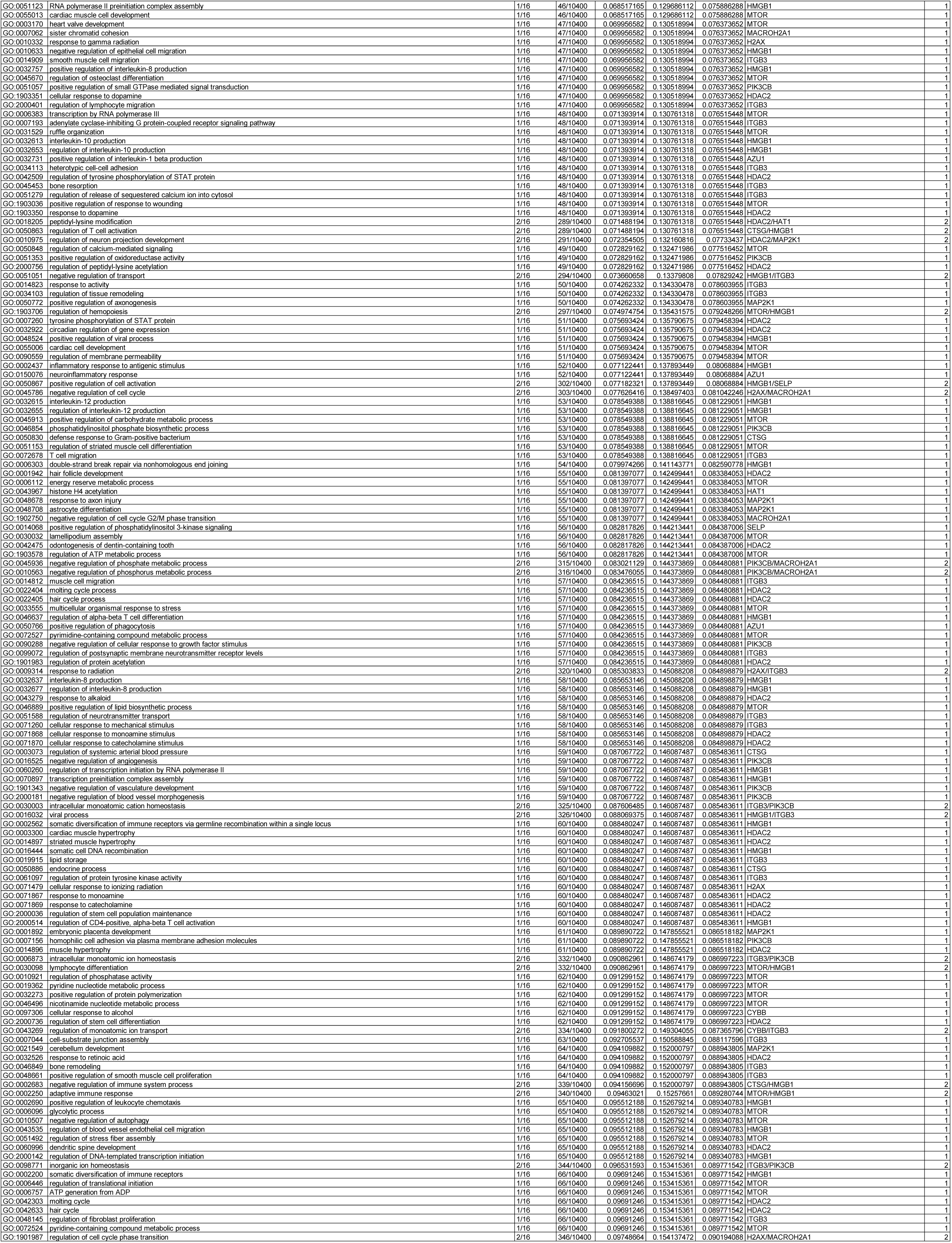
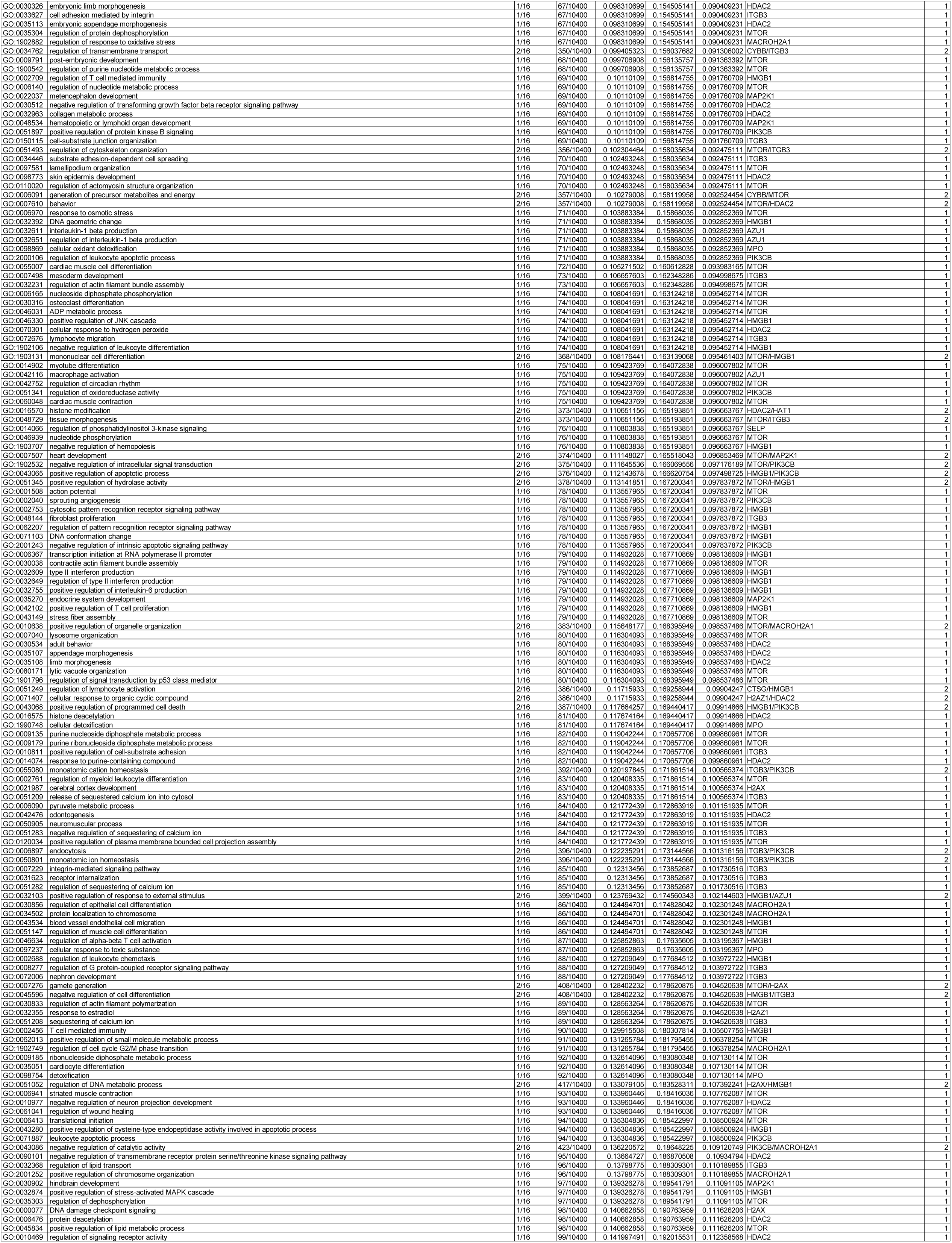
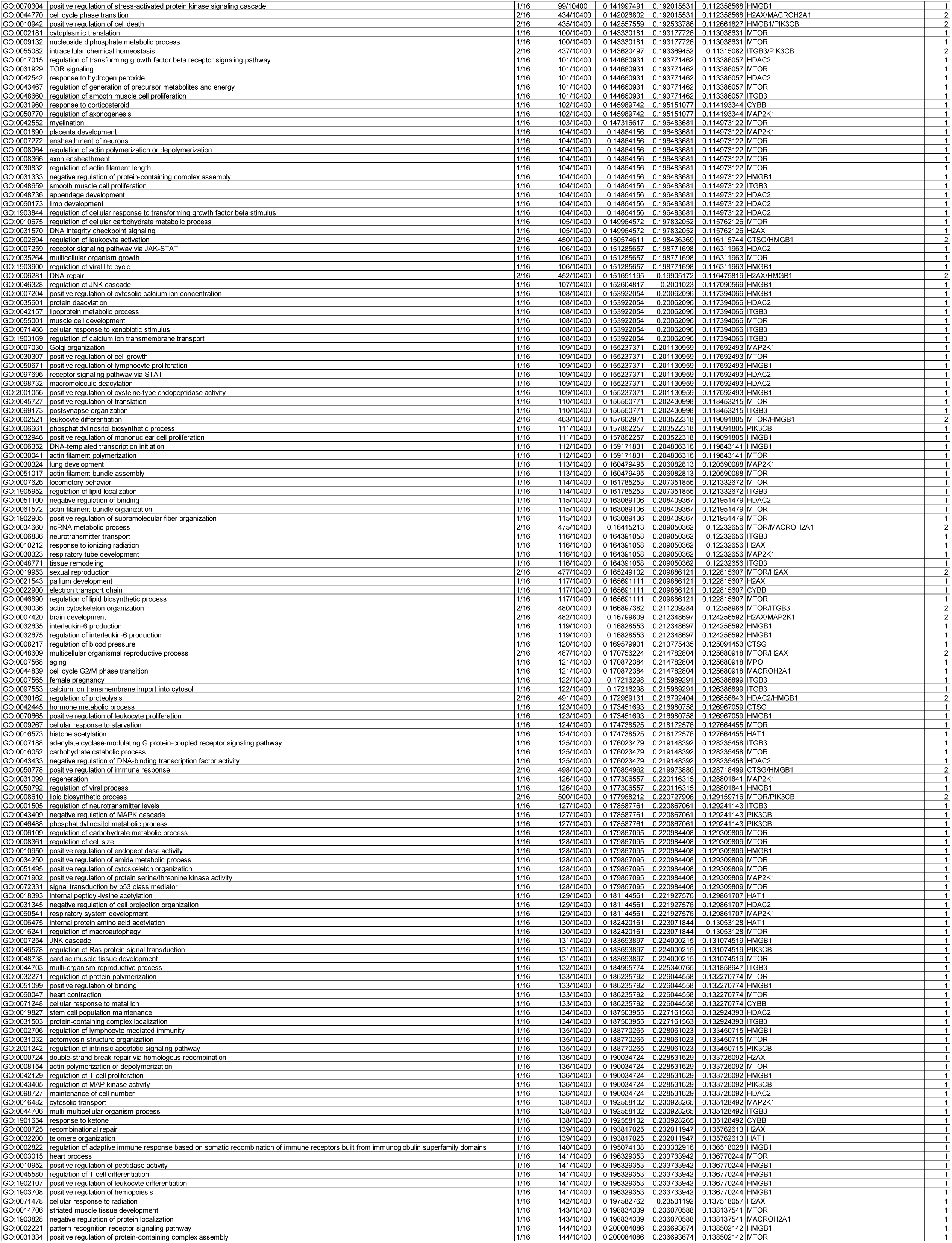
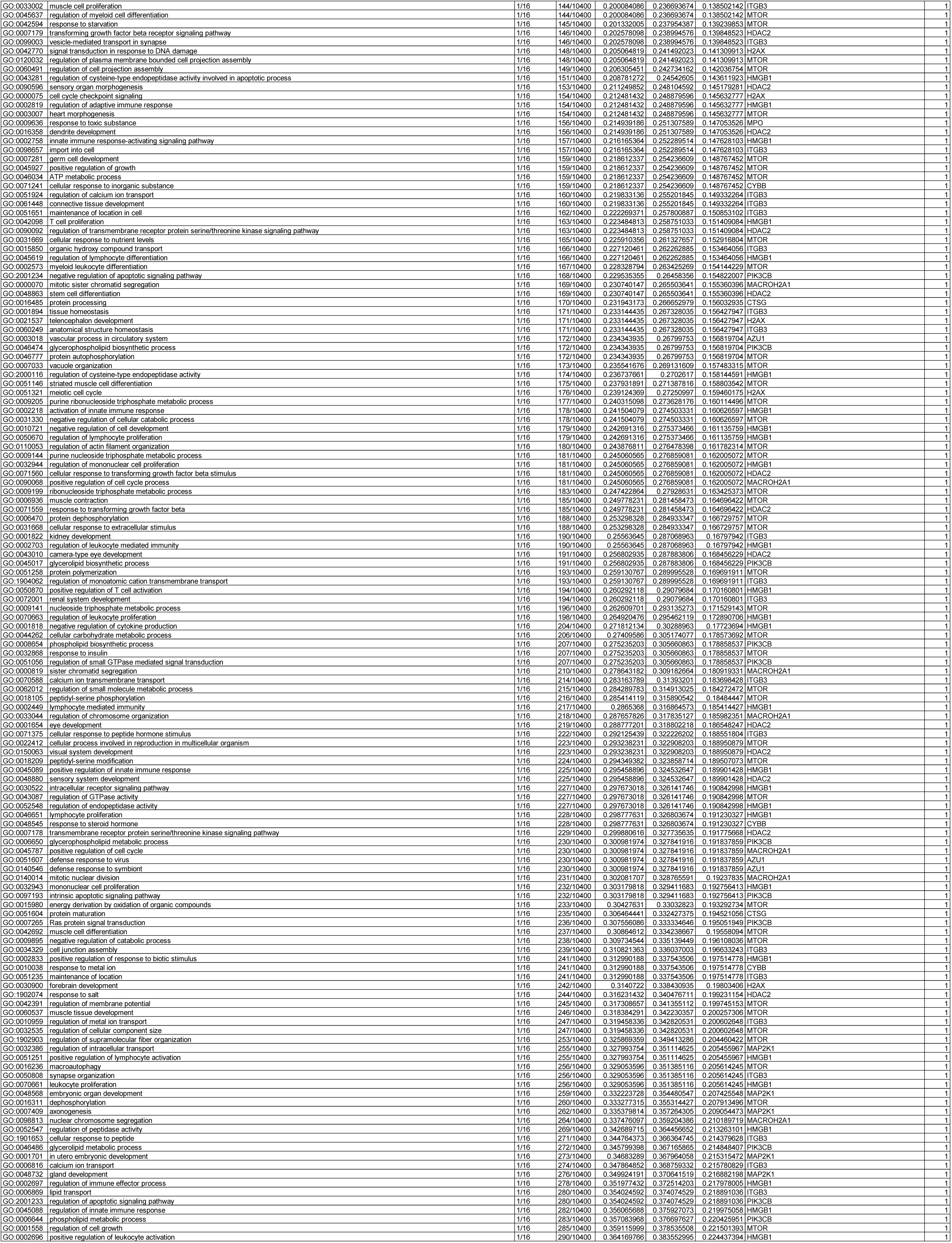
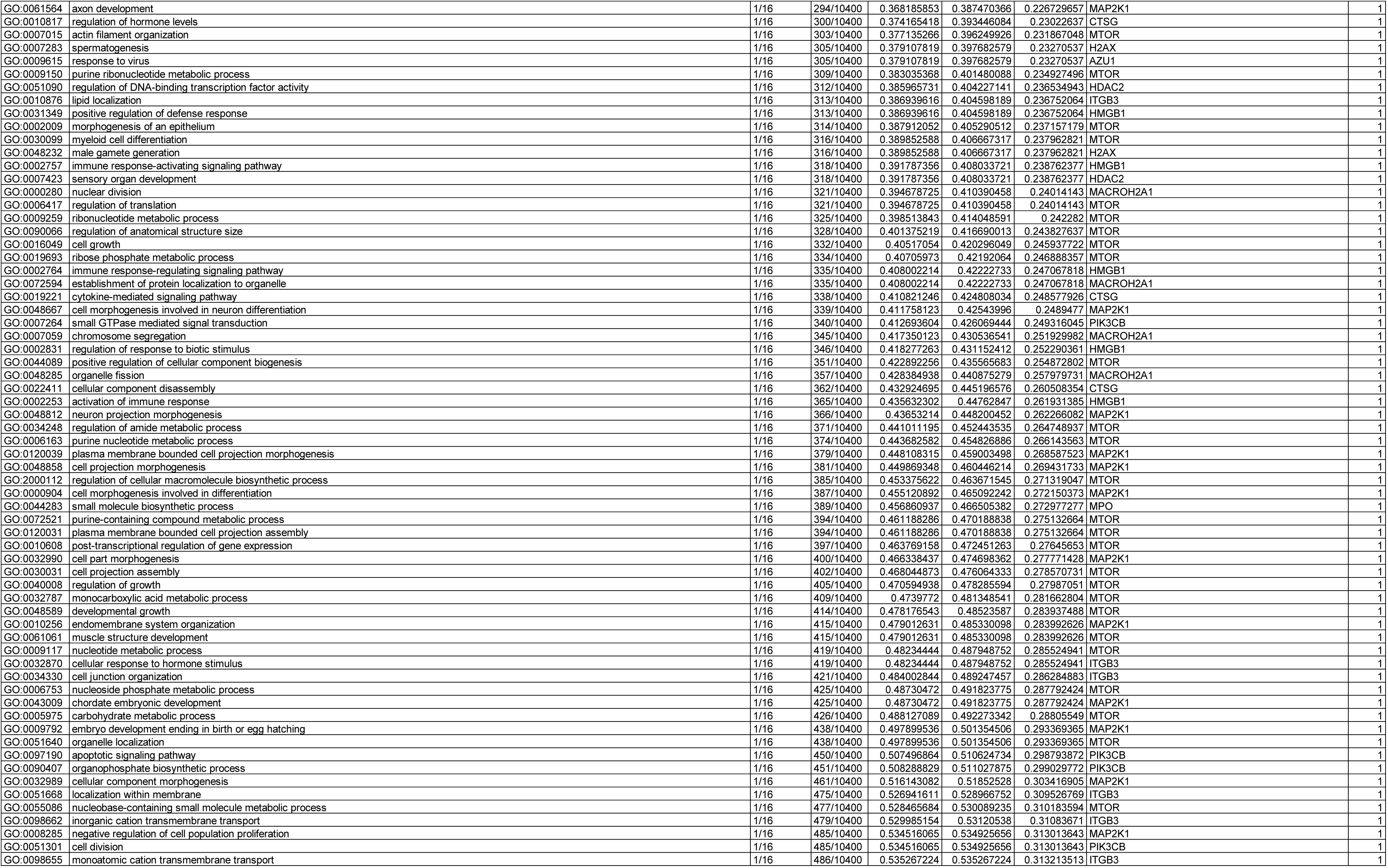

**Supplementary Table 7.**
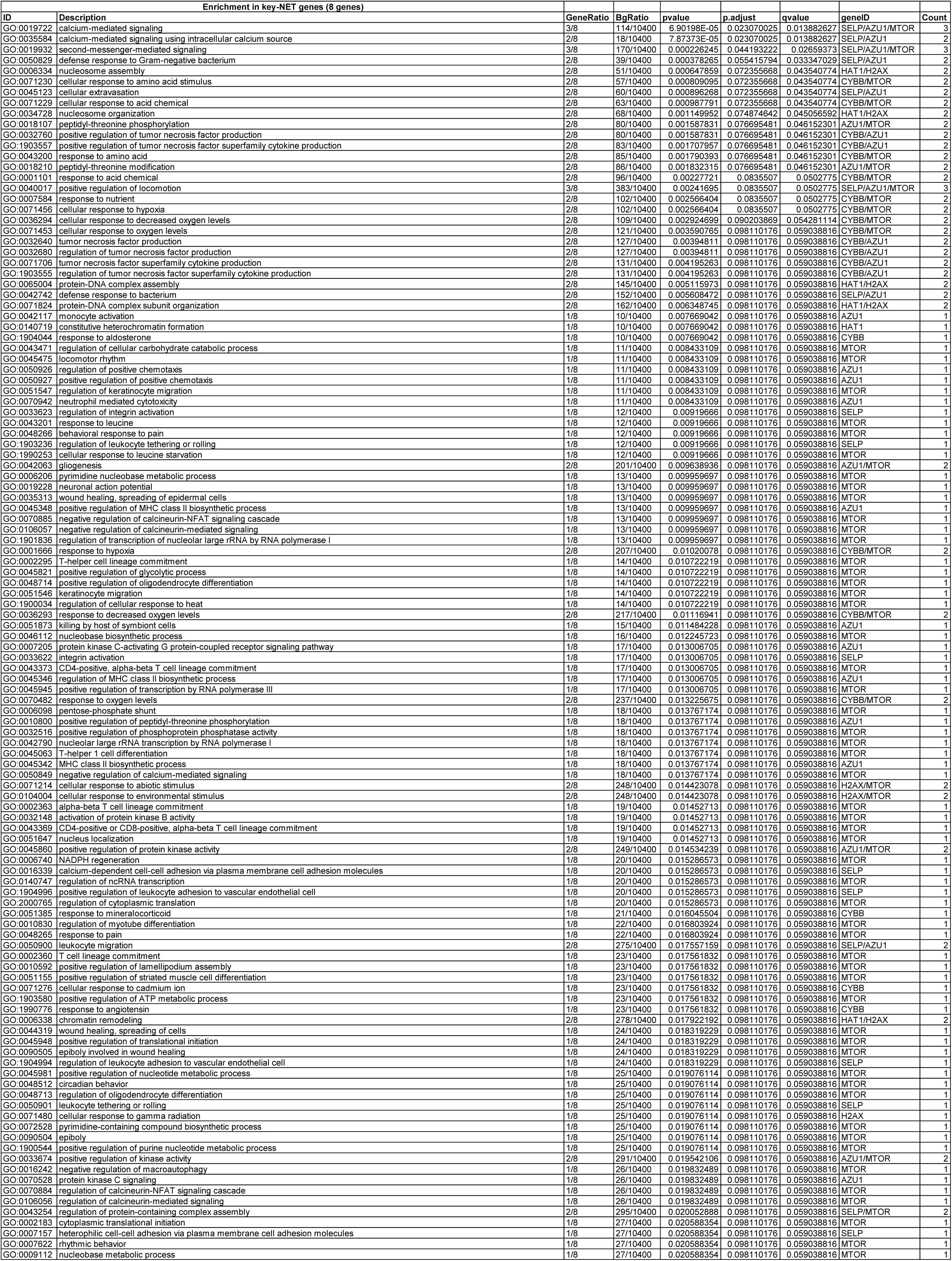
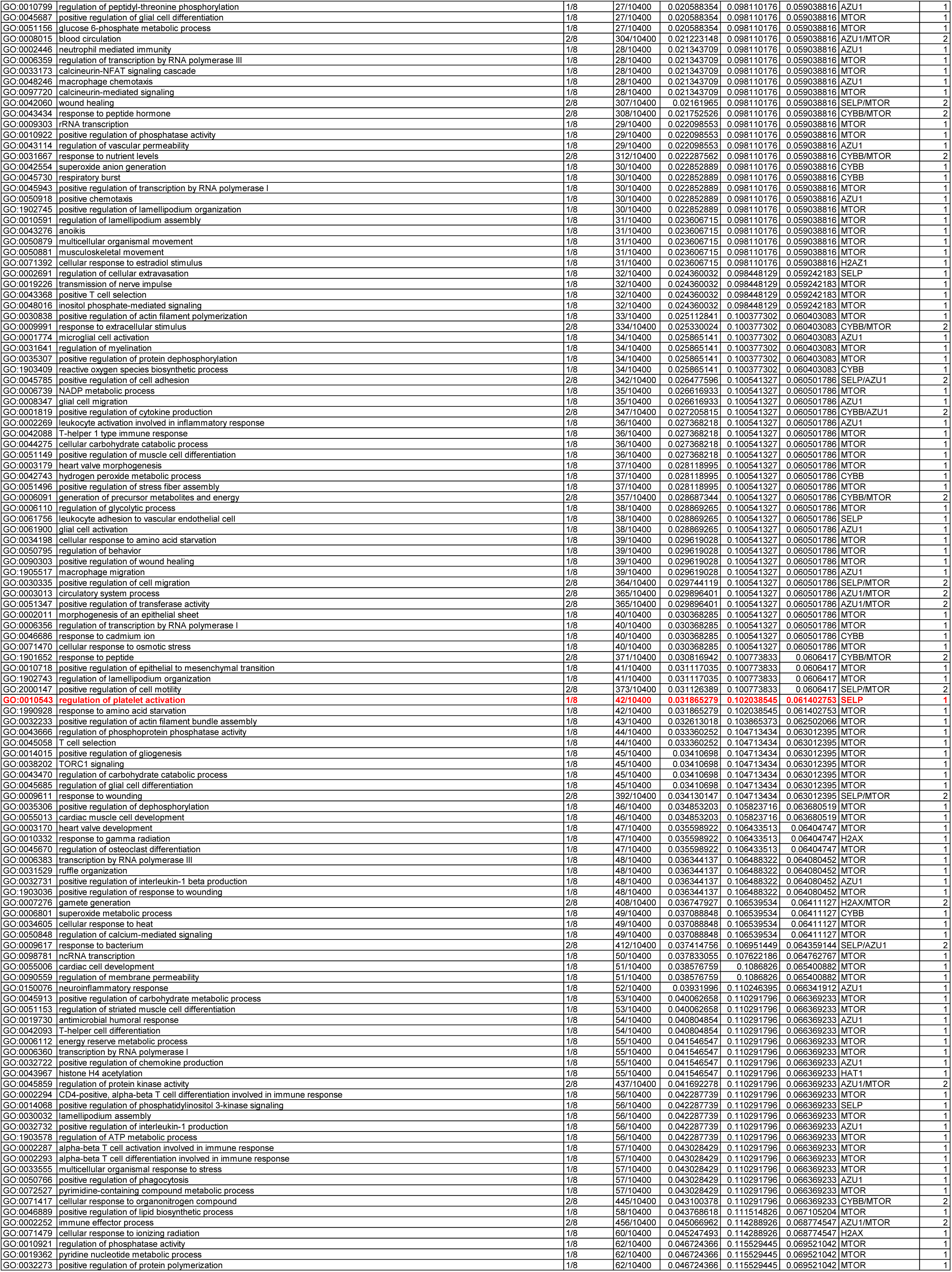
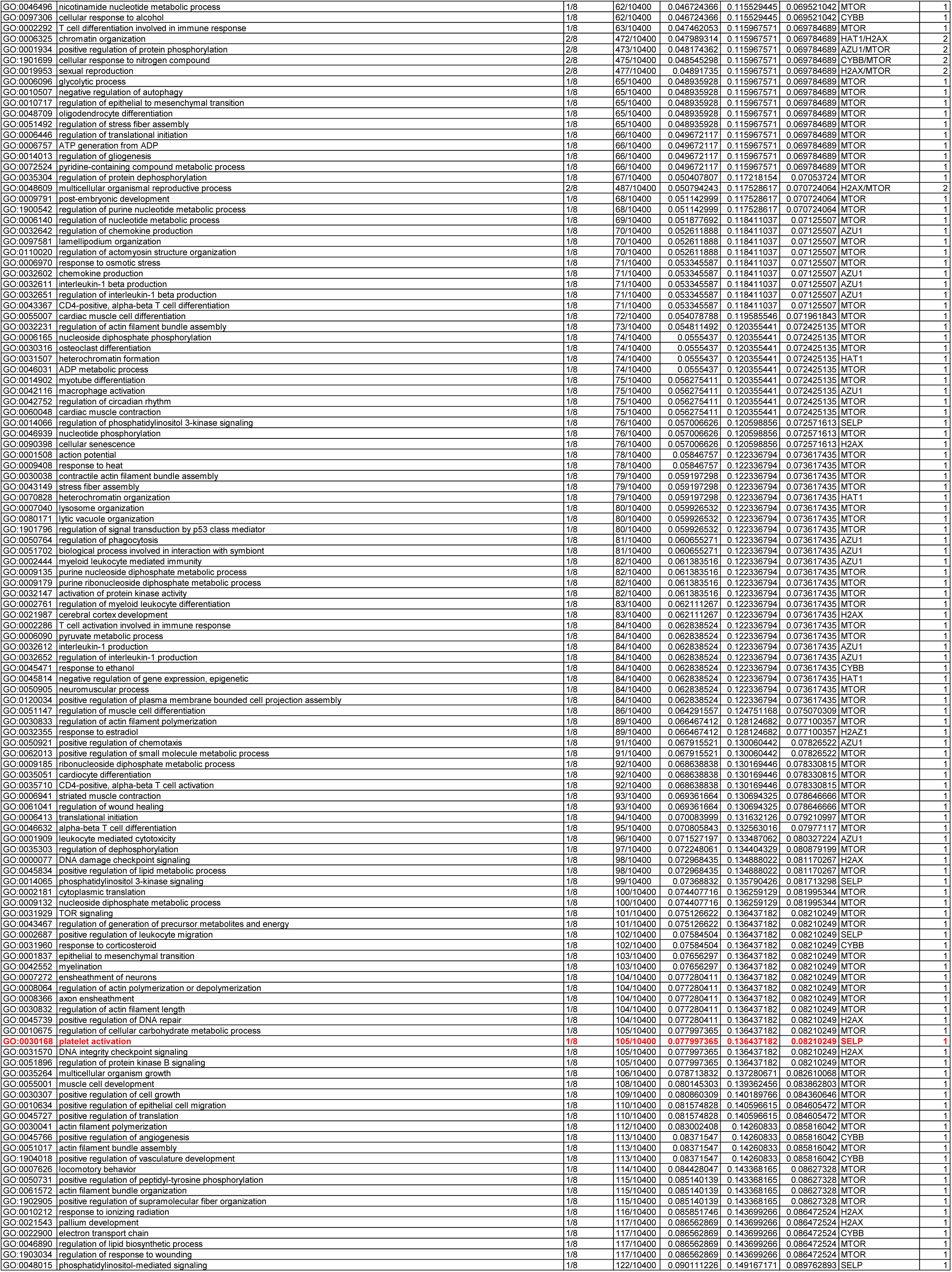
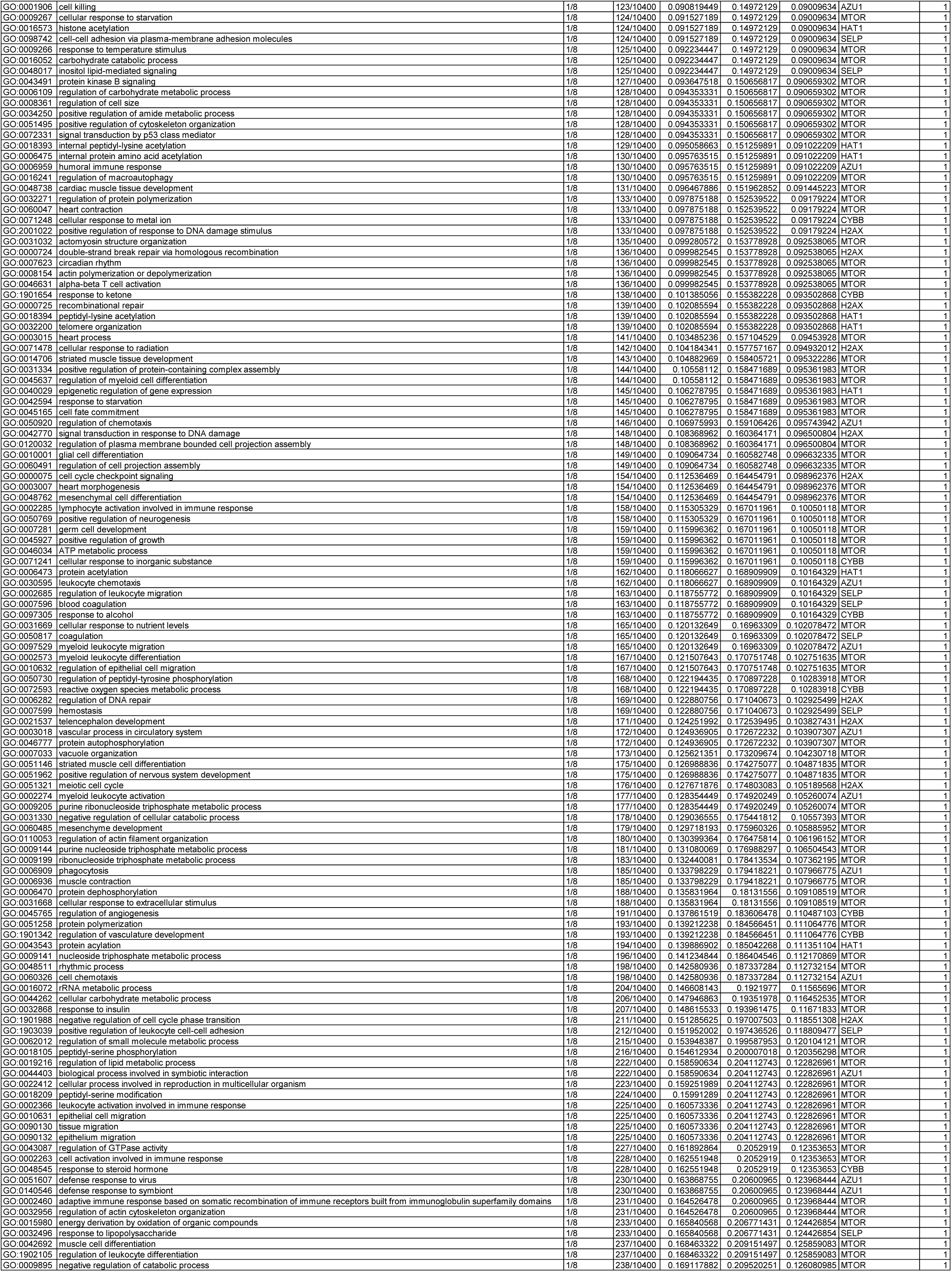
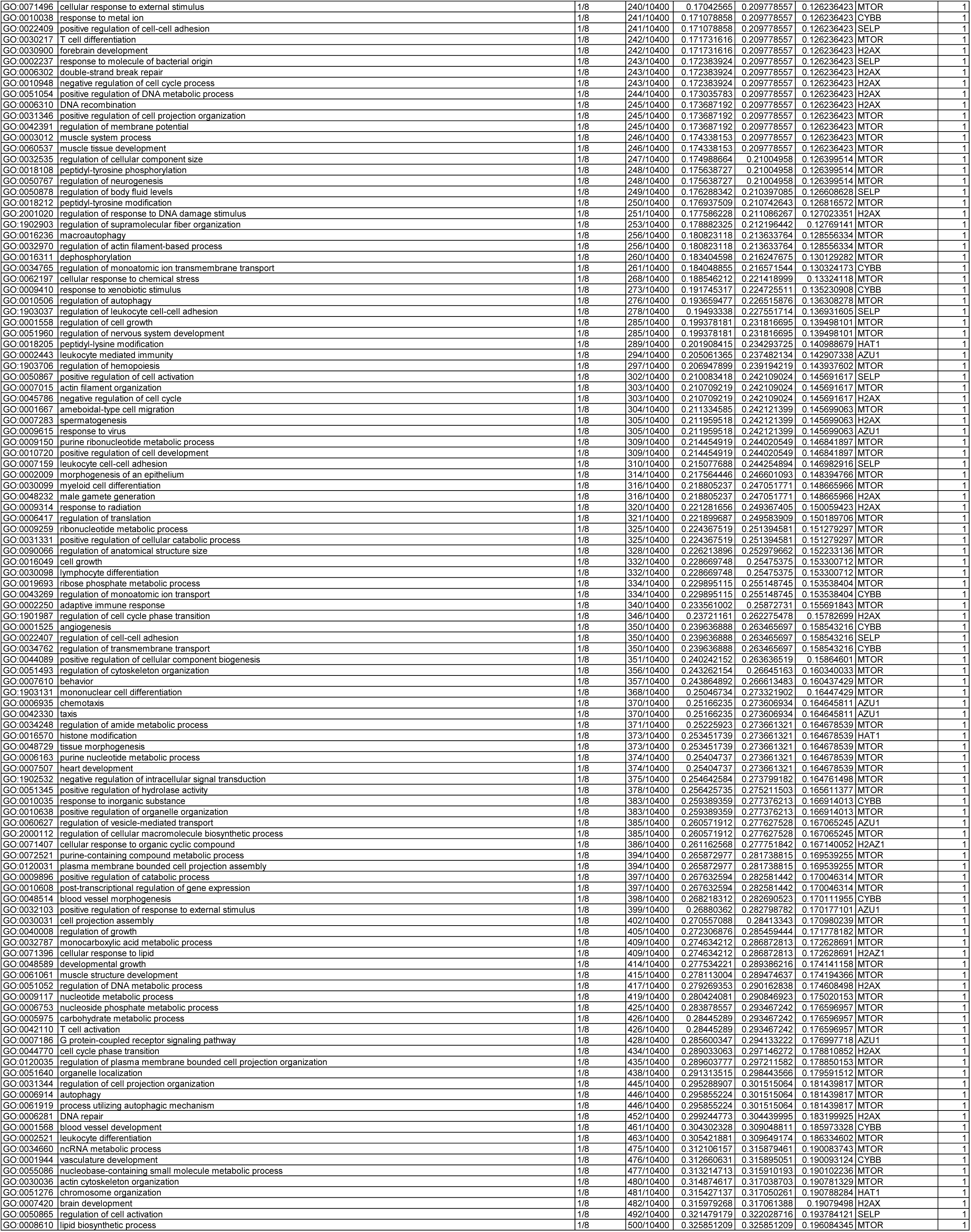

**Supplementary Table 8.**
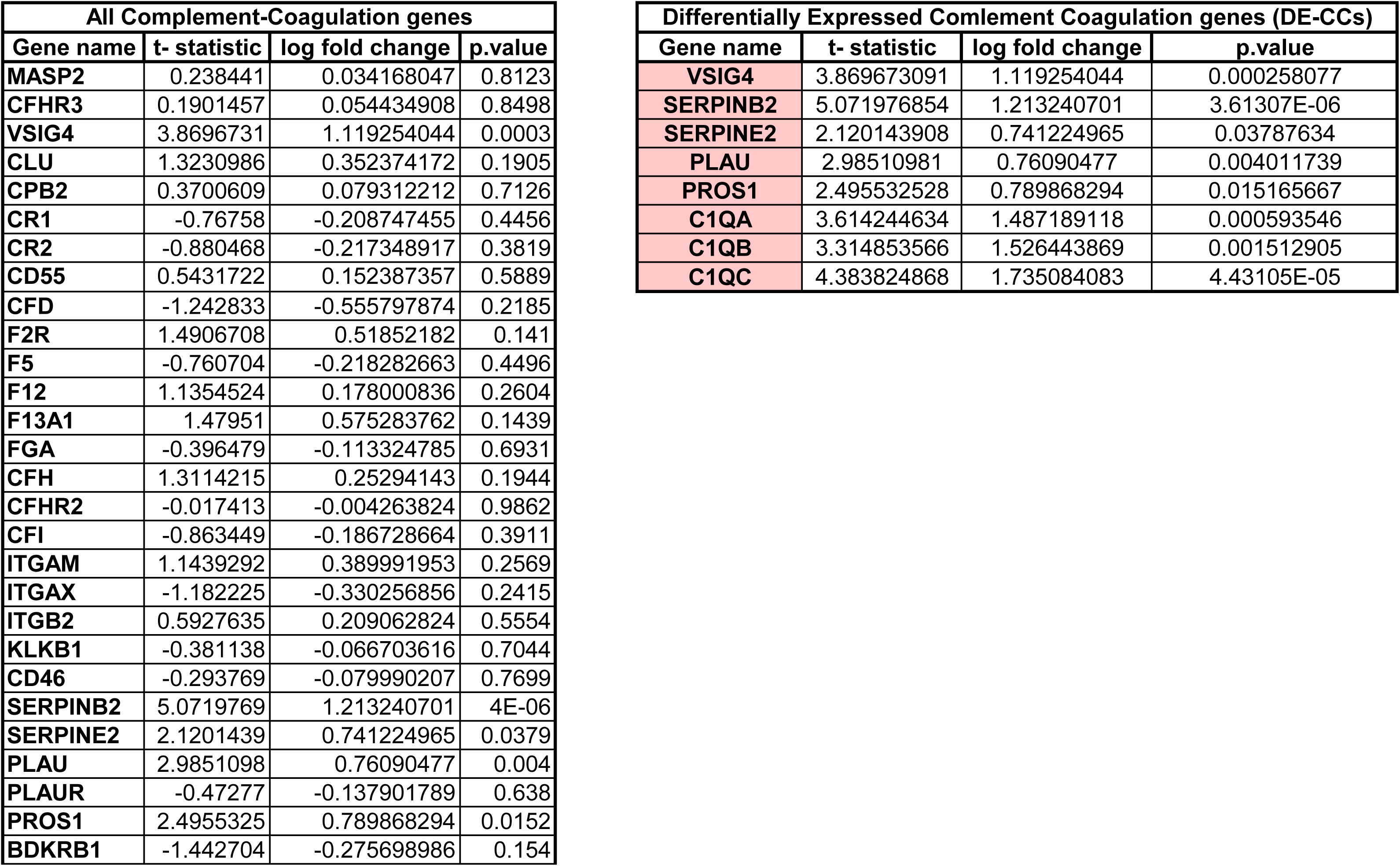
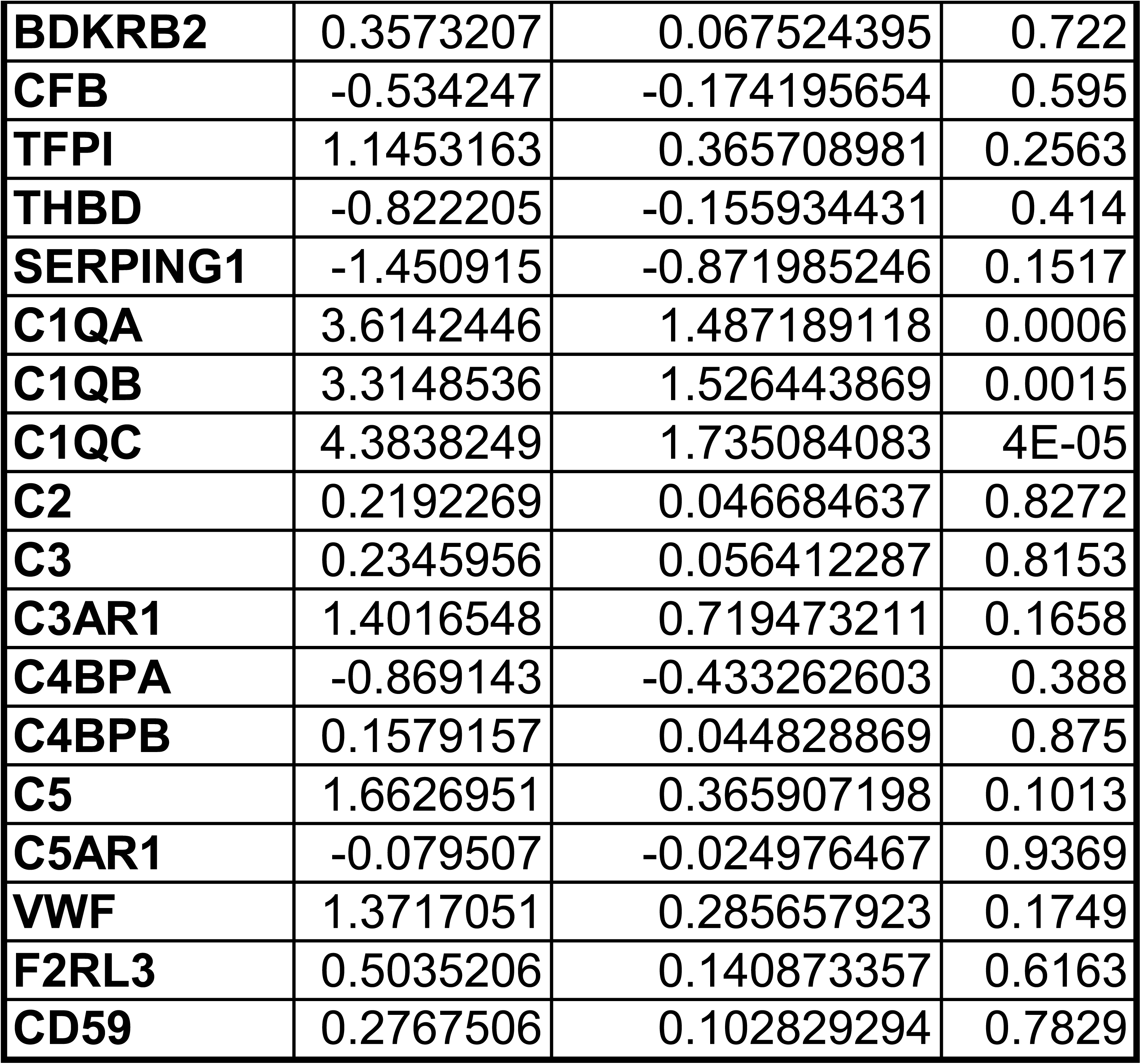

**Supplementary Table 9.**
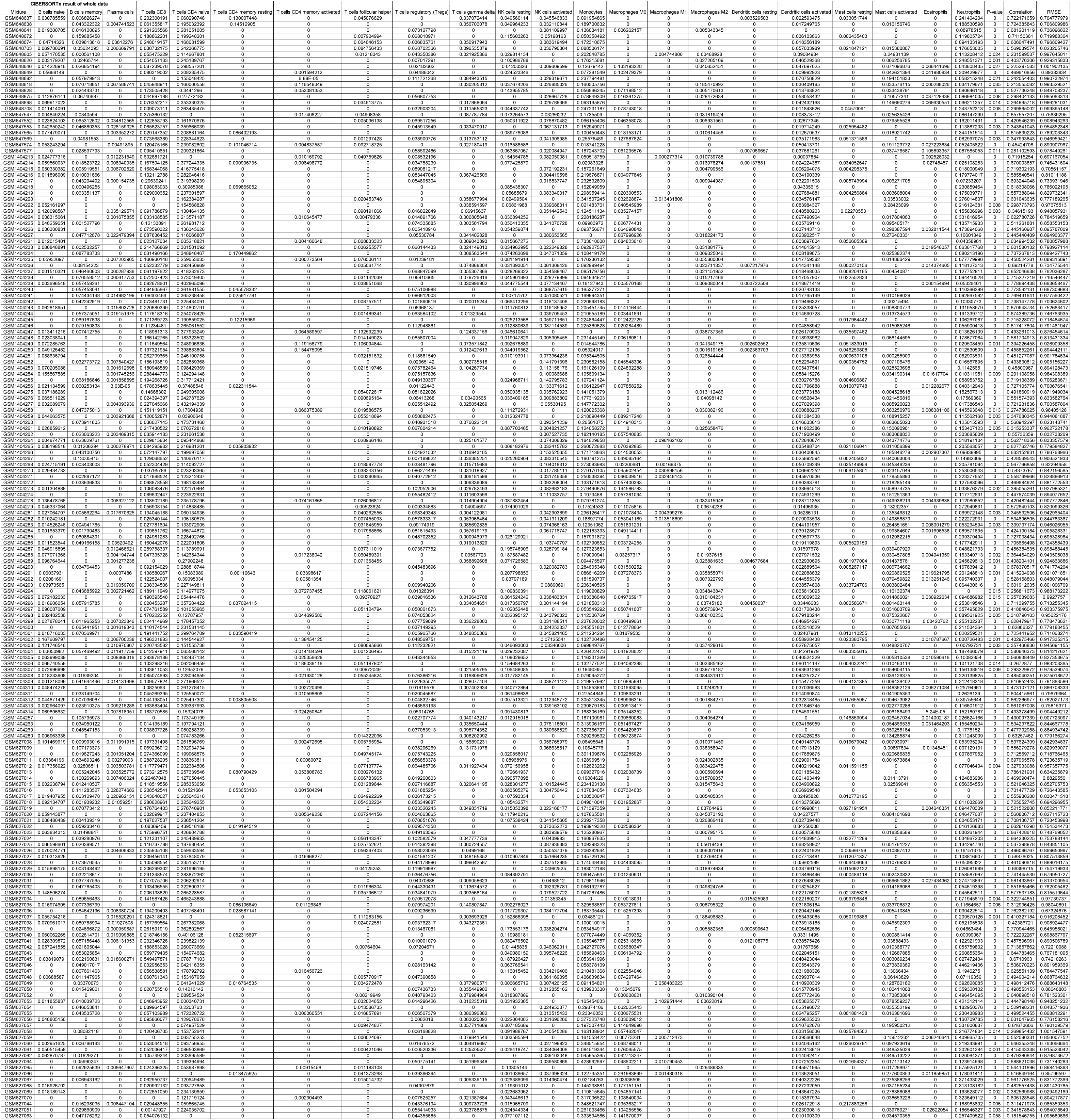

**Supplementary Table 10.**
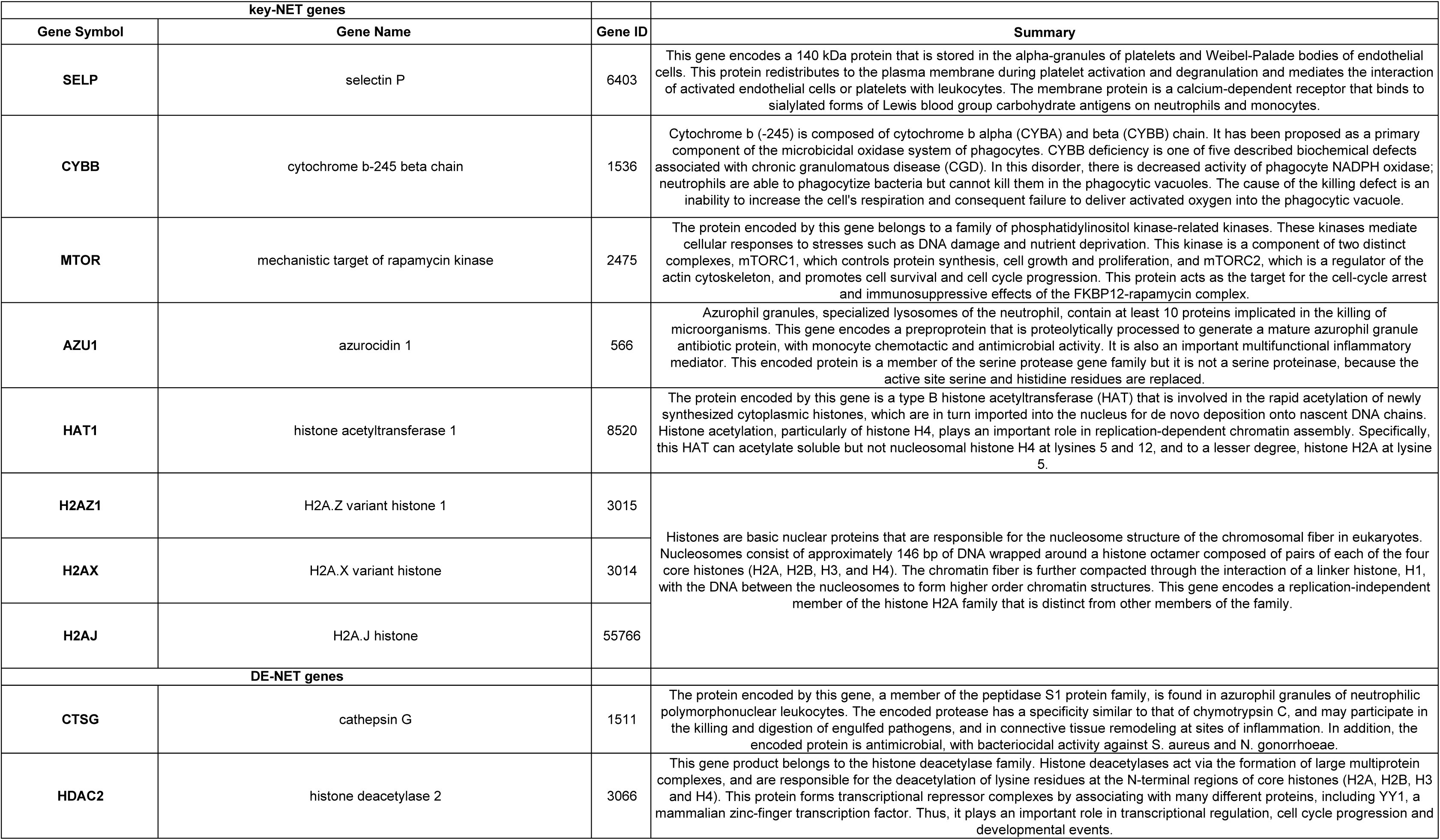
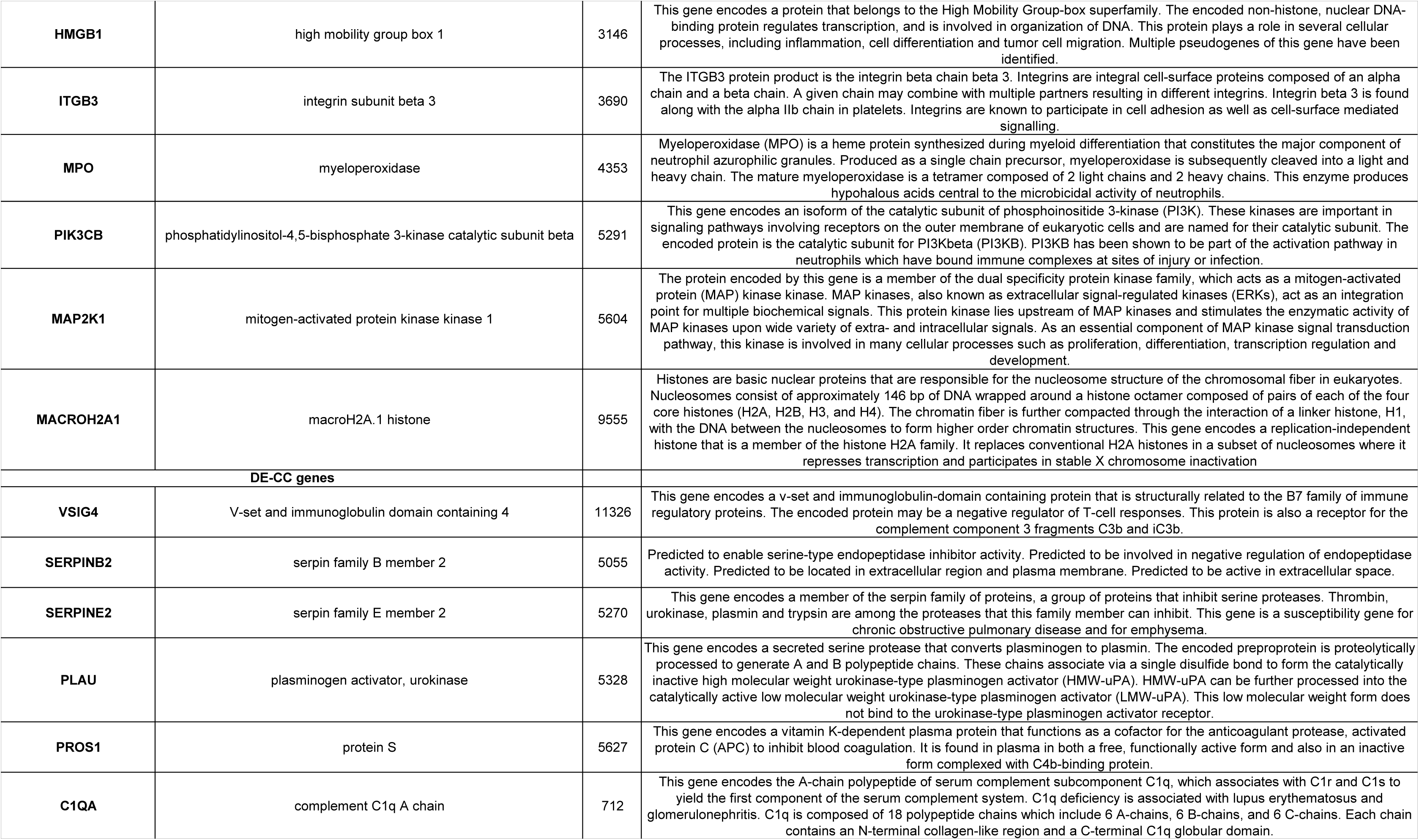
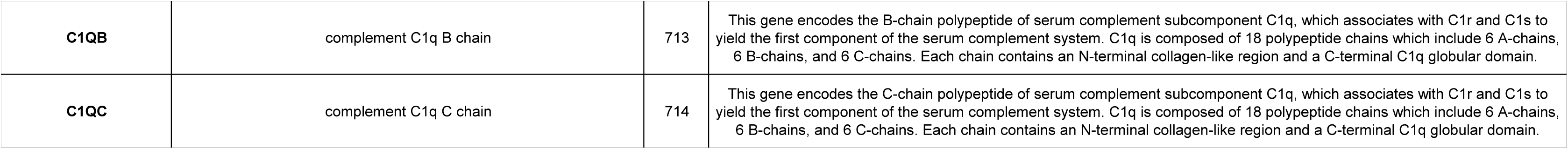

## Reference

1. Singer M, Deutschman CS, Seymour CW, Shankar-Hari M, Annane D, Bauer M, et al. The Third International Consensus Definitions for Sepsis and Septic Shock (Sepsis-3). JAMA. 2016 Feb 23;315(8):801–10.

2. Shane AL, Sánchez PJ, Stoll BJ. Neonatal sepsis. The Lancet. 2017 Oct 14;390(10104):1770–80.

3. Zonneveld R, Martinelli R, Shapiro NI, Kuijpers TW, Plötz FB, Carman CV. Soluble adhesion molecules as markers for sepsis and the potential pathophysiological discrepancy in neonates, children and adults. Critical Care. 2014 Feb 18;18(1):204.

4. Zhang X, Zhivaki D, Lo-Man R. Unique aspects of the perinatal immune system. Nat Rev Immunol. 2017 Aug;17(8):495–507.

5. Molloy EJ, Wynn JL, Bliss J, Koenig JM, Keij FM, McGovern M, et al. Neonatal sepsis: need for consensus definition, collaboration and core outcomes. Pediatr Res. 2020 Jul;88(1):2–4.

6. Fleischmann-Struzek C, Goldfarb DM, Schlattmann P, Schlapbach LJ, Reinhart K, Kissoon N. The global burden of paediatric and neonatal sepsis: a systematic review. The Lancet Respiratory Medicine. 2018 Mar 1;6(3):223–30.

7. Strunk T, Molloy EJ, Mishra A, Bhutta ZA. Neonatal bacterial sepsis. The Lancet. 2024 Jul 20;404(10449):277–93.

8. Schlapbach LJ, Watson RS, Sorce LR, Argent AC, Menon K, Hall MW, et al. International Consensus Criteria for Pediatric Sepsis and Septic Shock. JAMA. 2024 Feb 27;331(8):665–74.

9. Ziesmann MT, Marshall JC. Multiple Organ Dysfunction: The Defining Syndrome of Sepsis. Surgical Infections. 2018 Feb;19(2):184–90.

10. Khaertynov KS, Boichuk SV, Khaiboullina SF, Anokhin VA, Andreeva AA, Lombardi VC, et al. Comparative Assessment of Cytokine Pattern in Early and Late Onset of Neonatal Sepsis. J Immunol Res. 2017;2017:8601063.

11. Brinkmann V, Reichard U, Goosmann C, Fauler B, Uhlemann Y, Weiss DS, et al. Neutrophil Extracellular Traps Kill Bacteria. Science. 2004 Mar 5;303(5663):1532–5.

12. Delgado-Rizo V, Martínez-Guzmán MA, Iñiguez-Gutierrez L, García-Orozco A, Alvarado-Navarro A, Fafutis-Morris M. Neutrophil Extracellular Traps and Its Implications in Inflammation: An Overview. Front Immunol. 2017 Feb 6;8:81.

13. Kaplan MJ, Radic M. Neutrophil extracellular traps (NETs): Double-edged swords of innate immunity. J Immunol. 2012 Sep 15;189(6):2689–95.

14. de Bont CM, Boelens WC, Pruijn GJM. NETosis, complement, and coagulation: a triangular relationship. Cell Mol Immunol. 2019 Jan;16(1):19–27.

15. Denning NL, Aziz M, Gurien SD, Wang P. DAMPs and NETs in Sepsis. Front Immunol. 2019 Oct 30;10:2536.

16. Barrett T, Wilhite SE, Ledoux P, Evangelista C, Kim IF, Tomashevsky M, et al. NCBI GEO: archive for functional genomics data sets—update. Nucleic Acids Research. 2013 Jan 1;41(D1):D991–5.

17. Leek JT, Johnson WE, Parker HS, Jaffe AE, Storey JD. The sva package for removing batch effects and other unwanted variation in high-throughput experiments. Bioinformatics. 2012 Mar 15;28(6):882–3.

18. Gentleman R. Category: Cateogory Analysis [Internet]. 2023. Available from: https://bioconductor.org/packages/Category

19. Kanehisa M, Goto S. KEGG: Kyoto Encyclopedia of Genes and Genomes. Nucleic Acids Res. 2000 Jan 1;28(1):27–30.

20. Tibshirani R. Regression Shrinkage and Selection via the Lasso. Journal of the Royal Statistical Society Series B (Methodological). 1996;58(1):267–88.

21. Friedman JH, Hastie T, Tibshirani R. Regularization Paths for Generalized Linear Models via Coordinate Descent. Journal of Statistical Software. 2010 Feb 2;33:1–22.

22. Robin X. pROC: an open-source package for R and S+ to analyze and compare ROC curves [Internet]. 2011. Available from: https://xrobin.github.io/pROC/

23. Barbie DA, Tamayo P, Boehm JS, Kim SY, Moody SE, Dunn IF, et al. Systematic RNA interference reveals that oncogenic KRAS-driven cancers require TBK1. Nature. 2009 Nov 5;462(7269):108–12.

24. Hänzelmann S, Castelo R, Guinney J. GSVA: gene set variation analysis for microarray and RNA-Seq data. BMC Bioinformatics. 2013 Jan 16;14(1):7.

25. Yu G, Wang LG, Han Y, He QY. clusterProfiler: an R Package for Comparing Biological Themes Among Gene Clusters. OMICS. 2012 May;16(5):284–7.

26. The Gene Ontology (GO) database and informatics resource. Nucleic Acids Res. 2004 Jan 1;32(Database issue):D258–61.

27. MacKinnon DP, Fairchild AJ, Fritz MS. Mediation Analysis. Annu Rev Psychol. 2007;58:593.

28. Rosseel Y. lavaan: An R Package for Structural Equation Modeling. Journal of Statistical Software. 2012 May 24;48:1–36.

29. Szklarczyk D, Gable AL, Nastou KC, Lyon D, Kirsch R, Pyysalo S, et al. The STRING database in 2021: customizable protein-protein networks, and functional characterization of user-uploaded gene/measurement sets. Nucleic Acids Res. 2021 Jan 8;49(D1):D605–12.

30. Newman AM, Liu CL, Green MR, Gentles AJ, Feng W, Xu Y, et al. Robust enumeration of cell subsets from tissue expression profiles. Nat Methods. 2015 May;12(5):453–7.

31. R Core Team RCT. R: A Language and Environment for Statistical Computing [Internet]. Vienna, Austria: R Foundation for Statistical Computing; 2023. Available from: https://www.R-project.org/

32. Kolde R. pheatmap: Pretty Heatmaps [Internet]. 2018. Available from: https://github.com/raivokolde/pheatmap

33. Wei and Simko T and V. R package “corrplot”: Visualization of a Correlation Matrix [Internet]. 2024. Available from: https://github.com/taiyun/corrplot

34. Blighe K, Rana S, Lewis M. EnhancedVolcano: Publication-ready volcano plots with enhanced colouring and labeling. [Internet]. 2024. Available from: https://github.com/kevinblighe/EnhancedVolcano

35. Hadley Wickham. ggplot2: Elegant Graphics for Data Analysis [Internet]. 2016. Available from: https://ggplot2.tidyverse.org

36. Wynn JL, Cvijanovich NZ, Allen GL, Thomas NJ, Freishtat RJ, Anas N, et al. The Influence of Developmental Age on the Early Transcriptomic Response of Children with Septic Shock. Mol Med. 2011 Nov;17(11–12):1146–56.

37. Wynn JL, Guthrie SO, Wong HR, Lahni P, Ungaro R, Lopez MC, et al. Postnatal Age Is a Critical Determinant of the Neonatal Host Response to Sepsis. Mol Med. 2015 Jun 2;21(1):496–504.

38. Papayannopoulos V. Neutrophil extracellular traps in immunity and disease. Nat Rev Immunol. 2018 Feb;18(2):134–47.

39. Clark SR, Ma AC, Tavener SA, McDonald B, Goodarzi Z, Kelly MM, et al. Platelet TLR4 activates neutrophil extracellular traps to ensnare bacteria in septic blood. Nat Med. 2007 Apr;13(4):463–9.

40. Margraf S, Lögters T, Reipen J, Altrichter J, Scholz M, Windolf J. Neutrophil-derived circulating free DNA (cf-DNA/NETs): a potential prognostic marker for posttraumatic development of inflammatory second hit and sepsis. Shock. 2008 Oct;30(4):352–8.

41. Maruchi Y, Tsuda M, Mori H, Takenaka N, Gocho T, Huq MA, et al. Plasma myeloperoxidase-conjugated DNA level predicts outcomes and organ dysfunction in patients with septic shock. Critical Care. 2018 Jul 13;22(1):176.

42. Denorme F, Rustad JL, Portier I, Crandell JL, de Araujo CV, Cody MJ, et al. Neutrophil extracellular trap inhibition improves survival in neonatal mouse infectious peritonitis. Pediatr Res. 2023 Mar;93(4):862–9.

43. Kumar S, Payal N, Srivastava VK, Kaushik S, Saxena J, Jyoti A. Neutrophil extracellular traps and organ dysfunction in sepsis. Clinica Chimica Acta. 2021 Dec 1;523:152–62.

44. Caraballo C, Jaimes F. Organ Dysfunction in Sepsis: An Ominous Trajectory From Infection To Death. Yale J Biol Med. 2019 Dec 20;92(4):629–40.

45. Zhang Y, Wu C, Li L, Pandeya A, Zhang G, Cui J, et al. Extracellular Histones Trigger Disseminated Intravascular Coagulation by Lytic Cell Death. Int J Mol Sci. 2022 Jun 18;23(12):6800.

46. Marchant EA, Boyce GK, Sadarangani M, Lavoie PM. Neonatal Sepsis due to Coagulase-Negative Staphylococci. Clin Dev Immunol. 2013;2013:586076.

47. Raymond SL, López MC, Baker HV, Larson SD, Efron PA, Sweeney TE, et al. Unique transcriptomic response to sepsis is observed among patients of different age groups. PLOS ONE. 2017 Sep 8;12(9):e0184159.

48. Easton AS, Dorovini-Zis K. The kinetics, function, and regulation of P-selectin expressed by human brain microvessel endothelial cells in primary culture. Microvasc Res. 2001 Nov;62(3):335–45.

49. Zarbock A, Polanowska-Grabowska RK, Ley K. Platelet-neutrophil-interactions: linking hemostasis and inflammation. Blood Rev. 2007 Mar;21(2):99–111.

50. Mosad E, Elsayh KI, Eltayeb AA. Tissue factor pathway inhibitor and P-selectin as markers of sepsis-induced non-overt disseminated intravascular coagulopathy. Clin Appl Thromb Hemost. 2011 Feb;17(1):80–7.

51. Etulain J, Martinod K, Wong SL, Cifuni SM, Schattner M, Wagner DD. P-selectin promotes neutrophil extracellular trap formation in mice. Blood. 2015 Jul 9;126(2):242–6.

52. Leung HHL, Perdomo J, Ahmadi Z, Yan F, McKenzie SE, Chong BH. Inhibition of NADPH oxidase blocks NETosis and reduces thrombosis in heparin-induced thrombocytopenia. Blood Adv. 2021 Dec 13;5(23):5439–51.

53. Lopes-Pires ME, Frade-Guanaes JO, Quinlan GJ. Clotting Dysfunction in Sepsis: A Role for ROS and Potential for Therapeutic Intervention. Antioxidants. 2022 Jan;11(1):88.

54. You G, Zhao X, Liu J, Yao K, Yi X, Chen H, et al. Machine learning-based identification of CYBB and FCAR as potential neutrophil extracellular trap-related treatment targets in sepsis. Front Immunol [Internet]. 2023 Oct 13 [cited 2024 May 22];14. Available from: https://www.frontiersin.org/journals/immunology/articles/10.3389/fimmu.2023.1253833/full

55. Tiseo G, Cavarretta E, Forniti A, Nocella C, Sciarretta S, Spagnolello O, et al. Interplay between Nox2 Activity and Platelet Activation in Patients with Sepsis and Septic Shock: A Prospective Study. Oxidative Medicine and Cellular Longevity. 2020;2020(1):4165358.

56. Foster KG, Fingar DC. Mammalian Target of Rapamycin (mTOR): Conducting the Cellular Signaling Symphony. J Biol Chem. 2010 May 7;285(19):14071–7.

57. McInturff AM, Cody MJ, Elliott EA, Glenn JW, Rowley JW, Rondina MT, et al. Mammalian target of rapamycin regulates neutrophil extracellular trap formation via induction of hypoxia-inducible factor 1 α. Blood. 2012 Oct 11;120(15):3118–25.

58. Cheng MM W, Long Y, Wang H, Han MM W, Zhang J, Cui N. Role of the mTOR Signalling Pathway in Human Sepsis-Induced Myocardial Dysfunction. Canadian Journal of Cardiology. 2019 Jul 1;35(7):875– 83.

59. Snäll J, Linnér A, Uhlmann J, Siemens N, Ibold H, Janos M, et al. Differential neutrophil responses to bacterial stimuli: Streptococcal strains are potent inducers of heparin-binding protein and resistin-release. Sci Rep. 2016 Feb 18;6(1):21288.

60. Tapper H, Karlsson A, Mörgelin M, Flodgaard H, Herwald H. Secretion of heparin-binding protein from human neutrophils is determined by its localization in azurophilic granules and secretory vesicles. Blood. 2002 Mar 1;99(5):1785–93.

61. Fisher J, Linder A. Heparin-binding protein: a key player in the pathophysiology of organ dysfunction in sepsis. J Intern Med. 2017 Jun;281(6):562–74.

62. Zhou Y, Liu Z, Huang J, Li G, Li F, Cheng Y, et al. Usefulness of the heparin-binding protein level to diagnose sepsis and septic shock according to Sepsis-3 compared with procalcitonin and C reactive protein: a prospective cohort study in China. BMJ Open. 2019 Apr 23;9(4):e026527.

63. von Knethen A, Brüne B. Histone Deacetylation Inhibitors as Therapy Concept in Sepsis. Int J Mol Sci. 2019 Jan 16;20(2):346.

64. Li Y, Wan D, Luo X, Song T, Wang Y, Yu Q, et al. Circulating Histones in Sepsis: Potential Outcome Predictors and Therapeutic Targets. Front Immunol. 2021 Mar 24;12:650184.

65. Iqbal F, Chandra P, Khan AA, Edward S Lewis L, Acharya D, Vandana KE, et al. Prediction of mortality among neonates with sepsis in the neonatal intensive care unit: A machine learning approach. Clinical Epidemiology and Global Health. 2023 Nov 1;24:101414.

66. Safdari R, Kadivar M, Langarizadeh M, Nejad AF, Kermani F. Developing a Fuzzy Expert System to Predict the Risk of Neonatal Death. Acta Inform Med. 2016 Feb;24(1):34–7.

67. Cooper JN, Minneci PC, Deans KJ. Postoperative neonatal mortality prediction using superlearning. J Surg Res. 2018 Jan;221:311–9.

68. Beluzo CE, Silva E, Alves LC, Bresan RC, Arruda NM, Sovat R, et al. Towards neonatal mortality risk classification: A data-driven approach using neonatal, maternal, and social factors. Informatics in Medicine Unlocked. 2020 Jan 1;20:100398.

69. Hsu JF, Chang YF, Cheng HJ, Yang C, Lin CY, Chu SM, et al. Machine Learning Approaches to Predict In-Hospital Mortality among Neonates with Clinically Suspected Sepsis in the Neonatal Intensive Care Unit. Journal of Personalized Medicine. 2021 Aug;11(8):695.

